# MACHINE: a robust and scalable multi-ancestry fine-mapping method using a continuous global-local shrinkage prior

**DOI:** 10.1101/2025.09.28.25336857

**Authors:** Xiang Li, Zewei Xiong, Pak Chung Sham, Yan Dora Zhang

## Abstract

Fine mapping aims to identify causal genetic variants with nonzero phenotypic effects. Leveraging genome-wide association study (GWAS) data from diverse ancestries enhances fine-mapping accuracy and resolution by exploiting differences in linkage disequilibrium (LD) and increasing sample sizes. However, existing multi-ancestry fine-mapping methods rely on discrete priors and assume that all causal variants are shared across ancestries – an assumption that may not hold in practice. Although MESuSiE accounts for both shared and ancestry-specific causal effects, it requires a priori specification of prior probabilities for causal variant sharing. Moreover, methods based on discrete priors are prone to sub-optimal convergence. To address these limitations, we introduce Multi-AnCestry Heritability INducEd Dirichlet decomposition (MACHINE), a flexible Bayesian framework that employs a continuous prior to model both shared and ancestry-specific effects without restrictive assumptions. Importantly, we propose an approach to control false discovery rate (FDR) for fine mapping with GWAS summary statistics and out-of-sample LD matrices, a challenge not addressed by existing multi-ancestry fine-mapping methods. We further improve fine-mapping performance by incorporating functional annotations of variants using generalized LD score regression (g-LDSC). Simulation studies across diverse genetic architectures demonstrate robustness and superior FDR control of MACHINE + g-LDSC compared to existing methods. In the real data analyses, we applied MACHINE + g-LDSC to four lipid traits and schizophrenia, identifying previously unknown causal variants and depicting their genetic architectures across ancestries.

## Introduction

Genome-wide association studies (GWAS) have successfully identified numerous genetic variants associated with a wide range of complex traits and diseases^1^. However, most of these variants are not causal but rather in linkage disequilibrium (LD) with the true causal variants. Statistical fine mapping aims to identify variants with nonzero effects on the trait, a task that is essentially a variable selection problem. To date, most fine-mapping studies have focused on populations of European (EUR) ancestry. However, the extensive LD among single nucleotide polymorphisms (SNPs) in EUR populations often hinders the precise identification of causal variants^2^. One promising strategy to improve fine-mapping resolution and power is to leverage multiple GWAS datasets from diverse ancestries. On one hand, differences in LD structures across ancestries can help refine causal variant identification and reduce the size of Bayesian fine-mapping credible sets (CSs)^2,3^. On the other hand, GWAS datasets from African (AFR) and East Asian (EAS) ancestries often suffer from limited sample sizes. Multi-ancestry fine mapping can increase the effective sample size and improve power across these populations.

The shared genetic architecture across ancestries serves as the fundamental principle of multi-ancestry fine mapping. Most existing multi-ancestry fine-mapping methods rely on a strong assumption that all causal variants are shared across all ancestries. However, growing evidence suggests that genetic architectures for certain traits may exhibit substantial heterogeneity across ancestries. For instance, around 40% of causal variants for high-density lipoprotein in EUR populations are not causal in EAS populations^4^. Similarly, a trans-ancestry GWAS of major depressive disorder reported a genetic correlation of only 0.41 between EUR and EAS cohorts^5^. Such population-specific effects may arise from stronger gene-environment interaction at loci influenced by selection pressures, particularly positive selection^6^. The assumption of all causal variants are shared across ancestries fails to account for this heterogeneity, potentially leading to false discoveries.

So far, several multi-ancestry fine-mapping methods have been developed, including PAINTOR^7^, MsCAVIAR^8^, MGfm^9^, XMAP^10^, MESuSiE^11^, SuSiEx^12^, and MultiSuSiE^13^. Among these methods, XMAP, MESuSiE, SuSiEx, and MultiSuSiE are all multi-ancestry extensions of SuSiE^14,15^, differing in their modeling assumptions, prior specifications, and posterior inference approaches (Table 1). In summary, MESuSiE is currently the only method that can model ancestry-specific causal variants. However, it relies on pre-specified hyper-parameters that define the prior probabilities of a causal variant being shared across ancestries or ancestry-specific. Mis-specification of these prior probabilities may lead to reduced performance. Additionally, although MESuSiE can, in principle, be extended to more than two ancestries, both its number of parameters and computational complexity grow exponentially with the number of ancestries, posing scalability challenges. In contrast, XMAP, SuSiEx, and MultiSuSiE assume the causal status of variants is shared across ancestries. Although XMAP attempts to infer ancestry-specific CSs by applying the purity filter separately to each ancestry, SuSiEx computes post hoc probabilities to assess whether a CS is specific to a given ancestry, these procedures appear to contradict their underlying model assumptions and their theoretical justification remains unclear.

**Table 1:** Summary of MACHINE and SuSiE-based multi-ancestry fine-mapping methods.

| Method | Ancestry-specific<br>variant sets | Ancestry-specific<br>causal variants | Ancestry-specific CSs | FDR control<br>with out-of-sample LD |
| --- | --- | --- | --- | --- |
| MACHINE | ✓ | ✓ | ✓ | ✓ |
| MESuSiE<sup>11</sup> | ✗ | ✓ | ✓ | ✗ |
| SuSiEx<sup>12</sup> | ✗ | ✗ | ✓ | ✗ |
| XMAP<sup>10</sup> | ✗ | ✗ | ✓ | ✗ |
| MultiSuSiE<sup>13</sup> | ✗ | ✗ | ✗ | ✗ |

Ancestry-specific effects may arise from variants present in some ancestries but absent in others^16^. Nonetheless, all existing multi-ancestry fine-mapping methods can analyze only the intersection of variants measured across all ancestries. In practice, heterogeneous genotyping and imputation panels, differences in allele frequencies, and quality-control filters cause a non-negligible proportion of variants to be missing or poorly measured in some ancestries^11^. Consequently, as the number of ancestries increases, the set of variants that can be jointly modeled shrinks. In summary, there is no multi-ancestry fine-mapping method that flexibly and robustly models ancestry specificity.

In practice, obtaining individual-level GWAS data is often challenging. As an alternative, summary statistics for trait-variant associations and LD matrix derived from some reference panels are widely used for fine mapping. Nevertheless, discrepancies between the LD matrix and GWAS summary statistics can lead to miscalibrated results^2,17^. These discrepancies may be attributed to the heterogeneity between reference panel and GWAS samples, or sample size heterogeneity across summary statistics. Although most existing fine-mapping methods accommodate summary data as input, these methods can only produce reliable results when in-sample LD matrices are used. When out-of-sample LD matrices are employed, these methods fail to control the false discovery rate (FDR). To date, several approaches have been proposed to address this issue. SLALOM utilizes a quality control method to detect outliers in summary statistics based on local LD structure, but it assumes that there is at most one causal variant in the analyzed locus ^17^. CARMA introduces a Bayesian hypothesis testing procedure to detect and remove outliers associated with the discrepancies^18^. However, when the discrepancies between the LD matrix and summary statistics are global and substantial, removing a few outliers may not fully resolve the issue. Additionally, true causal variants are also likely to be detected as outliers, resulting in reduced coverage of CSs. The recently developed method h2-D2, based on a continuous global-local shrinkage prior, suggests setting a large global shrinkage parameter when using out-of-sample LD matrices^19^. However, it lacks a robust criterion for selecting this parameter. RSparsePro proposes a latent variable model, which introduces latent *z*-scores that would be obtained if the GWAS were conducted on individuals from the reference panel, and assumes that the observed GWAS *z*-scores are error-contaminated observations of the latent *z*-scores^20^. While this model accounts for global discrepancies, its performance depends on a hyper-parameter quantifying the magnitude of the discrepancy. Although RSparsePro sets this hyper-parameter by checking the convergence of algorithm, this approach does not guarantee the discrepancies are fully resolved and may still result in mis-calibrated outcomes. To date, these approaches have only been employed in single-ancestry fine-mapping methods and have not been integrated with multi-ancestry fine-mapping methods.

In this paper, we introduce a multi-ancestry fine-mapping method, termed Multi-AnCestry Heritability INducEd Dirichlet decomposition (MACHINE; Fig. **1**). MACHINE is conceptually different from existing multi-ancestry fine-mapping methods in several key aspects. First, as an extension of single-ancestry fine-mapping model h2-D2, MACHINE utilizes a continuous global-local shrinkage prior, which does not require assumptions about the number or proportion of causal variants. Second, MACHINE does not enforce the restrictive assumption that all causal variants are shared across all ancestries. Unlike MESuSiE that explicitly modeling causal variant sharing patterns, MACHINE implicitly accommodates both shared and ancestry-specific effects through a unified hierarchical prior, enabling robust performance across diverse genetic architectures. Third, MACHINE operates on ancestry-specific variant sets, eliminating the requirement for a common set of variants across ancestries. Furthermore, to address the discrepancies when out-of-sample LD matrix is used, MACHINE adopts the latent variable model proposed by RSparsePro, but introduces a novel approach for hyper-parameter specification, achieving improved FDR control. Last but not least, we incorporate functional annotations of variants by specifying the hyper-parameters of MACHINE based on the per-variant heritabilities estimated by generalized linkage disequilibrium score regression (g-LDSC)^21^, which improves fine-mapping performance by prioritizing variants that are more likely to be causal.

**Fig. 1:**
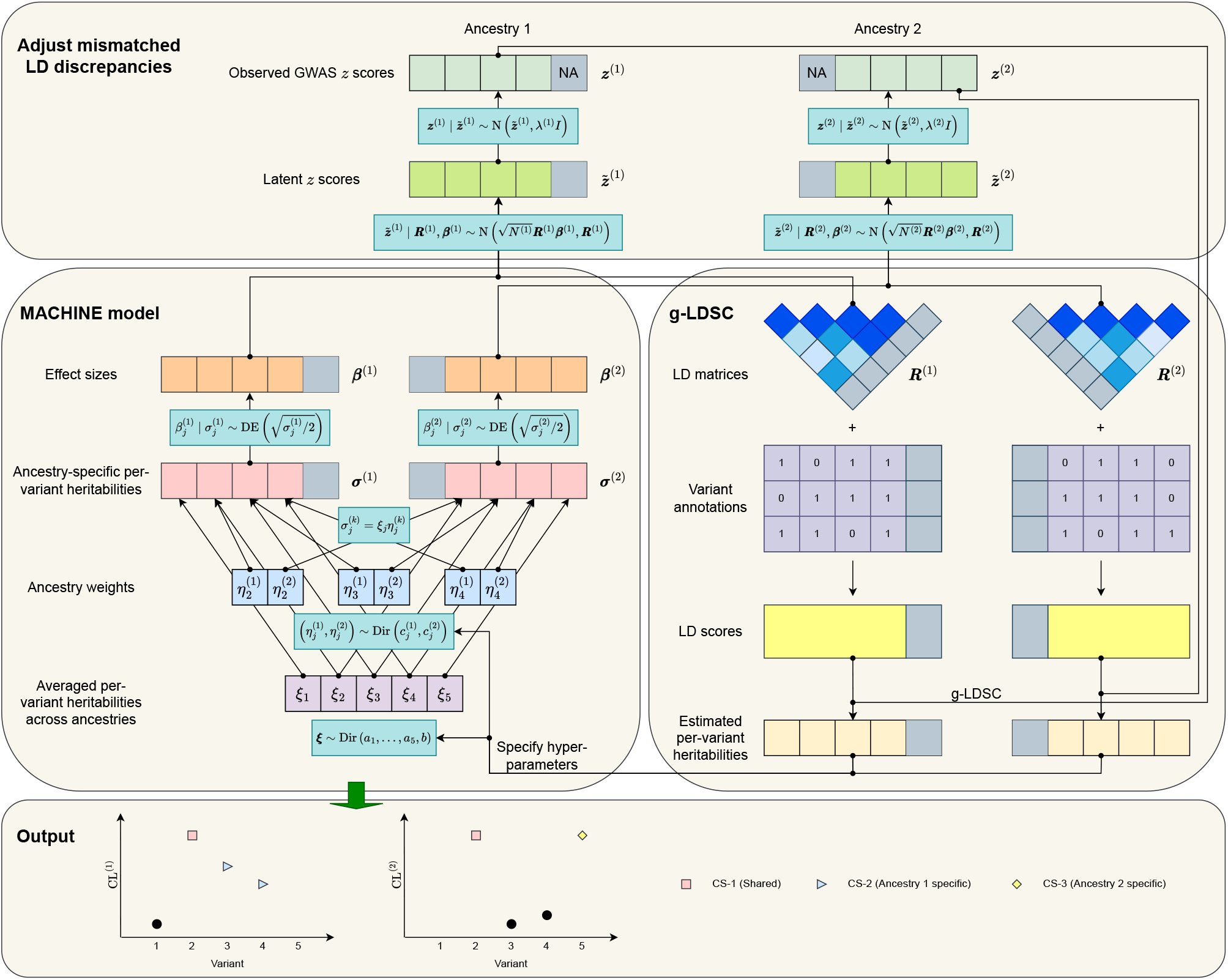
Overview of MACHINE + g-LDSC. MACHINE takes the marginal *z*-scores and LD matrices as inputs. If out-of-sample LD matrices are provided, the discrepancies between *z*-scores and LD matrices are adjusted by a latent variable model. MACHINE employs a unified hierarchical continuous global-local shrinkage prior to model both shared and ancestry-specific effects, and accommodate missing variants in some ancestries. Functional annotations of variants are incorporated by specifying the hyper-parameters of MACHINE based on the per-variant heritabilities estimated by g-LDSC. MACHINE produces credible levels (CLs) and credible sets (CSs) separately for each ancestry.

## Results

### Simulations

We conducted comprehensive simulation studies to evaluate the performance of MACHINE. Briefly, we randomly selected 200 regions on chromosome 1 and simulated GWAS summary data for variants within these regions using LD matrices of EUR and EAS ancestries derived from the UK Biobank (UKBB) reference panel (Supplementary Table 1). We considered three scenarios with varying genetic architecture heterogeneity between the two ancestries. Scenario 1: both ancestries have 5 shared causal variants in each region. Scenario 2: both ancestries have 3 shared causal variants, while each ancestry also has 1 specific causal variant in each region. Scenario 3: both ancestries have 1 shared causal variant, while each ancestry also has 2 specific causal variants in each region. Details of the simulation studies are provided in Methods.

For the primary analyses, we used in-sample LD matrices for fine mapping, and compared the performance of MACHINE with existing multi-ancestry fine-mapping methods, including MESuSiE, SuSiEx, XMAP, and MultiSuSiE, along with single-ancestry fine-mapping methods h2-D2, SuSiE, and CARMA. We also assessed the impact of incorporating variant functional annotations on the performance of MACHINE and h2-D2 using either g-LDSC^21^ or PolyFun^22^.

We evaluated the performance of all fine-mapping methods in four tasks: identifying cross-ancestry causal variants (i.e., variants with nonzero effects in at least one ancestry), identifying causal variants for each ancestry, and identifying causal variants shared by both ancestries. First, we assessed the empirical FDR and power for variants with a credible level (CL) or posterior inclusion probability (PIP) greater than or equal to 0.9 (Fig. **2**a,b, Supplementary Table 2). For identifying cross-ancestry causal variants, SuSiEx exhibited inflated FDR and significantly lower power in the settings with *N* ^(2)^ ≈ 200k, whereas all the other methods maintained well-controlled FDR. MACHINE, MESuSiE, XMAP, and MultiSuSiE demonstrated comparable power across all settings, with MACHINE being slightly less powerful. For identifying causal variants of each ancestry and shared causal variants, XMAP and MultiSuSiE displayed inflated FDR in Scenarios 2 and 3 due to their assumption that all causal variants are shared. MACHINE exhibited lower power but also lower FDR than MESuSiE and consistently outperformed SuSiEx. Notably, in Scenario 3, MESuSiE demonstrated inflated FDR in identifying EAS and shared causal variants. The power difference between MESuSiE and MACHINE in identifying EAS and shared causal variants was more pronounced when *N* ^(2)^ = 20k compared to *N* ^(2)^ = 200k. These results suggest that MESuSiE is more likely to classify a causal variant as shared than MACHINE, despite its default prior probability for a causal variant being shared (1/7) is lower than the actual values in the three scenarios (1, 3/5, and 1/5, respectively). Among single-ancestry fine-mapping methods, CARMA achieved higher power but also higher FDR compared to h2-D2 and SuSiE. Overall, MACHINE exhibited the lowest FDR among multi-ancestry fine-mapping methods, while h2-D2 had the lowest FDR among single-ancestry fine-mapping methods. MACHINE also produced better-calibrated statistics than other methods across different CL or PIP thresholds (Supplementary Fig. 1, Supplementary Table 3) and demonstrated greater power than h2-D2. As expected, the power improvement of MACHINE increased with the number of shared causal variants across all tasks, and increased with *N* ^(2)^ for cross-ancestry and EUR causal variants, but decreased with *N* ^(2)^ for EAS and shared causal variants (Supplementary Fig. 2, Supplementary Table 4). Incorporating functional annotations generally enhanced the power and reduced the FDR of both MACHINE and h2-D2 in most cases. MACHINE / h2-D2 + g-LDSC consistently outperformed MACHINE / h2-D2 + PolyFun. For both methods, we compared the estimated per-variant heritability of non-causal and causal variants. While both methods are able to prioritize causal variants, PolyFun exhibited a more dispersed per-variant heritability distribution for non-causal variants (Supplementary Fig. 3). Wilcoxon tests revealed that g-LDSC estimated per-variant heritabilities showed more significant differences between non-causal and causal variants in most cases (Supplementary Fig. 4, Supplementary Table 5).

**Fig. 2:**
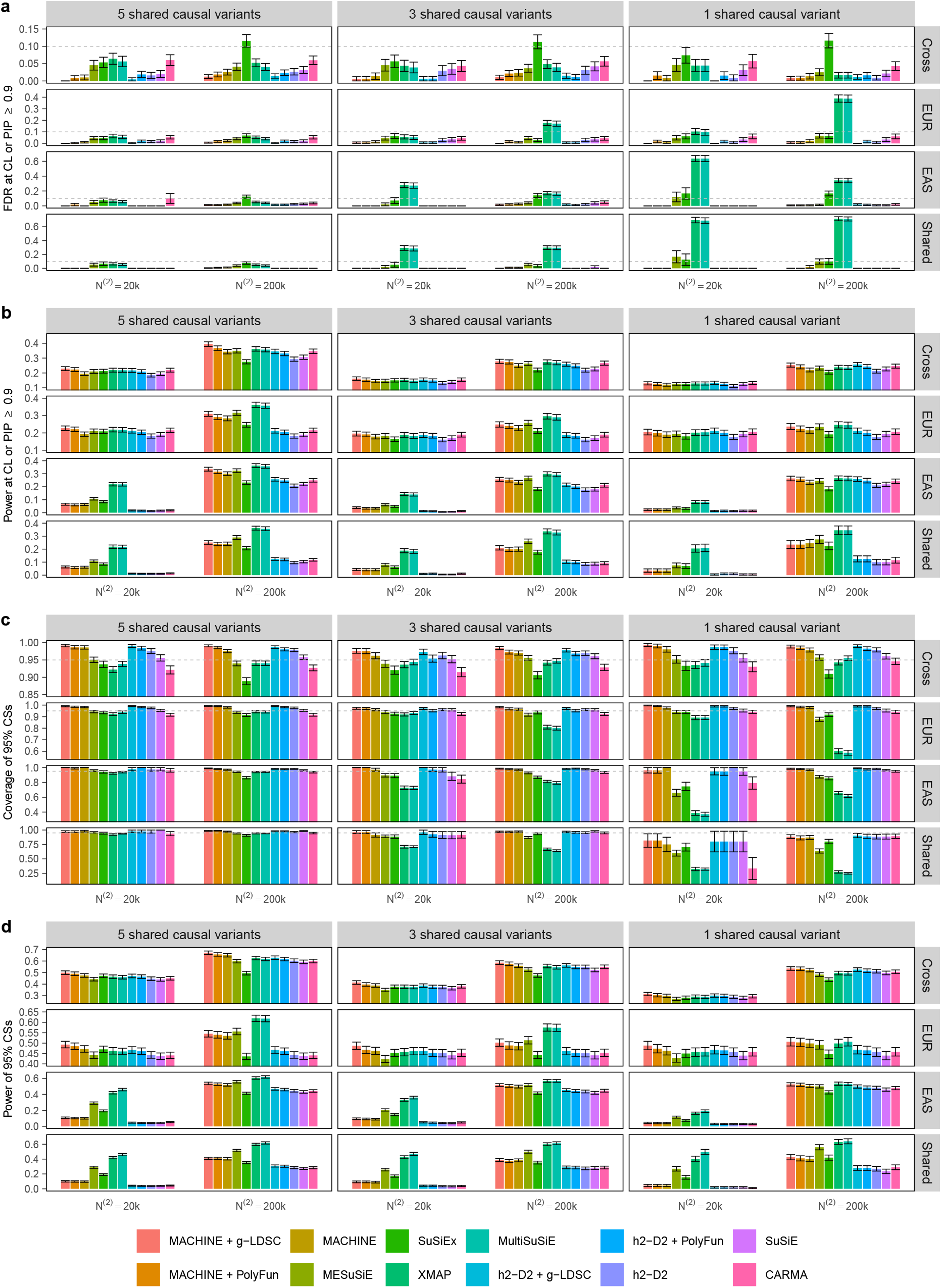
Comparison of fine-mapping methods when in-sample LD matrices are used. All metrics are calculated by aggregating results from 200 simulated datasets. Error bars represent standard errors. **a**, FDR at CL or PIP ⩾ 0.9, defined as the proportion of non-causal variants among those with CL or PIP ⩾ 0.9. **b**, Power at CL or PIP ⩾ 0.9, defined as the proportion of causal variants with CL or PIP ⩾ 0.9. **c**, Coverage of 95% CSs, defined as the proportion of CSs that contain at least one causal variant. **d**, Power of 95% CSs, defined as the proportion of causal variants included in the CSs. Numerical results are available in Supplementary Table 2.

Then, we assessed the coverage and power of 95% CSs generated by all methods (Fig. **2**c,d, Supplementary Table 2). In most cases, the 95% CSs from MESuSiE, SuSiEx, XMAP, MultiSuSiE, and CARMA failed to reach the target coverage level of 0.95. Notably, the coverage of 95% CSs from MACHINE and h2-D2 surpassed that of all other methods and exceeded 0.95 across most settings, except for shared causal variants. MACHINE’s 95% CSs exhibited the highest power in identifying cross-ancestry causal variants. For causal variants of each ancestry and shared causal variants, XMAP and MultiSuSiE demonstrated significantly lower coverage in Scenarios 2 and 3, while SuSiEx showed comparable coverage but lower power than MESuSiE. Compared to MESuSiE, MACHINE displayed lower power in 95% CSs for EAS and shared causal variants, and for EUR causal variants in Scenarios 1 and 2 when *N* ^(2)^ = 200k. For MESuSiE, the coverage of 95% CSs for causal variants of each ancestry and shared causal variants decreased with the number of shared causal variants per region. Additionally, its coverage for EUR causal variants decreased with *N* ^(2)^, while its coverage for EAS causal variants increased with *N* ^(2)^. These findings further underscore MESuSiE’s tendency to identify causal variants as shared. The coverage of 95% CSs for shared causal variants was substantially lower, especially in Scenario 3, with MESuSiE’s coverage being significantly lower than that of MACHINE, SuSiEx, and single-ancestry fine-mapping methods. In summary, these results highlight MACHINE’s robustness across scenarios with varying genetic architecture heterogeneity. Furthermore, incorporating functional annotations by g-LDSC or PolyFun improved the power of MACHINE and h2-D2 while maintaining high coverage of 95% CSs, with MACHINE / h2-D2 + g-LDSC demonstrating superior performance than MACHINE / h2-D2 + PolyFun.

As discussed in ref.^19^, h2-D2 generates CSs with larger sizes and lower purity compared to SuSiE. We evaluated the average sizes and purity of 95% CSs from MACHINE and other methods across all simulation settings (Supplementary Fig. 5, Supplementary Table 2). Overall, 95% CSs from MACHINE had smaller sizes and higher purity than those from h2-D2 but larger sizes and lower purity than those from other multi-ancestry fine-mapping methods. To investigate whether the higher coverage and power of MACHINE’s CSs were attributable solely to their larger sizes, we grouped 95% CSs from each method by their sizes and computed the coverage for each group (Supplementary Figs. 6-8, Supplementary Table 6). In most cases, MACHINE’s 95% CSs achieved the target coverage of 0.95 within each size group, whereas other multi-ancestry fine-mapping methods and CARMA exhibited lower coverage even in groups of larger sizes. These results highlight the advantage of MACHINE that sampling from the full posterior distribution provides a more comprehensive representation of uncertainty in fine-mapping results, ensuring robust performance across diverse scenarios.

Next, we evaluated the performance of MACHINE when out-of-sample LD matrices computed from 1000 Genomes reference panel were used. To date, no multi-ancestry fine-mapping method has explicitly addressed the issue of out-of-sample LD, so we compared MACHINE only with single-ancestry fine-mapping methods, including h2-D2, RSparse-Pro, and CARMA. As illustrated in Fig. **3**, MACHINE and h2-D2 exhibited lower power than RSparsePro and CARMA. However, in most cases, MACHINE and h2-D2 effectively controlled the FDR and produce 95% CSs with coverage close to or exceeding 0.95, whereas RSparsePro and CARMA showed inflated FDR and significantly lower coverage of 95% CSs. In addition, MACHINE consistently generated well-calibrated statistics across different thresholds of CL or PIP (Supplementary Fig. 9, Supplementary Table 3). Although MACHINE/h2-D2 and RSparsePro employ the same model to adjust the discrepancies in *z*-scores, they differ in their methods for assigning the hyper-parameter *λ*. Out of 1, 800 simulated single-ancestry fine-mapping datasets, h2-D2 assigned a higher *λ* than RSparsePro in 1, 648 datasets. These results indicate that RSparsePro’s hyper-parameter setting criterion – convergence of the fine-mapping algorithm – does not fully address the discrepancies. Due to the higher uncertainty assigned to *z*-scores by MACHINE and h2-D2, the average sizes of their 95% CSs were significantly larger than those from RSparsePro and CARMA (Supplementary Fig. 10, Supplementary Table 2).

**Fig. 3:**
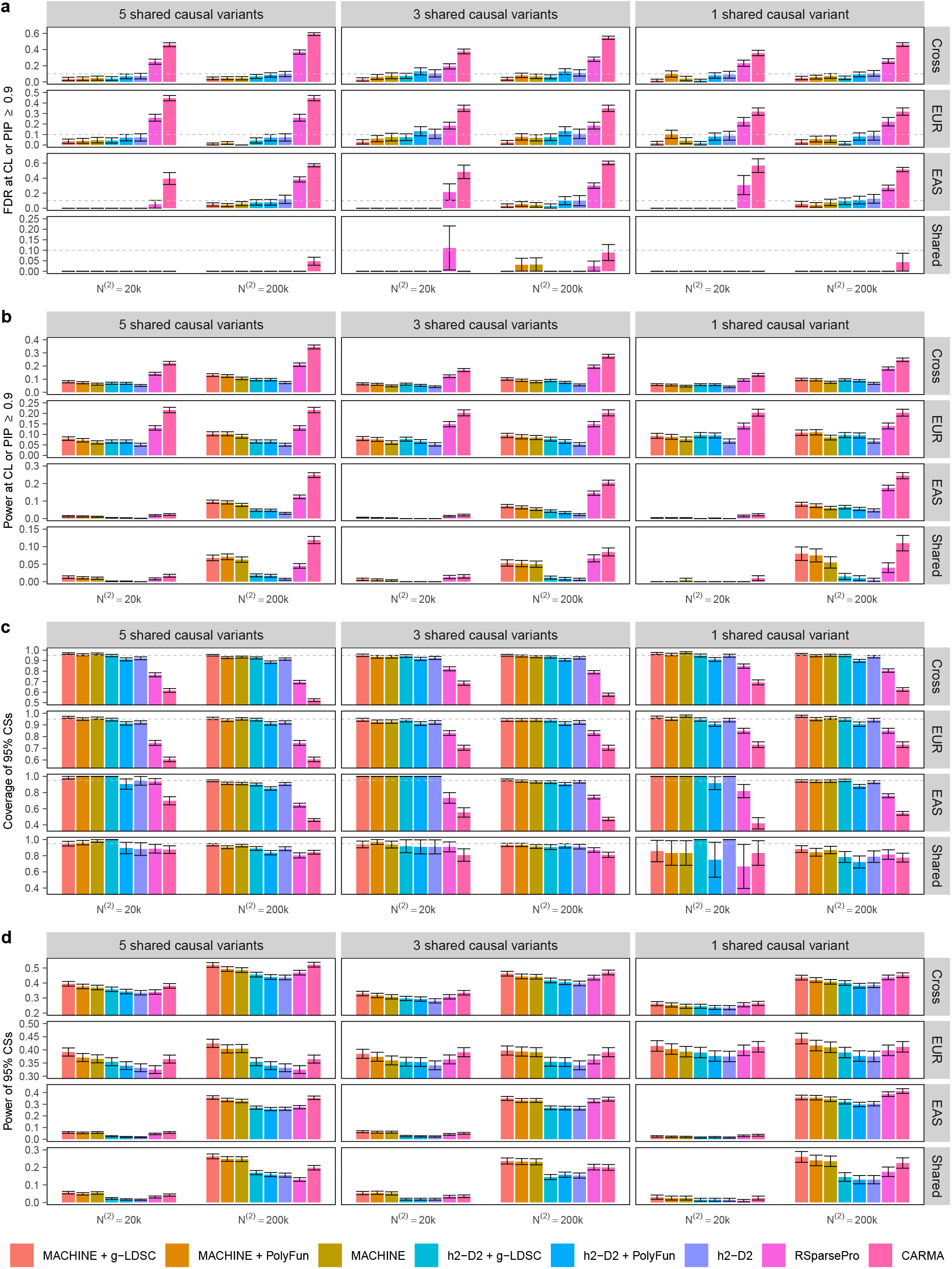
Comparison of fine-mapping methods when out-of-sample LD matrices are used. All metrics are calculated by aggregating results from 200 simulated datasets. Error bars represent standard errors. **a**, FDR at CL or PIP ⩾ 0.9, defined as the proportion of non-causal variants among those with CL or PIP ⩾ 0.9. **b**, Power at CL or PIP ⩾ 0.9, defined as the proportion of causal variants with CL or PIP ⩾ 0.9. **c**, Coverage of 95% CSs, defined as the proportion of CSs that contain at least one causal variant. **d**, Power of 95% CSs, defined as the proportion of causal variants included in the CSs. Numerical results are available in Supplementary Table 2.

When 95% CSs were grouped by size, MACHINE and h2-D2 maintained high coverage within each group (Supplementary Figs. 11-13, Supplementary Table 6). As expected, MACHINE demonstrated greater power than h2-D2 (Supplementary Fig. 14, Supplementary Table 4). Incorporating functional annotations through g-LDSC or PolyFun further enhanced the power of MACHINE and h2-D2 while maintaining low FDR of variants and high coverage of 95% CSs, even though the LD scores were derived from the 1000 Genomes reference panel, with g-LDSC consistently outperformed PolyFun (Supplementary Figs. 15-16, Supplementary Table 5).

Finally, we evaluated the computational time of each method. Theoretically, the time complexity of MACHINE and h2-D2 scales quadratically with the number of variants. This relationship was confirmed by their computational times in simulation studies (Supplementary Fig. 17, Supplementary Table 7). Similarly, SuSiEx and SuSiE exhibited quadratic complexity. In contrast, MESuSiE and XMAP demonstrated cubic complexity. CARMA, which employs a shotgun stochastic search algorithm for sampling, had linear complexity. The computational times of MultiSuSiE and RSparsePro did not show a strong linear association with the number of variants. Regarding the number of ancestries, theoretically, both MACHINE and SuSiEx scale linearly, while XMAP and MultiSuSiE scale cubically, and MESuSiE scales exponentially. Although in practice, the number of ancestries is typically limited, MACHINE’s linear scalability positions it as a more advantageous choice when data from additional ancestries become available for multi-ancestry fine mapping.

### Real data analysis for lipid traits

We applied several fine-mapping methods, including MACHINE + g-LDSC, MACHINE, MESuSiE, h2-D2 + g-LDSC, h2-D2, SuSiE, RSparsePro, and CARMA to conduct multi-ancestry or single-ancestry fine-mapping analyses for four blood lipid traits: high-density lipoprotein (HDL) cholesterol, low-density lipoprotein (LDL) cholesterol, triglycerides (TG), and total cholesterol (TC). These analyses utilized meta-analysis summary statistics from the Global Lipids Genetics Consortium (GLGC) ^23^, which integrated data across EUR (*N* = 1, 108, 161 − 1, 127, 950), AFR (*N* = 85, 162 − 89, 489), and EAS (*N* = 74, 329 − 131, 843) ancestries, aggregating results from 201 studies, including the UKBB (Supplementary Table 8). For each trait, we analyzed genomic regions containing at least one variant with a marginal association *P* < 10^−5^ in any ancestry, resulting in a total of 3,732 regions across all traits (767-1,134 per trait). LD matrices derived from the UKBB reference panel for three ancestries were used for fine mapping. The large sample sizes posed significant challenges for fine mapping with out-of-sample LD matrices, as discrepancies between *z*-scores and LD matrices could be substantial.

To evaluate the stability of fine-mapping results in real data, we adopted the replication failure rate (RFR) criterion outlined in ref. ^24^ to assess fine-mapping consistency. We used UKBB summary statistics for three ancestries obtained from Pan-UKBB as downsampled data^25^. At the variant level, the RFR was defined as the proportion of high-confidence variants (CL or PIP ⩾ 0.9) identified in UKBB fine mapping that failed to replicate (CL or PIP < 0.1) in GLGC fine mapping. We also evaluated the RFR for 95% CSs, defined as the proportion of 95% CSs identified in UKBB fine mapping that did not overlap with any 95% CSs identified in GLGC fine mapping. Across all four traits and three ancestries, MACHINE / MACHINE + g-LDSC and h2-D2 / h2-D2 + g-LDSC exhibited well-controlled RFRs for high-confidence variants, whereas other methods exhibited inflated RFRs (Fig. **4**a, Supplementary Table 9). We further compared the distributions of CL or PIP in GLGC fine mapping for variants with CL or PIP ⩾ 0.5 in UKBB fine mapping. Variants with CL ⩾ 0.5 identified by MACHINE / MACHINE + g-LDSC or h2-D2 / h2-D2 + g-LDSC in UKBB fine mapping generally retained high CL in GLGC fine mapping. In contrast, a large proportion of variants prioritized by other methods showed discordance, with many high-PIP variants in UKBB fine mapping collapsing to low PIP in GLGC fine mapping (Supplementary Figs. 18-20, Supplementary Table 10). The RFRs for 95% CSs were generally higher than those for high-confidence variants (Fig. **4**b, Supplementary Table 9). Nonetheless, the 95% CS RFRs of MACHINE / MACHINE + g-LDSC and h2-D2 / h2-D2 + g-LDSC remained lower than those of the other methods. We also examined the consistency of shared causal signals identified by each method across the two datasets. While SuSiE, RSparsePro, and CARMA identified more ancestry-specific 95% CSs than MACHINE / MACHINE + g-LDSC and h2-D2 / h2-D2 + g-LDSC, the latter methods captured more potentially shared signals with lower RFRs (Supplementary Figs. 21-23, Supplementary Tables 11-12). Although MESuSiE identified a greater number of shared signals, over half of these shared signals identified in UKBB fine mapping failed to replicate in GLGC fine mapping. These results underscore the superior robustness of MACHINE and h2-D2 for fine mapping with out-of-sample LD in real data applications.

**Fig. 4:**
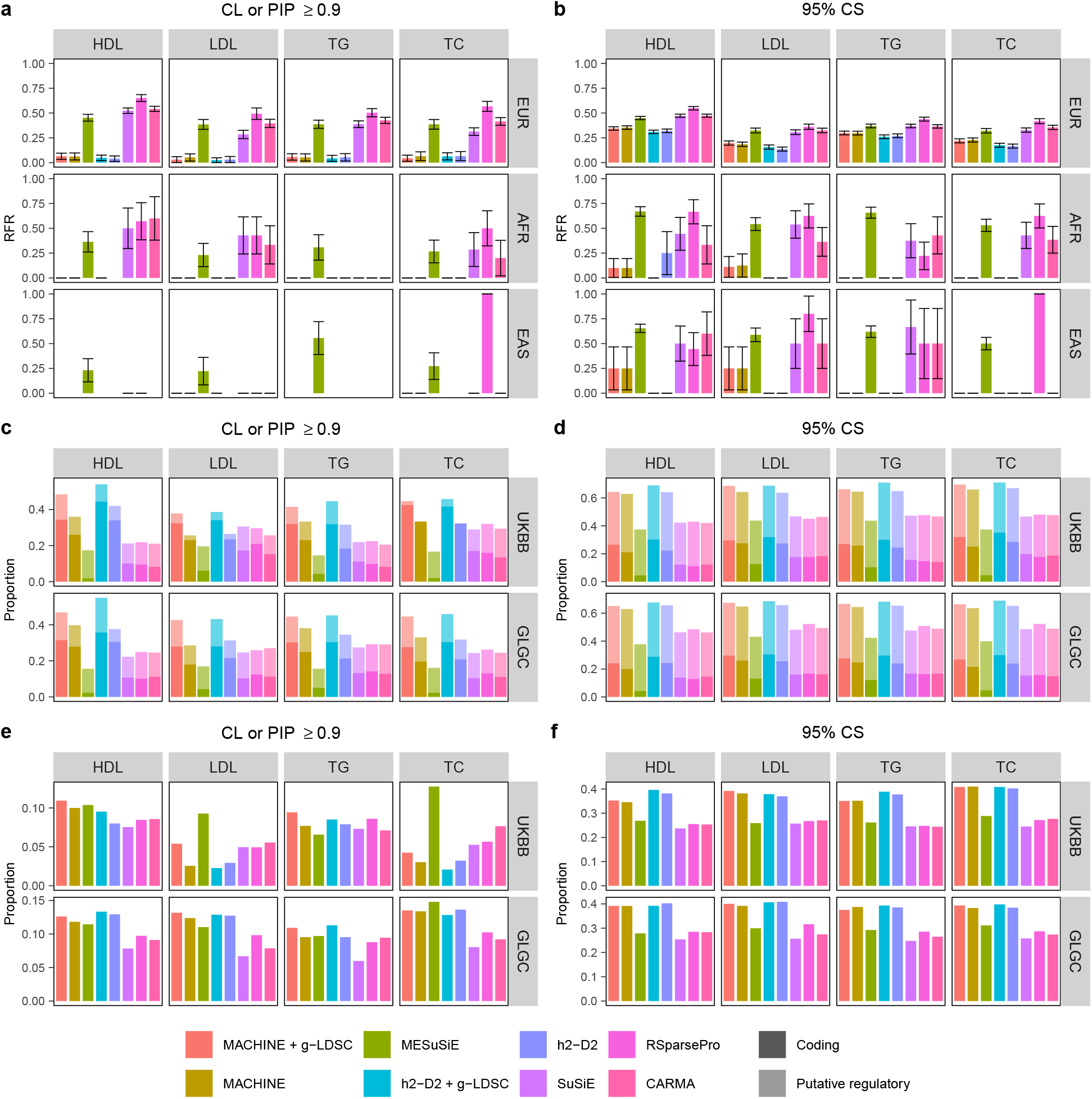
Comparison of fine-mapping methods in real data analysis of lipid traits. **a**, RFRs for variants with CL or PIP ⩾ 0.9. **b**, RFRs for 95% CSs. For **a** and **b**, error bars represent standard errors. **c**, Proportion of coding and putative regulatory variants among variants with CL or PIP ⩾ 0.9. **d**, Proportion of 95% CSs containing at least one coding variant, and proportion of 95% CSs containing at least one putative regulatory variant but no coding variant. **e**, Proportion of fine-mapped eQTLs among variants with CL or PIP ⩾ 0.9. **f**, Proportion of 95% CSs containing at least one fine-mapped eQTL. For **c**-**f**, results are aggregated across ancestries. Numerical results are available in Supplementary Tables 9 and 13.

Next, we examined the functional annotations of high-confidence variants and variants within 95% CSs identified by each method. Two categories of functional annotations not included in the g-LDSC baseline model were considered: (1) gene-based annotations, including coding and putative regulatory variants from the dbSNP database (build 151)^26,27^, and (2) fine-mapped expression quantitative trait loci (eQTLs) of adipose, liver, and whole blood tissues from the Genotype-Tissue Expression (GTEx) V10 database^28^. Overall, high-confident variants and 95% CSs identified by MACHINE / MACHINE+g-LDSC and h2-D2 / h2-D2+g-LDSC exhibited significant enrichment for coding variants, putative regulatory variants, and eQTLs compared to other methods (Fig. **4**c-f, Supplementary Table 13). Notably, functionally important variants were particularly overrepresented among high-confidence variants and 95% CSs identified by MACHINE + g-LDSC and h2-D2 + g-LDSC, suggesting that incorporating functional annotations into the prior enhances the prioritization of potential causal variants over methods relying solely on association statistics.

We present three illustrative examples of GLGC fine mapping to highlight the advantages of MACHINE + g-LDSC. The first example involves a locus on chromosome 8 (chr8:126,413,669-126,708,156) associated with TC (Figure **5**). Within this locus, two SNPs, rs28601761 (*P* ^EUR^ = 1.12 × 10^−579^) and rs112875651 (*P* ^EUR^ = 6.73 × 10^−532^, respectively), exhibit strong associations in the EUR ancestry but weak associations in the AFR ancestry (*P* ^AFR^ = 0.15 and 0.40, respectively). Using h2-D2 + g-LDSC, two 95% CSs, CS-3 and CS-4, were identified in EUR. CS-3 contains only the lead SNP rs28601761. However, neither rs28601761, rs112875651, nor the variants in CS-4 harbor functional annotations related to nearby genes. In contrast, MACHINE + g-LDSC identified two 95% CSs, CS-1 and CS-2, which are shared across all three ancestries and do not overlap with CS-3 or CS-4. CS-1 contains a single variant, rs2980888 (*P* ^EUR^ = 1.07 × 10^−283^, *P* ^AFR^ = 2.87 × 10^−10^, and *P* ^EAS^ = 8.95 × 10^−29^), which is an eQTL of the long non-coding RNA gene *TRIB1AL* in liver tissue (*P =* 4.46 × 10^−7^). *TRIB1AL* is associated with the protein-coding gene *TRIB1*, whose ortholog regulates hepatic lipogenesis and very low density lipoprotein production in mice^29^. The variant with the highest CL within CS-2 is rs8180991 (*P* ^EUR^ = 1.38 × 10^−163^, *P* ^AFR^ = 6.70 × 10^−3^, and *P* ^EAS^ = 7.62 × 10^−13^), which is located within an enhancer linked to *TRIB1* ^30^. These findings suggest that CS-1 and CS-2 are more likely to capture true causal variants. The strong marginal associations of rs28601761 and rs112875651 in EUR can be attributed to their moderate LD with rs2980888 and rs8180991. Nevertheless, in AFR and EAS, the LD is week, resulting in less significant marginal associations for rs28601761 and rs112875651 and enabling the putative causal variants to be prioritized in multi-ancestry fine mapping.

**Fig. 5:**
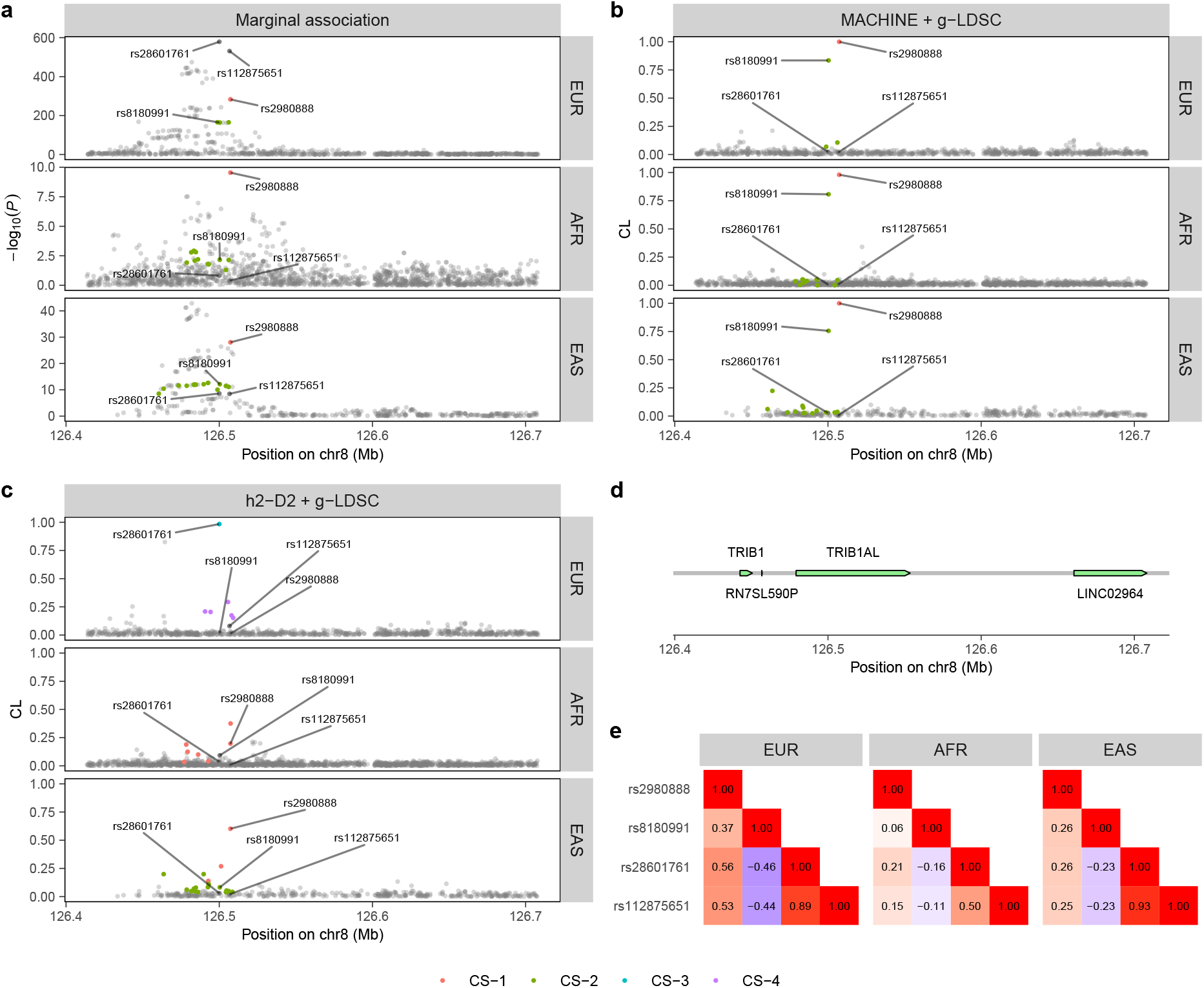
LocusZoom plot of fine-mapping results for TC in chr8:126,413,669-126,708,156. Each color represents a 95% CS. **a**, Two-sided −log_10_(*P* value) from GWAS. 95% CSs identified by MACHINE + g-LDSC are colored. **b**, CLs of variants obtained by MACHINE + g-LDSC. **c**, CLs of variants obtained by h2-D2 + g-LDSC. **d**, Gene positions within the locus. **e**, LD heatmaps of lead variants for each ancestry.

**Fig. 6:**
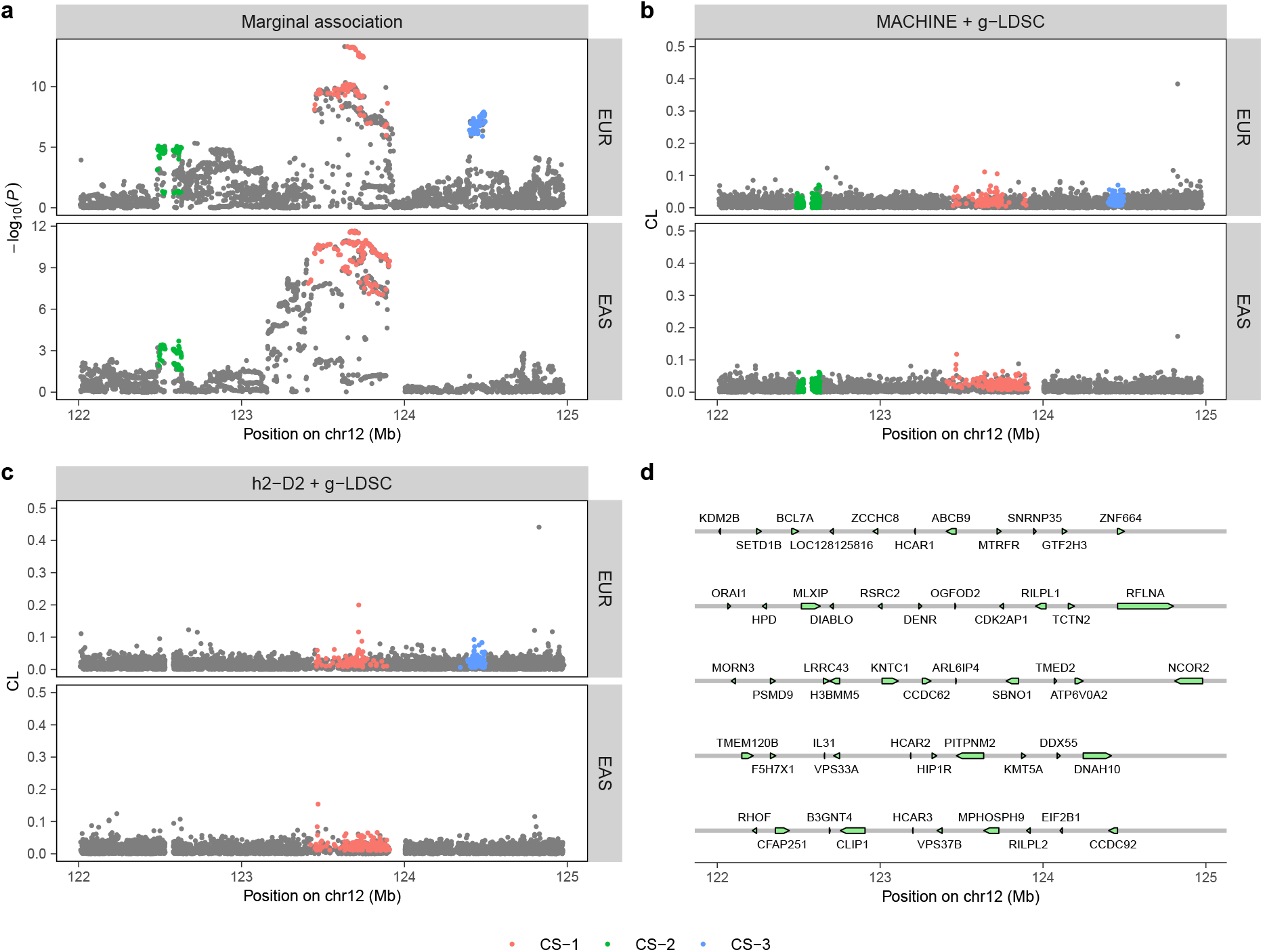
LocusZoom plot of fine-mapping results for SCZ in chr12:122,009,818-124,979,851. Each color represents a 95% CS. **a**, Two-sided −log_10_(*P* value) in GWAS. 95% CSs identified by MACHINE + g-LDSC are colored. **b**, CLs of variants obtained by MACHINE + g-LDSC. **c**, CLs of variants obtained by h2-D2 + g-LDSC. **d**, Gene positions within the locus.

The second example focuses on a locus on chromosome 19 (chr19:11,283,869-11,527,628) associated with LDL (Extended Data Fig. **1**). In this locus, h2-D2 + g-LDSC identified two 95% CSs for the EUR ancestry, CS-1 and CS-2. Whereas the variants in these CSs are located in the intronic region or 5’ flanking region of *DOCK6*, existing studies suggest that the target gene of this GWAS hit is more likely to be *ANGPTL8* rather than *DOCK6* ^31–33^. Several variants in CS-1 and CS-2 are located within enhancers of *ANGPTL8*. MACHINE + g-LDSC not only identified these two CSs but also detected an additional CS, CS-3, which is shared by all three ancestries. Variants in CS-3 are less significant than those in CS-1 in EUR but are among the most significant variants in this locus in AFR and EAS. CS-3 is located upstream of *ANGPTL8*, further supporting its potential regulatory role. Additionally, both MACHINE + g-LDSC and h2-D2 + g-LDSC identified a CS specific to the AFR ancestry, CS-4. This CS contains two variants, rs141803407 and rs140028947, which exhibit significant marginal associations with LDL in AFR (*P* ^AFR^ = 4.43 × 10^−32^ and 1.60 × 10^−30^, respectively) but week associations in EUR (*P* ^EUR^ = 0.420 and 0.0534, respectively). These two variants are located within an intron of *TSPAN16* and within an enhancer that regulates several nearby genes. The relationship between these genes and LDL remains unclear and deserves further investigation. This example highlights the ability of MACHINE + g-LDSC to identify additional shared and ancestry-specific causal signals, providing a more comprehensive understanding of potential causal variants.

The third example demonstrates the benefits of incorporating functional annotations into the fine-mapping analysis. For TG in the locus chr17:63,513,466-64,800,552, both MACHINE + g-LDSC and MACHINE identified three 95% CSs (Extended Data Fig. **2**). CS-1 is shared by EUR and AFR ancestries, CS-2 is shared by EUR and EAS ancestries, and CS-3 is specific to EUR. Within these CSs, rs1801689 in CS-1 and rs1801690 CS-2 are likely to be the causal variants, since both of them are missense variants of *APOH*, a gene involved in TG metabolism^34^. While MACHINE included these two variants in the CSs, there CLs were relative low (CL^EUR^ = 0.238 and 0.236). In contrast, MACHINE + g-LDSC successfully prioritized these two variants (CL^EUR^ = 0.617 and 0.506) by assigning larger shape parameters to them.

### Real data analysis for schizophrenia

We further applied MACHINE + g-LDSC and other methods on schizophrenia (SCZ) using GWAS summary statistics of EUR and EAS from the Psychiatric Genomics Consortium^35^. The sample sizes of EUR and EAS are 130,644 (53,386 cases and 77,258 controls) and 30761 (14,004 cases and 16,757 controls), respectively. Our analysis focused on 571 genomic loci that harbor at least one variant with a marginal association *P* < 10^−5^ in any ancestry.

In summary, MACHINE + g-LDSC identified 366 EUR-specific causal signals, 40 EAS-specific causal signals, and 31 causal signals shared by EUR and EAS (Extended Data Fig. **3**a, Supplementary Table 14). MACHINE + g-LDSC identified more shared signals than MACHINE and single-ancestry fine-mapping methods, but significantly fewer than MESuSiE. Variants with CL ⩾ 0.5 and 95% CSs identified by MACHINE / MACHINE + g-LDSC and h2-D2 / h2-D2 + g-LDSC exhibits a significant enrichment of coding variants, putative regulatory variants, and fine-mapped brain eQTLs, surpassing the enrichment level of those identified by MESuSiE, SuSiE, RSparsePro, and CARMA (Extended Data Fig. **3**b-e, Supplementary Table 15), demonstrating the effectiveness of our integrative approach in prioritizing functionally important variants.

We provided an example to illustrate the advantages of MACHINE + g-LDSC. In the gene-dense region chr12:122,009,818-124,979,851, h2-D2 + g-LDSC detected a CS shared by both ancestries (CS-1) as well as an EUR-specific CS (CS-3). Within CS-1, rs3825141 is a missense variant of *SBNO1*, a gene involved in neurite outgrowth of cortical neurons^36^. In CS-3, rs11057401 is a missense variant of *CCDC92*. Existing research indicates that the *CCDC92* mRNA is downregulated in SCZ patients and may play a role in competitive endogenous RNA regulatory network associated with SCZ^37^. MACHINE + g-LDSC identified an additional CS shared by two ancestries, CS-2, which was not detected by other methods. The putative target gene of CS-2 is *BCL7A*, which showed higher expression in SCZ patients compared to controls^38^.

## Discussion

We have introduced MACHINE, a multi-ancestry fine-mapping method that addresses several key challenges in multi-ancestry fine mapping. MACHINE extends the single-ancestry fine-mapping method h2-D2 to multi-ancestry contexts and adapts a hierarchi-cal continuous global-local shrinkage prior, enabling more extensive exploration of causal variant configurations and is less likely to obtain sub-optimal solutions ^19^. Notably, MACHINE does not explicitly model the patterns of casual variant sharing across ancestries, allowing for greater flexibility. MACHINE also enables modeling ancestry-specific variant sets, avoiding the requirement of modeling a common set of variants across ancestries. Simulation studies demonstrate that MACHINE achieves better control of the FDR than SuSiE-based multi-ancestry fine-mapping methods, and exhibits robust performance in identifying cross-ancestry, ancestry-specific, and shared causal variants across diverse genetic architectures with varying degrees of heterogeneity among ancestries.

Another important contribution of our work is the development of an approach to control the FDR when performing fine mapping with out-of-sample LD matrices. This approach improves the latent variable model of RSparsePro by specifying the hyper-parameter that quantifies the discrepancies between the LD matrix and summary statistics in a more principled manner. We have integrated this method into both MACHINE and h2-D2. In simulations using out-of-sample LD, high-confidence variants identified by MACHINE and h2-D2 demonstrate lower FDR compared to those identified by RSparsePro and CARMA. In real data analysis of four lipid traits, MACHINE and h2-D2 achieve lower RFR than other methods.

Furthermore, we enhanced fine mapping by incorporating functional annotations of variants using g-LDSC. Similar to PolyFun, this approach estimates per-variant heritability from functional annotations by fitting genome-wide summary statistics. Since MACHINE and h2-D2 model per-variant heritability directly, assigning hyper-parameters for MACHINE or h2-D2 based on estimated per-variant heritabilities is more appropriate than assigning prior probabilities for discrete-mixture prior-based methods like SuSiE or FINEMAP. Different from PolyFun which employs L2-regularized stratified LDSC (s-LDSC) and requires a cross-validation step to determine the regularization strength, g-LDSC utilizes a feasible generalized least-squares estimation that accounts for correlated error structure, resulting in more precise estimates of functional enrichment than s-LDSC. Simulation studies demonstrate that MACHINE / h2-D2 + g-LDSC consistently outperforms MACHINE / h2-D2 + PolyFun, even when out-of-sample LD matrices are used. In real data analyses, high-confidence variants and 95% CSs identified by MACHINE / h2-D2 + g-LDSC are more enriched for functionally important variants than those identified by MACHINE / h2-D2 and other methods.

Our study still has several limitations. First, although MACHINE accommodates missing variants in some ancestries, it produces CLs and CSs spearately for each ancestry. Consequently, the inference of shared causal signals relies on the overlap of 95% CSs across ancestries, a somewhat trivial approach that may complicate result interpretation. Second, while MACHINE effectively controls the FDR, it is less powerful than MESuSiE, XMAP, and MultiSuSiE for identifying shared causal variants. Further research is needed to leverage similarities in genetic architecture across ancestries to improve power. Third, in this study, we used allele frequencies as the only ancestry-specific functional annotation, while the remaining 52 annotations from the baseline model were shared across all ancestries. Expanding the collection of ancestry-specific functional annotations could enhance the estimation of per-variant heritabilities and facilitate the interpretation of ancestry-specific causal variants.

## Methods

### Overview of h2-D2

In this section, we review the single-ancestry fine-mapping model h2-D2^19^. Let ***y*** ∈ ℝ^*N*^ represent the standardized phenotype values for *N* individuals in a GWAS of a quantitative trait, and ***X*** ∈ ℝ^*N* × *J*^ denote their standardized genotype values for *J* genetic variants within a genomic region of interest. The relationship between the phenotype and genotypes can be modeled by a multiple linear regression model:

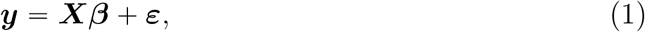

where ***β*** = (*β*_1_, …, *β*_*J*_)^⊤^ is a vector of effect sizes to be estimated, and ***ε*** is an *N* -vector of normally distributed error terms.

A double-exponential prior is assigned to each element of ***β***:

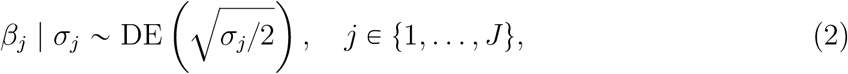

where DE(*δ*) denotes a double-exponential (Laplace) distribution with mean 0 and variance 2*δ*^2^. Thus, Var(*β*_*j*_) = *σ*_*j*_, and *σ*_*j*_ can be interpreted as the per-variant heritability of the *j*-th variant. The narrow-sense heritability *h*^2^ of the quantitative trait explained by these *J* variants is given by

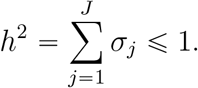

Based on this inequality, we impose a Dirichlet prior on the variance terms:

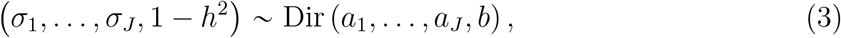

where *a*_1_, …, *a*_*J*_ ∈ (0, 1) and *b* > 0 are hyper-parameters. According to the properties of Dirichlet distribution, this prior encourages sparsity by shrinking most elements of *σ*_1_, …, *σ*_*J*_ toward zero while allowing a few to retain large values.

The h2-D2 prior can also be applied to binary traits by modeling the observed-scale heritability. An efficient Markov Chain Monte Carlo (MCMC) algorithm is developed to efficiently sample from the posterior distribution.

Unlike existing fine-mapping methods, h2-D2 outputs credible level (CL) for each variant to quantify how likely the variant is causal:

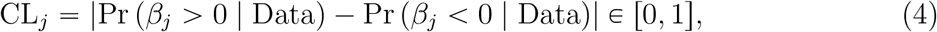

where the posterior probability 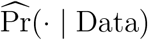 is estimated from the MCMC samples. This statistic serves a similar purpose to the posterior inclusion probability (PIP) used in discrete-mixture prior-based fine-mapping methods.

For a set of variants 𝒞 = {*j*_1_, …, *j*_*k*_}, its CL is defined as

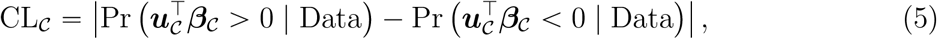

where ***u***_𝒞_ is an eigenvector of the LD matrix (i.e. correlation matrix) of variants in 𝒞 corresponding to its largest eigenvalue. If CL_𝒞_ ⩾ 1 − *α*, we can claim that 𝒞 is a level 1 − *α* credible set (CS).

### MACHINE model

In this section, we introduce the multi-ancestry fine-mapping model MACHINE. Suppose we have GWAS data of *K* ancestries. Our model allows some variant data to be missing in some ancestries. For *j* ∈ {1, …, *J*} and *k* ∈ {1, …, *K*}, let 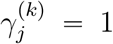 if the data of the *j*-th variant are available for the *k*-th ancestry, and 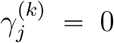 otherwise. Let 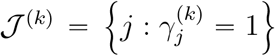 denote the index set of variants with available data in the *k*-th ancestry, while 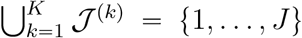, and *J*^(*k*)^ = | *J* ^(*k*)^ |. Let 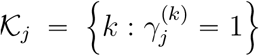 denote the index set of ancestries for which the *j*-th variant has data, and *K*_*j*_ = | *𝒦*_*j*_|.

For the *k*-th ancestry, denote 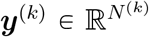 as the standardized phenotype values of *N*^(*k*)^ individuals, and 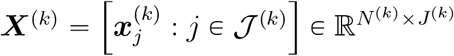 as the standardized genotype matrix. The relationship between phenotype and genotypes is modeled by the following multivariate linear regression model:

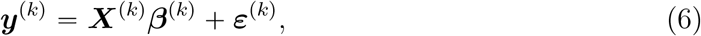

where 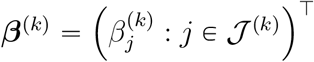 is a *J*^(*k*)^-vector of genetic effect sizes in the *k*-th ancestry, and ***ε***^(*k*)^ is an *N* ^(*k*)^-vector of normally distributed error terms. Let ***B*** = {***β***^(1)^, …, ***β***^(*K*)^} denote the effect sizes across all ancestries.

Following the framework of h2-D2, we consider a double-exponential prior for 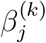,

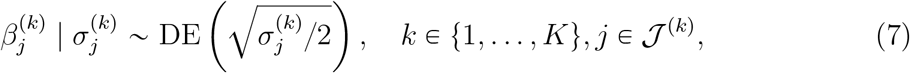

where 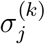 represents the per-variant heritability of the *j*-th variant in the *k*-th ancestry.

Let 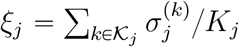 denote the averaged per-variant heritability of the *j*-th variant across ancestries, and ***ξ*** = (*ξ*_1_, …, *ξ*_*J*_)^⊤^. To achieve fine mapping, we consider a Dirichlet prior on ***ξ***, which encourages most elements of ***ξ*** to shrink toward zero while allowing some large values to be retained:

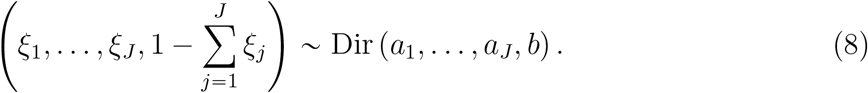

The next step is to partition the total variance of each variant into different ancestries. For {*j*: *K*_*j*_ = 1}, i.e. the variants that present in only one ancestry, 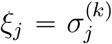, where *k* satisfies 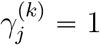, and no partition is required. For {*j*: *K*_*j*_ > 1}, let 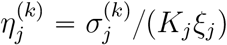 for *k* ∈ 𝒦_*j*_, which represent ancestry weights for the *j*-th variant. We have 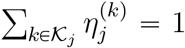. We further impose a Dirichlet prior on 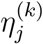:

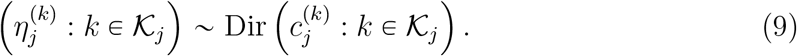

In summary, the prior distribution of 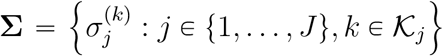 can be ex-pressed as

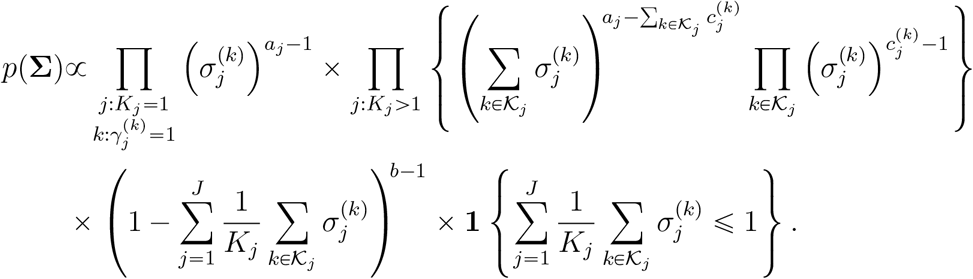

If all 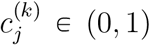 and 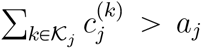, the prior induces a moderately sparse distribution over 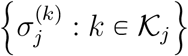, accommodating both shared and ancestry-specific causal variants.

We proposed an efficient MCMC algorithm to sample from the posterior distribution. Details are provided in the Supplementary Note.

MACHINE outputs posterior summaries for each ancestry separately. The CL of the *j*-th variant in the *k*-th ancestry is defined as

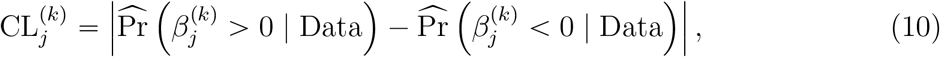

where the posterior probability 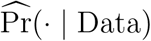 is estimated from the MCMC samples. For a set of variants 𝒞 = {*j*_1_, …, *j*_*k*_}, its CL in the *k*-th ancestry is defined as

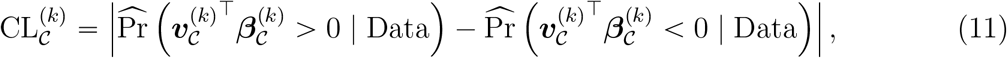

where 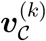 is an eigenvector of the LD matrix of variants in 𝒞 for the *k*-th ancestry corresponding to its largest eigenvalue. If 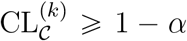, we can conclude that 𝒞 is a level 1 − *α* CS in the *k*-th ancestry.

### GWAS summary statistics

Due to ethical and practical concerns, accessing individual-level genotype and phenotype data is often challenging. Like many existing fine-mapping methods, MACHINE requires GWAS summary statistics only. For the *k*-th ancestry, let 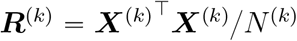 denote the in-sample LD matrix of variants in 𝒥^(*k*)^, and let 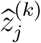 represent the single-variant *z*-score of the *j*-th variant. Define

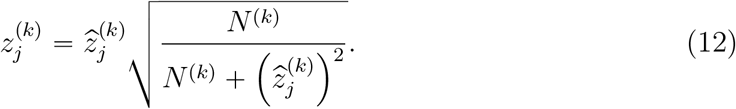

The joint probability distribution of 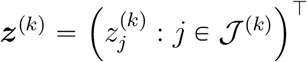 can be approximated by^39^

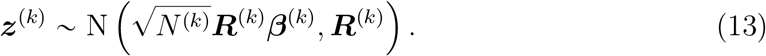

### Fine mapping with out-of-sample LD

In this section, we describe a robust approach for fine mapping with out-of-sample LD, focusing on the single-ancestry scenario for simplicity. The approach can be naturally extended to the multi-ancestry case.

Suppose the LD matrix ***R*** is estimated from some external reference panel. Let 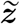 denote the hypothetical *z*-scores that would be obtained if the GWAS were conducted on individuals from the reference panel. Then,

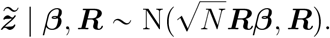

Following the RSparsePro framework^20^, we model the relationship between 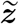 and the as observed *z*-scores ***z*** as

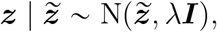

where *λ* is a hyper-parameter quantifying the discrepancy between ***z*** and quently, 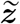. Conse-quently,

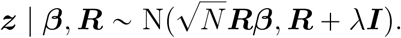

The choice of *λ* is critical for the fine-mapping results. If *λ* is set too small, fine mapping may yield an excess of false positives. RSparsePro determines *λ* solely by monitoring the convergence of the variational inference algorithm. However, algorithmic convergence does not guarantee that *λ* is sufficiently large to fully account for the discrepancies between summary statistics and the reference panel.

Here, we propose a more robust method to determine *λ*. Assume there is exactly one causal variant in the locus. The most likely causal variant is the one with the largest absolute *z*-score. Let *l* = argmax_*j*_ |*z*_*j*_|. The unbiased estimate of its effect size is

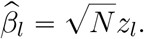

By subtracting the effect of the *l*-th variant, the residual term

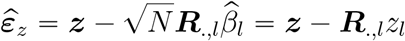

approximately follows N(0, ***R*** + *λ****I***).

To obtain a robust hyper-parameter setting for *λ*, we take the maximum of method of moments (MoM) estimate and the maximum likelihood estimate (MLE). The MoM estimate of *λ* is given by

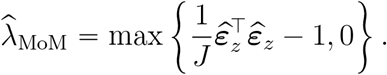

To compute the MLE, we perform eigendecomposition on ***R***, yielding ***R*** = ***UDU*** ^⊤^, where ***U*** is an orthogonal matrix whose columns are the eigenvectors of ***R***, and ***D*** = diag{*d*_1_, …, *d*_*J*_} is a diagonal matrix of eigenvalues. Consequently, we have

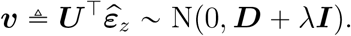

The log-likelihood function of ***v*** and its derivative are

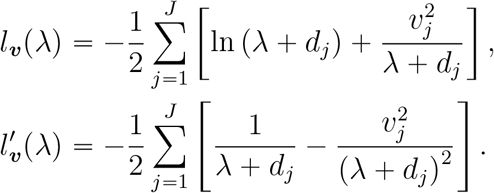

If 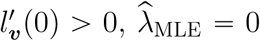. Otherwise, we use the R function “uniroot” to find the root of 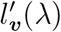. Finally, we set 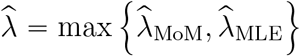.

If the assumptions are violated (e.g., if the *l*-th variant is not the true causal variant or if there are multiple causal variants in the locus), it is evident that our method would overestimate *λ*. The overestimation, however, contributes to producing robust fine-mapping results by accounting for potential discrepancies and reducing the risk of false positives.

### Summary statistics with sample size heterogeneity

In practice, GWAS summary statistics often exhibit varying sample sizes across variants. This heterogeneity can result from differences in genotyping quality control, the use of heterogeneous genotyping arrays across cohorts in meta-analyses, or the application of different imputation reference panels^17^. Performing fine mapping using summary statistics with heterogeneous sample sizes can lead to miscalibrated results. To address this issue, we propose the following data preprocessing approach.

Let *N*_*j*_ denote the sample size of the *j*-th variant. We set a threshold for the minimum acceptable sample size, for example, 90% of the maximum sample size across all variants:

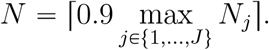

Variants with sample sizes less than *N* are excluded. For the remaining variants, we scale the *z*-scores as follows:

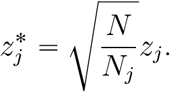

We then perform fine mapping using the scaled *z*-scores and sample size *N*, treating this scenario as fine mapping with a mismatched LD matrix.

### Choice of hyper-parameters

For Dirichlet prior (8), the hyper-parameter *a*_*j*_ controls the concentration on *ξ*_*j*_. By default, we set *a*_1_ = … = *a*_*J*_ = 0.005, which has been shown to provide robust performance in practice with varying numbers of causal variants. If external information, such as functional annotations of variants, suggests that the *j*-th variant is more likely to be causal, a larger value *a*_*j*_ can be assigned.

For Dirichlet prior (9), the hyper-parameters 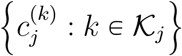 determine how the total variance of a variant is allocated across different ancestries. By default, we set 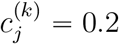. The hyper-parameter *b* represents global shrinkage. We employ an empirical Bayesian approach to determine *b* in a data-driven manner. Details of the algorithm are described in the Supplementary Note.

### Incorporating functional annotations

We incorporate functional annotations of variants by using generalized linkage disequilibrium score regression (g-LDSC)^21^ to estimate per-variant heritabilities from GWAS summary statistics. As both h2-D2 and MACHINE priors are imposed on per-variant heritabilities, it is natural to specify their hyper-parameters using these estimates.

We begin with the single-ancestry scenario. For the *j*-th variant, let ***s***_*j*_ = (*s*_*j*1_, …, *s*_*jP*_)^⊤^ denote the vector of *P* annotations, and ***S*** = [***s***_1_, …, ***s***_*J*_]^⊤^. The annotations can be binary, taking values in {0, 1}, or continuous, scaled to the [0, 1] interval.

The relationship between per-variant heritability and annotations is modeled as

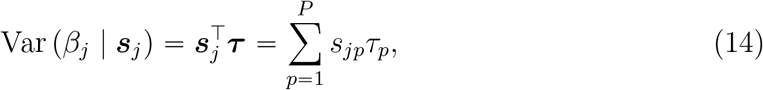

where *τ*_*p*_ represents the contribution of the *p*-th annotation to per-variant heritability.

Same as stratified LDSC (s-LDSC)^40^, g-LDSC estimates ***τ*** by regressing 𝒳^2^ statistics of variants on LD scores with respect to each functional annotation,

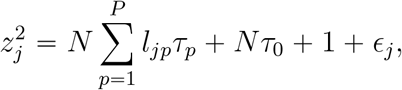

where 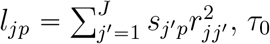 quantifies confounding biases, and *ϵ*_*j*_ denote the error term for the *j*-th variant. While s-LDSC only uses the variance of each *ϵ*_*j*_ to obtain a weighted least squares estimate of ***τ***, g-LDSC accounts for the covariance of ***ϵ*** = (*ϵ*_1_, …, *ϵ*_*J*_)^⊤^, thereby providing more precise estimates. The covariance matrix of ***ϵ*** is approximated by

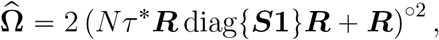

where 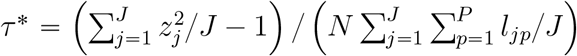.

Let 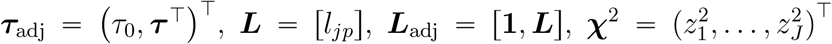. The g-LDSC estimate of ***τ***_adj_ is given by

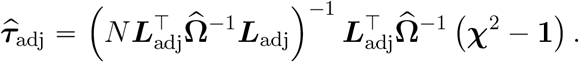

After obtaining 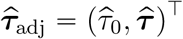, the unconstrained per-variant heritability estimate is 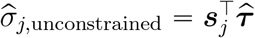. To ensure non-negativity of per-variant heritability, we let

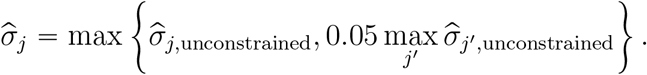

For Dirichlet prior (3), the hyper-parameter *a*_*j*_ is proportional to E (*σ*_*j*_). Thus, we specify

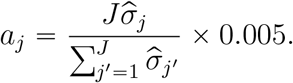

In the multi-ancestry scenario, per-variant heritabilities for each ancestry are estimated by g-LDSC separately. The averaged per-variant heritability for the *j*-th variant is 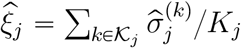. We then specify

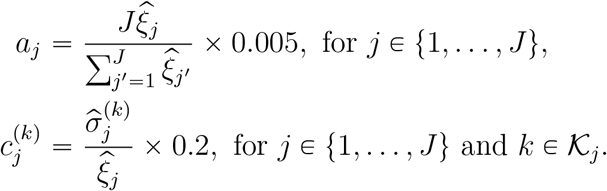

### UK Biobank data preprocessing

We filtered individuals from UKBB based on the following criteria. (1) Only individuals with available genotype data were included (data field 22005). (2) Individuals with inconsistent genetic sex (data field 22001) and self-reported sex (data field 31) were excluded. (3) Individuals recommended for exclusion from genomic analysis were removed (data field 22010). (4) Outlier individuals for heterozygosity or missing rate were excluded (data field 22027). (5) Individuals with close familial relationships were removed to avoid any potential bias in the analysis (data field 22018).

Next, we selected individuals from different ancestries based on data field 21000. Individuals with code “1”, “1001”, “1002”, or “1003” were assigned to EUR ancestry. Individuals with code “4”, “4001”, “4002”, or “4003” were assigned to AFR ancestry. Individuals with code “5” were assigned to EAS ancestry. As a result, 314, 673 EUR, 6, 492 AFR, and 1, 344 EAS individuals were selected.

For each ancestry, we filtered variants for imputed genotype data using the following criteria: (1) Only biallelic variants were included. (2) Variants with a minor allele frequency (MAF) of at least 0.1% were retained. (3) Variants with INFO score > 0.8 and Hardy-Weinberg equilibrium *P* -value > 10^−6^ were included. Genotype data preprocessing was performed using PLINK 2.0 (v2.00a3.3LM)^41^.

20 long-range LD regions were excluded from our analyses^42^. The remaining autosomal regions were partitioned into 3, 477 non-overlapping regions with approximately independent LD, based on the LD structure in EUR ancestry using the algorithm described in ref.^19^.

### 1000 Genomes Project data preprocessing

We included 502 unrelated individuals with superpopulation code “EUR” and 504 unrelated individuals with superpopulation code “EAS” from 1000 Genomes Project Phase 3 in our analysis^43,44^. For each population, variants with a MAF of 0 were removed. Genotype data preprocessing and computation of LD matrices were performed using PLINK (v1.90b6.26)^45^.

### Annotation data preprocessing

We used 54 functional annotations in the g-LDSC model. One is the base annotation, which includes all variants. The other 52 annotations are from the baseline model^40^. These annotations comprise 24 publicly available main annotations that are not specific to any cell type, such as coding, UTR, promoter, intronic regions, histone marks, and open chromatin regions, along with a 500-bp window around each main annotation and a 100-bp window around ChIP-seq peaks when appropriate. These 53 annotations are not ancestry-specific. In our real data application, we additionally considered an ancestry-specific annotation associated with the MAFs of variants in each ancestry from the UKBB reference panel, defined as 4 ∗ MAF ∗ (1 − MAF).

The gene-based annotations of variants and their associated genes were extracted from the dbSNP database (build 151) with GRCh37.p13 as the reference assembly^26^. These annotations include: NSF (non-synonymous frameshift), NSM (non-synonymous missense), NSN (non-synonymous nonsense), SYN (synonymous), R3 (in 3’ gene region), and R5 (in 5’ gene region), U3 (in 3’ UTR), U5 (in 5’ UTR), ASS (in acceptor splice site), DSS (in donor splice-site), INT (in intron). Following ref. ^24^, the “coding” category is defined as the union of NSF, NSM, and NSN, white the “putative regulatory” category is defined as the union of SYN, R3, R5, U3, and U5. Coding variants are excluded from the putative regulatory category.

SuSiE fine-mapped *cis*-eQTL data were obtained from GTEx V10 database^28^. The base-pair position, major allele, and minor allele of each variant in the GRCh37 reference were determined from the “variant_id_b37” column of the lookup table for all variants genotyped in GTEx V10.

### Simulation

We generated GWAS summary statistics using LD matrices of EUR (ancestry 1) and EAS (ancestry 2) computed from the UKBB reference panel. We selected 200 nearly independent LD blocks on chromosome 1 (supplementary). Only variants present in both ancestries in both the UKBB reference panel and the 1000 Genomes reference panel were retained. The sample size for EUR, *N* ^(1)^, was set to 200, 000, while the sample size for EAS, *N* ^(2)^, was set to either 20, 000 or 200, 000.

We considered three scenarios with varying degrees of heterogeneity in genetic architectures across two ancestries: (1) both ancestries have 5 shared causal variants; (2) both ancestries have 3 shared causal variants, and each ancestry has 1 specific causal variant; (3) both ancestries have 1 shared causal variant, and each ancestry has 2 specific causal variants. To sample causal variants, we first computed 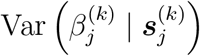 based on the linear model (14) using 52 functional annotations of variants from baseline model^40^ and a base annotation for *k* ∈ {1, 2}, where ***τ***^(*k*)^ are obtained from the g-LDSC estimate for schizophrenia GWAS in EUR and EAS ancestries^35^ (supplementary). Shared causal variants were then sampled with probabilities proportional to 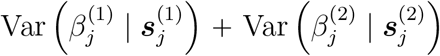, and ancestry-specific causal variants were sampled from the remaining variants with probabilities proportional to 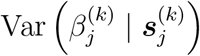 for *k* ∈ {1,2}. The effect sizes of shared causal variants were sampled independently from 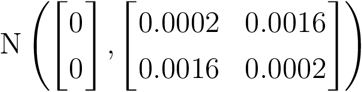, while the effect sizes of ancestry-specific causal variants were sampled independently from N(0, 0.0002). Finally, following the strategy of ref. ^46^, we simulated GWAS summary data for each block as

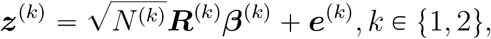

where ***e***^(*k*)^ is a noise term following N (**0, *R***^(*k*)^).

### Compared methods

For MACHINE, MACHINE + PolyFun, MACHINE + g-LDSC, h2-D2, h2-D2 + PolyFun, and h2-D2 + g-LDSC, the hyper-parameter *b* was set according to an empirical Bayesian approach with default settings (Supplementary Note). After determining *b*, we ran 5,500 iterations of MCMC, discarding the first 500 as burn-in, with parameters “thin = 1” and “stepsize = 1”. MCMC convergence was monitored using the potential scale reduction factor (PSRF)^19,47^. If the PSRF for any 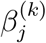 for *j* ∈ {1, …, *J*} and *k* ∈ 𝒦_*j*_ exceeded 1.2, the first 1,000 iterations were discarded and a further 1,000 iterations were run. This process was repeated until all PSRFs were less than 1.2. The CLs were computed for each ancestry separately. The CL for the *j*-th variant being a causal variant in at least one ancestry was computed as 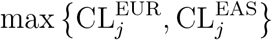, and the CL of the *j*-th variant being a shared causal variant was computed as 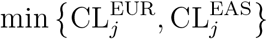. The 95% CSs were identified for each ancestry separately with settings “coverage = 0.95” and “purity = 0.5”, where the purity of a CS is the minimum absolute correlation between any pair of variants within the CS.

MESuSiE (v1.0) was run with default settings for all parameters, and the number of single effects L was set to 5 in simulations. The PIP for each variant being causal in at least one ancestry, ancestry-specific PIPs, the PIP being a shared causal variant, and the 95% CSs were obtained from the output. The PIP for the *j*-th variant being causal in the *k*-th ancestry is PIP^(*k*)-specific^ + PIP^shared^ for *k* ∈ {1, 2}. In real data analyses, only variants present in all ancestries were retained for MESuSiE fine mapping.

SuSiEx (v1.1.2) was run with options “keep-ambig = True”, “maf = 0.0001”, and “pval_thresh = 1”. The PIPs of all variants in each CS and the probability of each CS being causal in each population were obtained from the output files. The probability threshold 0.8 was used to infer whether a fine-mapped signal is causal in each population. If a CS was identified as causal in both populations, it was recognized as a shared CS.

XMAP (v1.0) was run with “K = 5”, “Omega = diag(1e-100,2)”, and “Sig_E = c(1,1)” in simulations. The “get_CS” function was used to obtain 95% CSs for each ancestry with “coverage = 0.95” and “min_abs_corr = 0.5”. The ancestry-specificity of each CS is determined by the purity filter. If the purity of a CS in an ancestry exceeded 0.5, the CS was inferred to be causal in that ancestry.

MultiSuSiE (v1.0) was run with ‘scale_prior_variance = 0.2”, “min_abs_corr = 0.5”, “L = 5”, and “low_memory_mode = False”. Since MultiSuSiE does not provide a criterion for ancestry specificity of CSs, every identified CS was inferred to be causal in all ancestries.

For both XMAP and MultiSuSiE, the output PIPs were used to evaluate whether each variant is causal in at least one ancestry, in a specific ancestry, or is shared by both ancestries.

SuSiE (v0.14.2) was run with “var_y = 1”, “estimate_residual_variance = TRUE”, and “refine = TRUE”. In simulations, we set “L = 5”.

RSparsePro was run with “minldthres 0.5”. In simulations, we set “K 5”.

CARMA (v1.0) was run with “lambda.list = list(1)” and “rho.index = 0.95”. We set “outlier.switch = FALSE” for simulations with in-sample LD matrices and “outlier.switch = TRUE” for simulations with out-of-sample LD matrices. Only 95% CSs with purities ⩾ 0.5 were retained.

For SuSiE, RSparsePro, and CARMA, the PIP of the *j*-th variant being causal in at least one ancestry was computed as 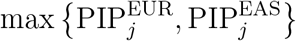, and the PIP for being a shared causal variant as 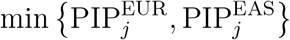.

For MESuSiE, SuSiE, and RSparsePro in real data analyses, we set the number of single effects L (or K for RSparsePro) as 5 initially. If the number of identified 95% CSs was equal to L, we increased L by 5 and repeated fine mapping, continuing until the number of identified 95% CSs was less than L or L reached 20.

For MACHINE, MACHINE + PolyFun, MACHINE + g-LDSC, h2-D2, h2-D2 + PolyFun, h2-D2 + g-LDSC, SuSiE, RSparsePro, and CARMA, each 95% CS in each ancestry served as a cross-ancestry 95% CS. If two 95% CSs from different ancestries had common variants, their union was inferred as a CS for shared causal variants.

### Lipid trait summary data preprocessing

UKBB summary statistics for four lipit traits were downloaded from Pan-UKBB ^25^. Meta-analysis summary statistics for four lipit traits were obtained from GLGC ^23^. The sample sizes in the GLGC summary statistics vary across variants. For each trait and each ancestry, we filtered out variants with sample sizes less than 90% of the maximum sample size across all variants, and scaled the *z*-scores as described above. Only variants present in both datasets and the UKBB reference panel were retained.

### SCZ summary data preprocessing

SCZ summary statistics were obtained from the Psychiatric Genomics Consortium ^35^. For each ancestry, variants with sample sizes equal to the maximum sample sizes, high imputation quality (imputation INFO > 0.8), and were present in the UKBB reference panel were retained.

### Statistics and reproducibility

Barplots were used to demonstrate the results in some figures. The length of the error bar represents the standard error. The standard error for a binary variable’s proportion is calculated as 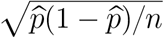, where 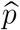 is the sample proportion and *n* is the sample size.

## Data availability

The UK Biobank data are accessed under Application Number 140822. 1000 Genomes data (phase 3) are available at http://ftp.1000genomes.ebi.ac.uk/. Meta-analysis summary data for the four lipid traits are available at https://csg.sph.umich.edu/willer/public/glgc-lipids2021/. UKBB summary data for the four lipid traits are available at https://pan.ukbb.broadinstitute.org. The bed files of 52 functional annotations from the baseline model v1.1 are available at https://console.cloud.google.com/storage/browser/broad-alkesgroup-public-requester-pays/LDSCORE/baseline_v1.1_bedfiles.tgz. The complete contents of dbSNP database (build 151) with GRCh37.p13 as reference assembly are available at https://ftp.ncbi.nih.gov/snp/organisms/human_9606_b151_GRCh37p13/. SuSiE fine-mapped *cis*-eQTL data from GTEx V10 database are available at https://storage.googleapis.com/adult-gtex/bulk-qtl/v10/susie-qtl/GTEx_v10_SuSiE_eQTL.tar. Schizophrenia GWAS summary statistics are available at https://figshare.com/articles/dataset/scz2022/19426775.

## Code availability

MACHINE (v1.0) is available at https://github.com/xiangli428/MACHINE. h2-D2 (v2.0) is available at https://github.com/xiangli428/h2D2. g-LDSC (v1.0.0) is available at https://github.com/xzw20046/gldsc. PolyFun is available at https://github.com/omerwe/polyfun. MESuSiE (v1.0) is available at https://github.com/borangao/MESuSiE. SuSiEx (v1.1.2) is available at https://github.com/getian107/SuSiEx. XMAP (v1.0) is available at https://github.com/YangLabHKUST/XMAP. MultiSuSiE (v1.0) is available at https://github.com/jordanero/MultiSuSiE. SuSiE (v0.14.2) is available at https://github.com/stephenslab/susieR. RSparsePro is available at https://github.com/zhwm/RSparsePro_LD. CARMA (v1.0) is available at https://github.com/Iuliana-Ionita-Laza/CARMA. PLINK (v1.9) is available at https://www.cog-genomics.org/plink/. PLINK (v2.0) is available at https://www.cog-genomics.org/plink/2.0/. BedTools (v2.31.0) is available at https://bedtools.readthedocs.io/en/latest/.

## Author contributions

X.L. and Y.D.Z. designed the method. X.L. developed the software package and under-took the simulation studies and real data analyses with the assistance of Z.X., X.L. wrote the original draft of the manuscript. Z.X., P.C.S. and Y.D.Z. provided critical feedback during the study and helped revise the manuscript.

---

**Extended Data Fig. 1:**
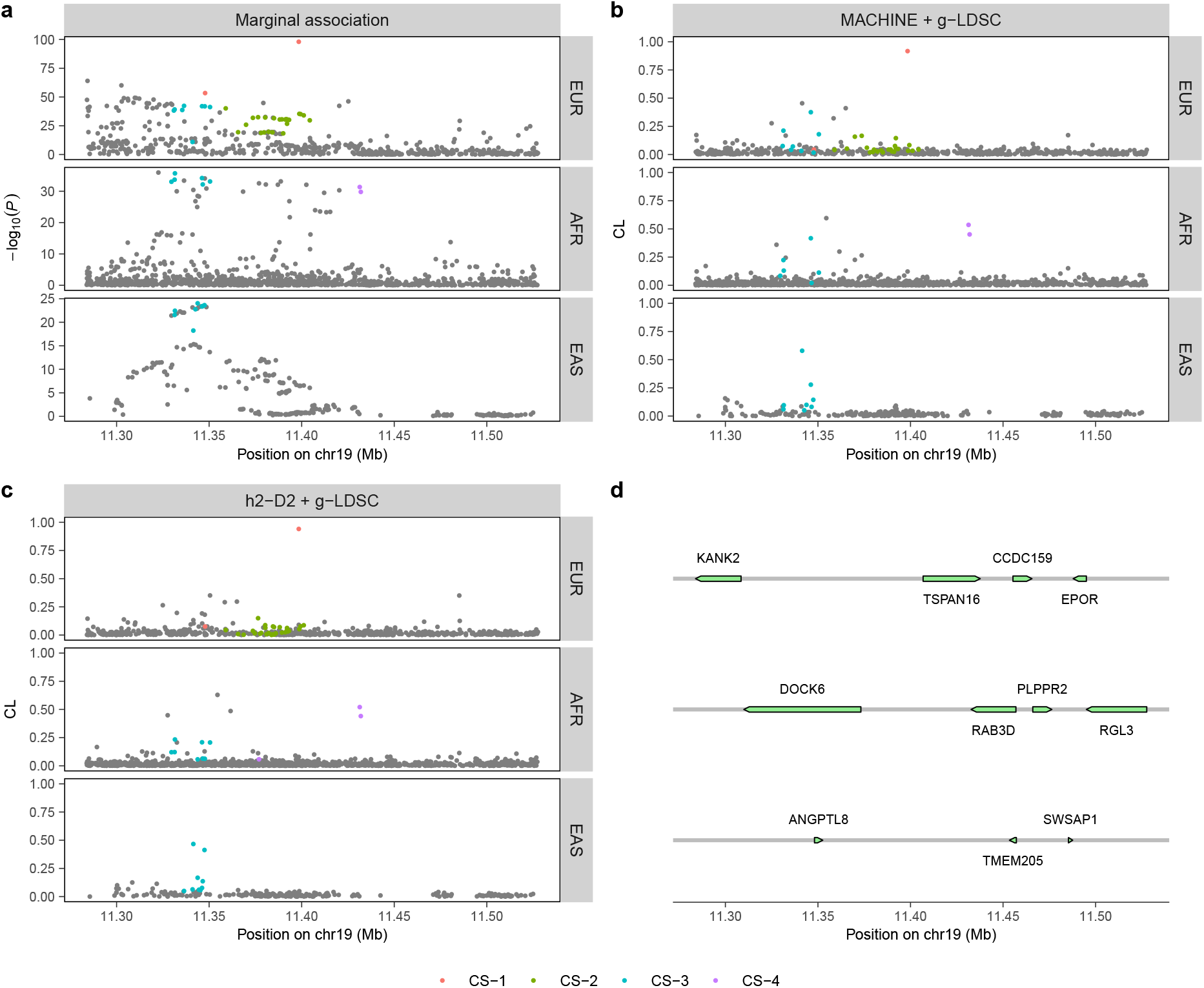
LocusZoom plot of fine-mapping results of LDL for chr19:11,283,869-11,527,628. Each color represents a 95% CS. **a**, Two-sided − log_10_(*P* value) in GWAS. 95% CSs identified by MACHINE + g-LDSC are colored. **b**, CLs of variants obtained by MACHINE + g-LDSC. **c**, CLs of variants obtained by h2-D2 + g-LDSC. **d**, Gene positions within the locus.

**Extended Data Fig. 2:**
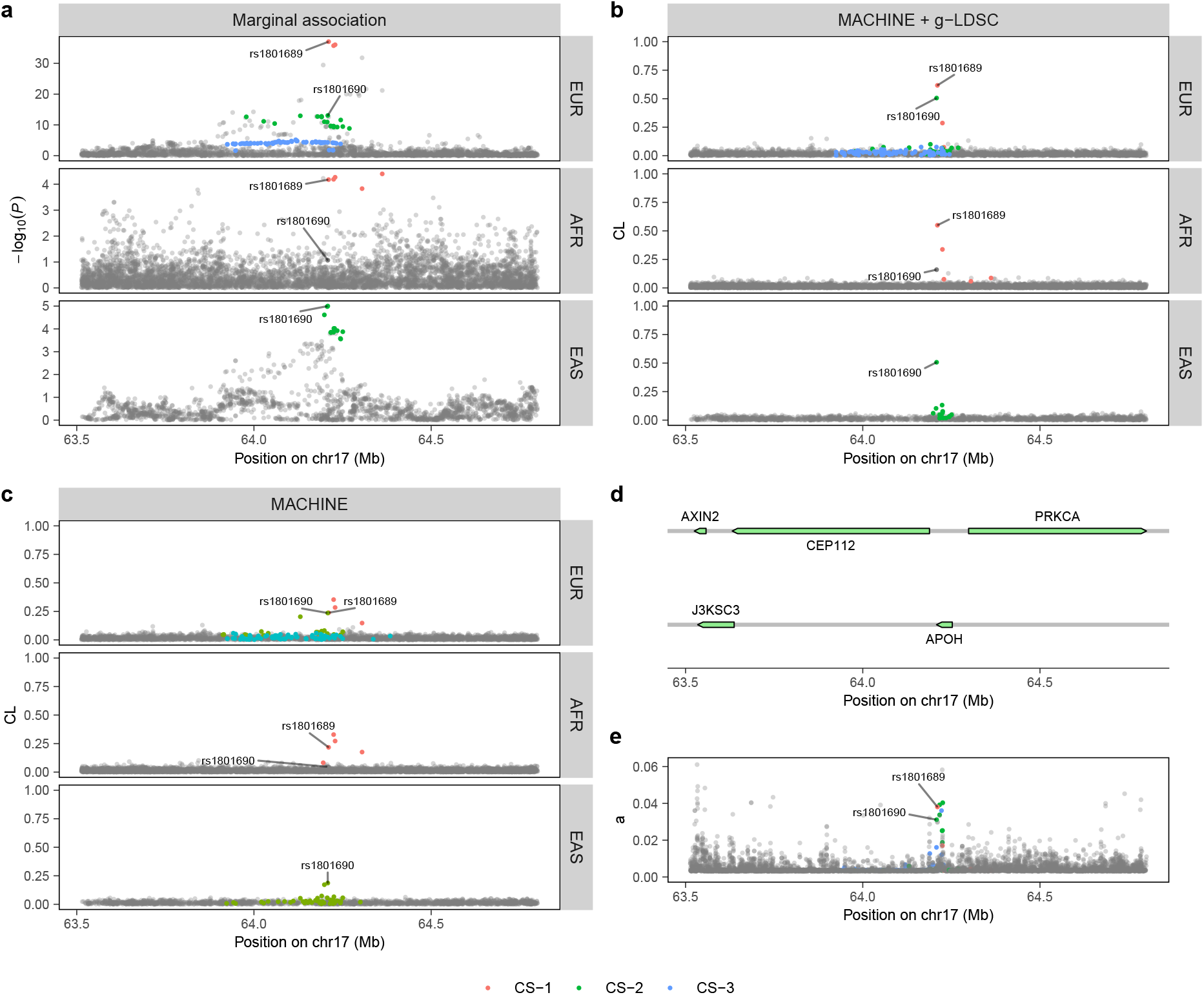
LocusZoom plot of fine-mapping results for TG in chr17:63,513,466-64,800,552. Each color represents a 95% CS. **a**, Two-sided −log_10_(*P* value) in GWAS. 95% CSs identified by MACHINE + g-LDSC are colored. **b**, CLs of variants obtained by MACHINE + g-LDSC. **c**, CLs of variants obtained by MACHINE. **d**, Gene positions within the locus. **e**, Hyper-parameters ***a*** of MACHINE + g-LDSC for all variants.

**Extended Data Fig. 3:**
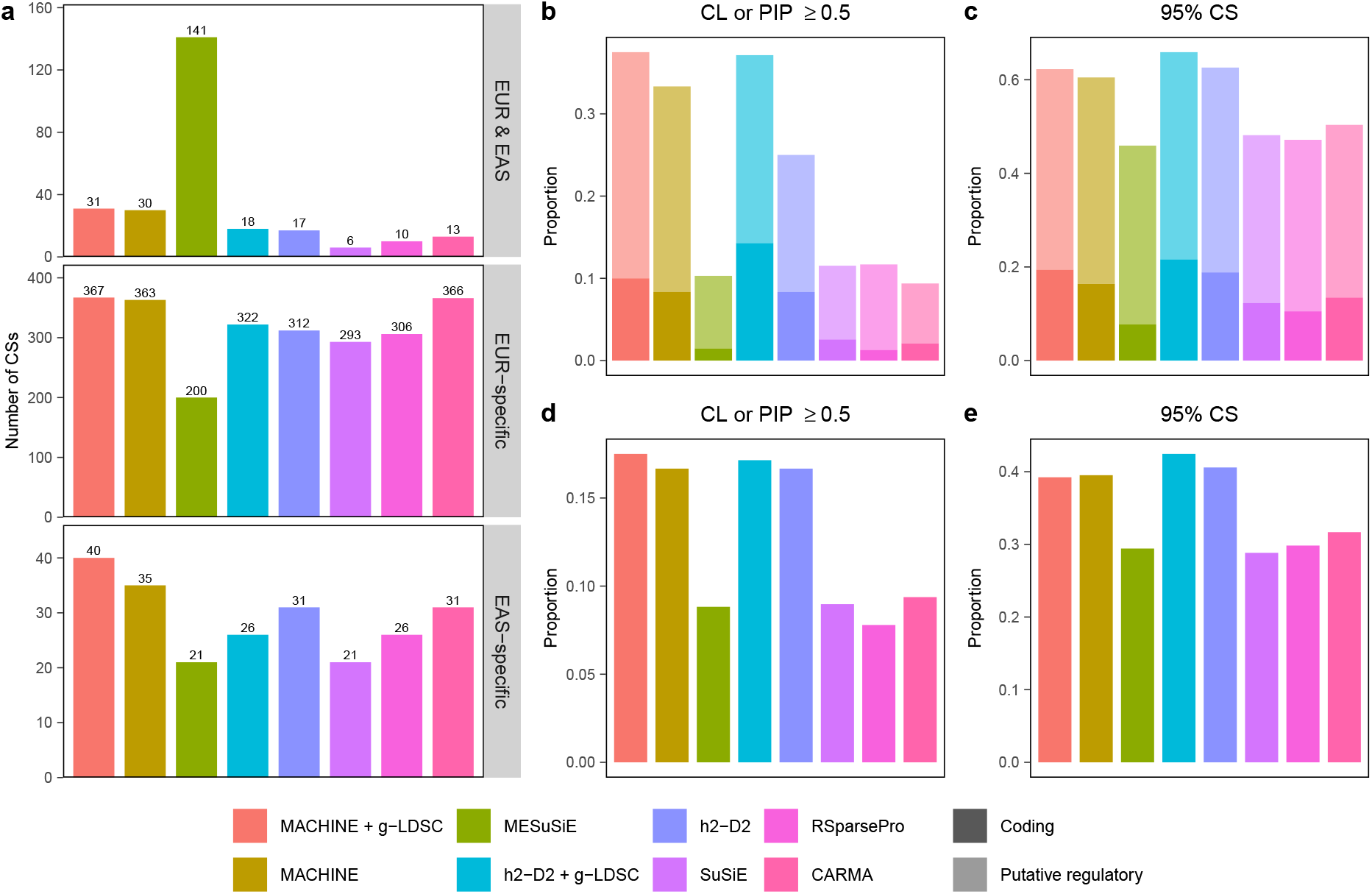
Comparison of fine-mapping methods in real data analysis of SCZ. **a**, Number of shared, EUR-specific, and EAS-specific 95% CSs identified by each method. **b**, Proportion of coding and putative regulatory variants among variants with CL or PIP ⩾ 0.5. **c**, Proportion of 95% CSs containing at least one coding variant, and proportion of 95% CSs containing at least one putative regulatory variant but no coding variant. **d**, Proportion of fine-mapped eQTLs among variants with CL or PIP ⩾ 0.5. **e**, Proportion of 95% CSs containing at least one fine-mapped eQTL. For **b**-**e**, results are aggregated across ancestries. Numerical results are available in Supplementary Tables 14-15.

