## Supplementary Information for "MACHINE: a robust and scalable multi-ancestry fine-mapping method using a continuous global-local shrinkage prior"

### Supplementary Figures

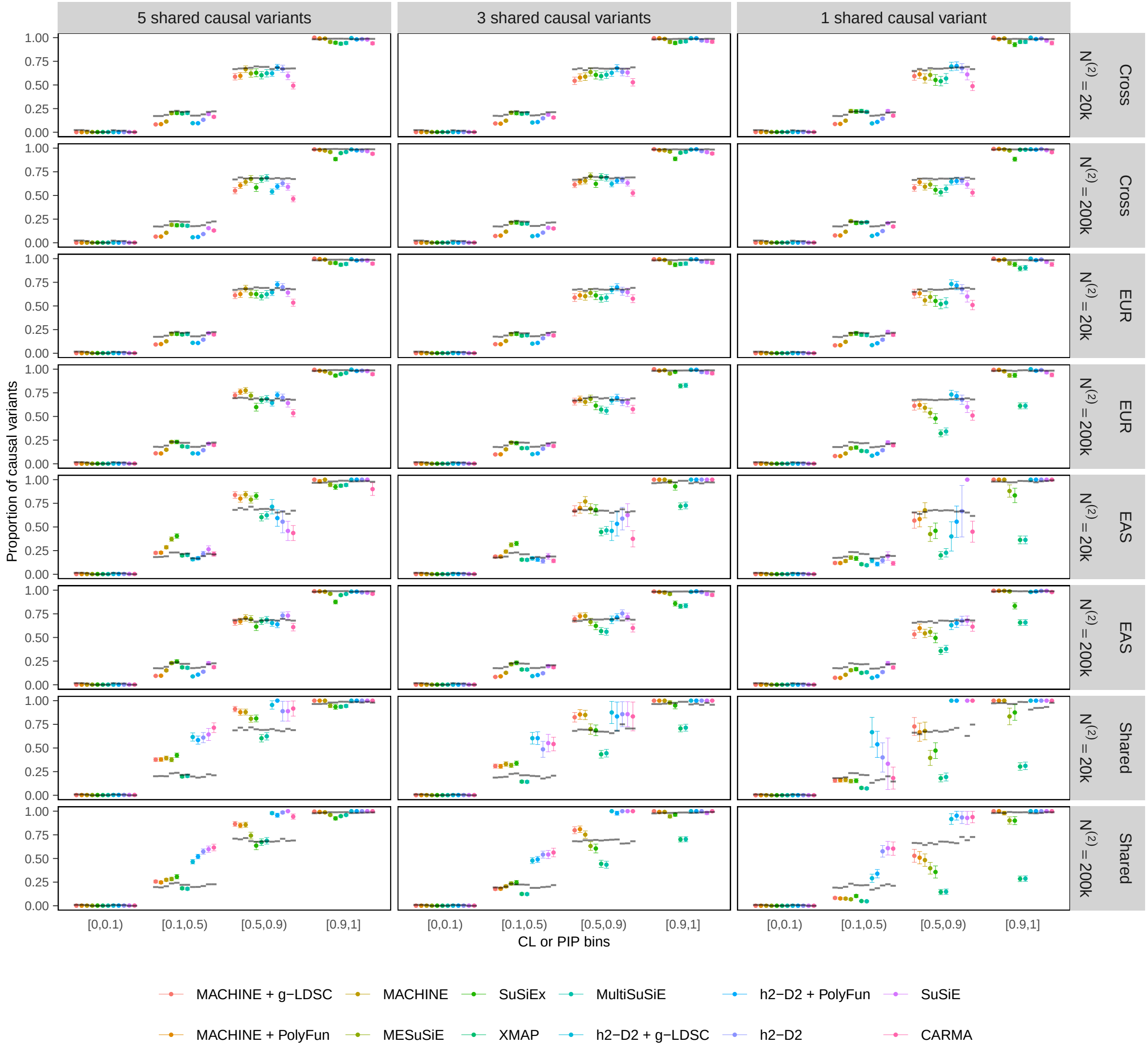


**Supplementary Fig. 1: Calibration plot of fine-mapping methods when in-sample LD matrices are used.** All values are computed by aggregating 200 simulated datasets. Colored dots represent the true proportion of causal variants among variants within each CL or PIP bin, with error bars indicating standard errors. Horizontal segments indicate the expected proportion of causal variants (i.e. the average CL or PIP within each bin). Numerical results are available in Supplementary Table 3.

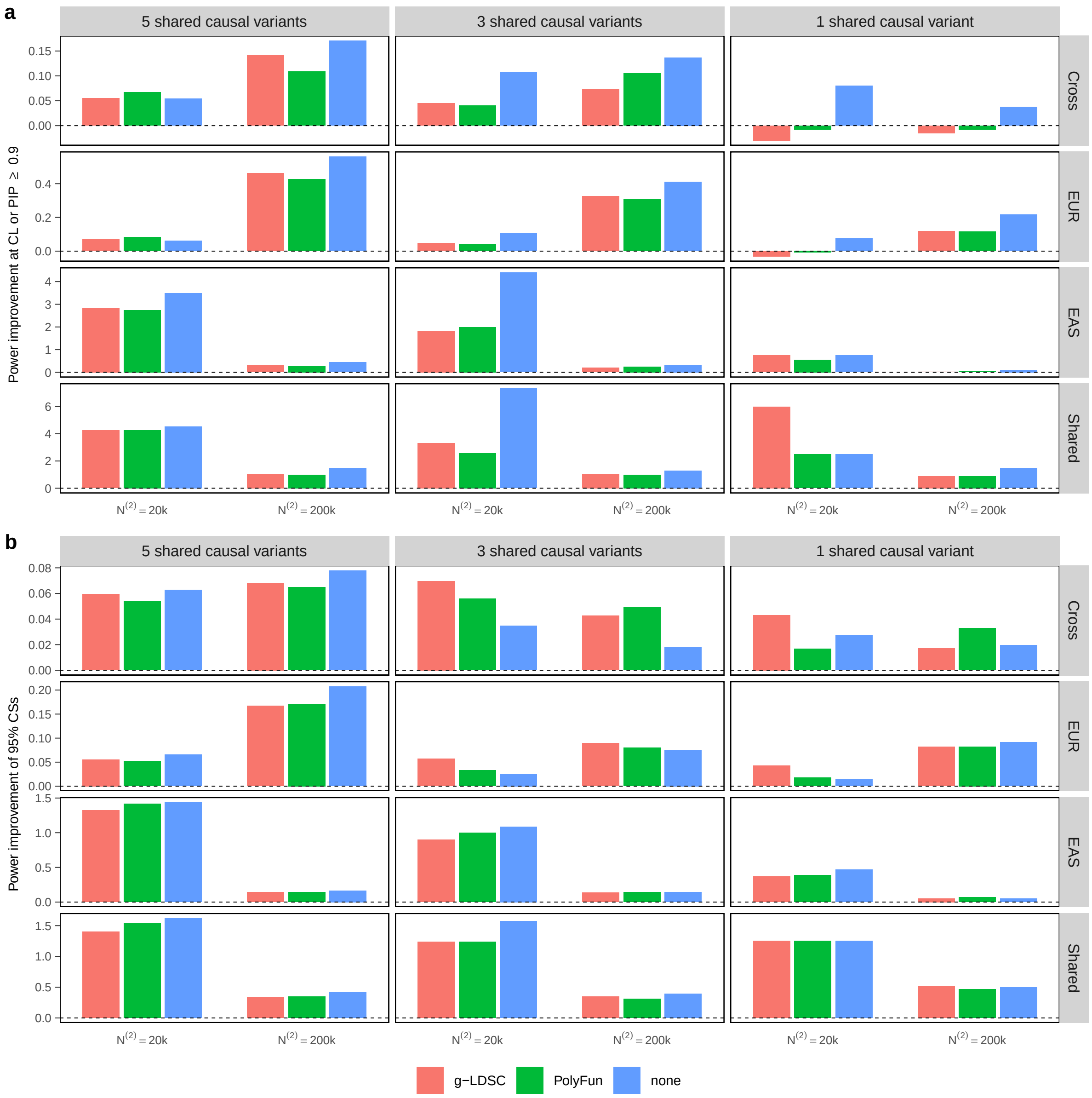


**Supplementary Fig. 2: Improvement in power of MACHINE compared to h2-D2 when in-sample LD matrices are used.** Power improvement is defined as $(\text{Power}^{(\text{MACHINE})} - \text{Power}^{(\text{h2-D2})})/\text{Power}^{(\text{h2-D2})}$. Numerical results are available in Supplementary Table 4.

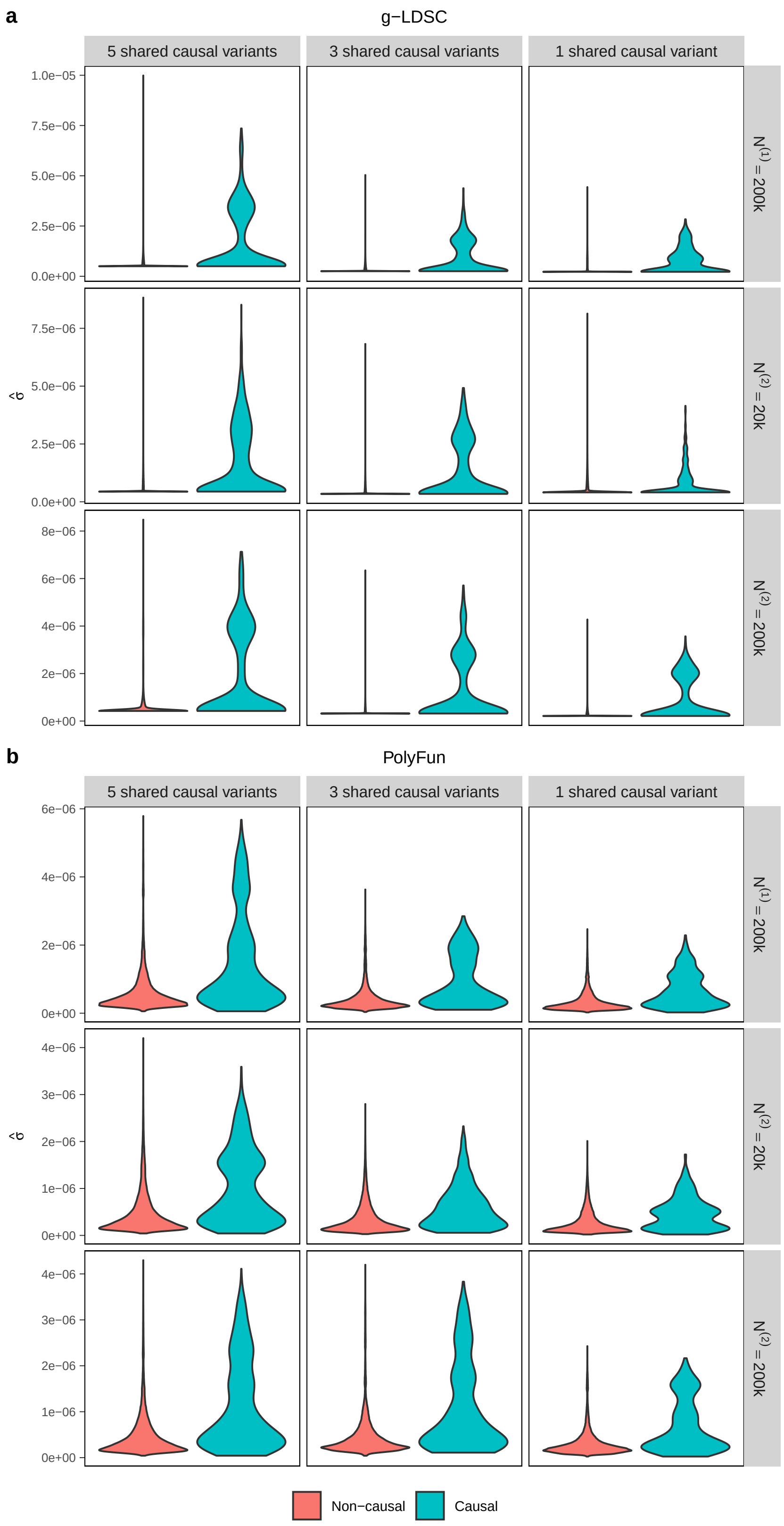


**Supplementary Fig. 3: Violin plots displaying the distribution of per-variant heritabilities estimated by (a) g-LDSC and (b) PolyFun for underlying causal and non-causal variants when in-sample LD matrices are used.** For each simulation setting, results are aggregated across 200 simulated datasets.

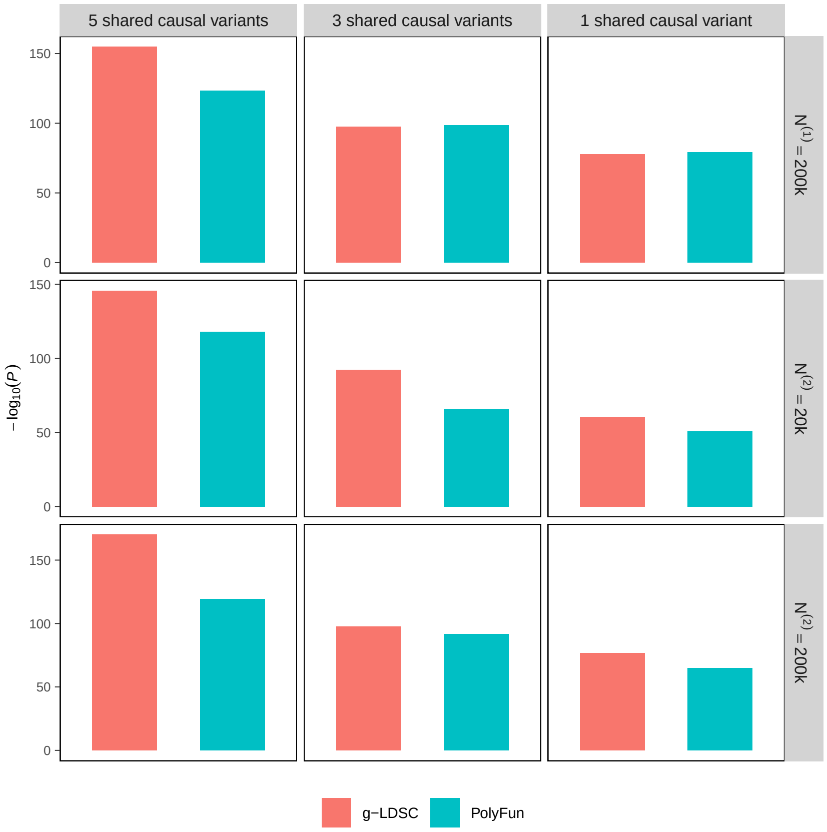


**Supplementary Fig. 4: Negative $\log_{10}$($P$ value) of Wilcoxon-Mann-Whitney tests comparing estimated per-variant heritabilities between causal and non-causal variants when in-sample LD matrices are used.** For each simulation setting, results are aggregated across 200 simulated datasets. Numerical results are available in Supplementary Table 5.

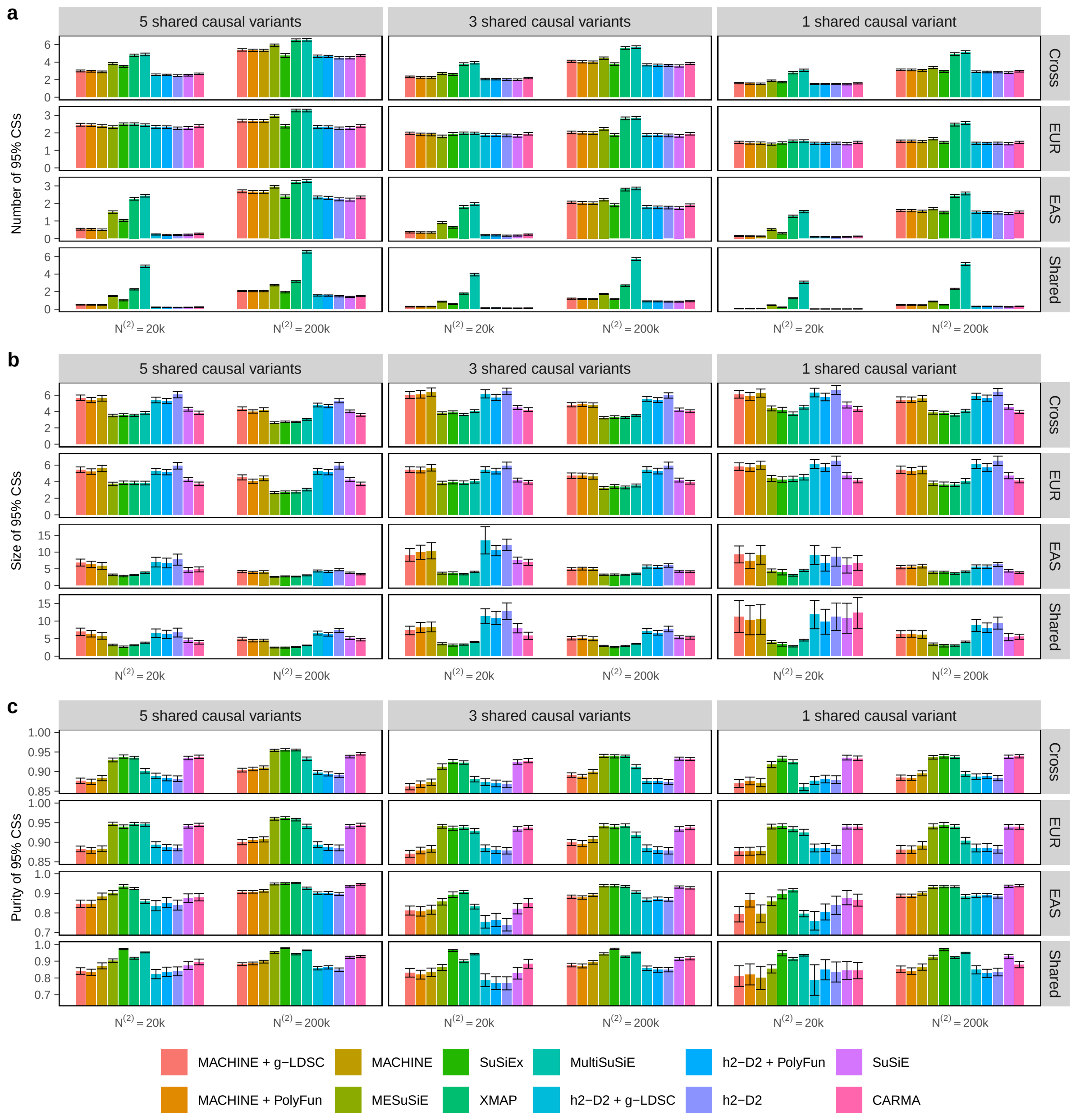


**Supplementary Fig. 5: Comparison of 95% CSs obtained by different fine-mapping methods when in-sample LD matrices are used.** All values are aggregated across 200 simulated datasets, with error bars indicating standard errors. **a,** Number of identified 95% CSs. **b,** Size of 95% CS, defined as the number of variants in each CS. **c,** Purity of 95% CS, defined as the minimum absolute correlation between any pair of variants within the CS. Numerical results are available in Supplementary Table 2.

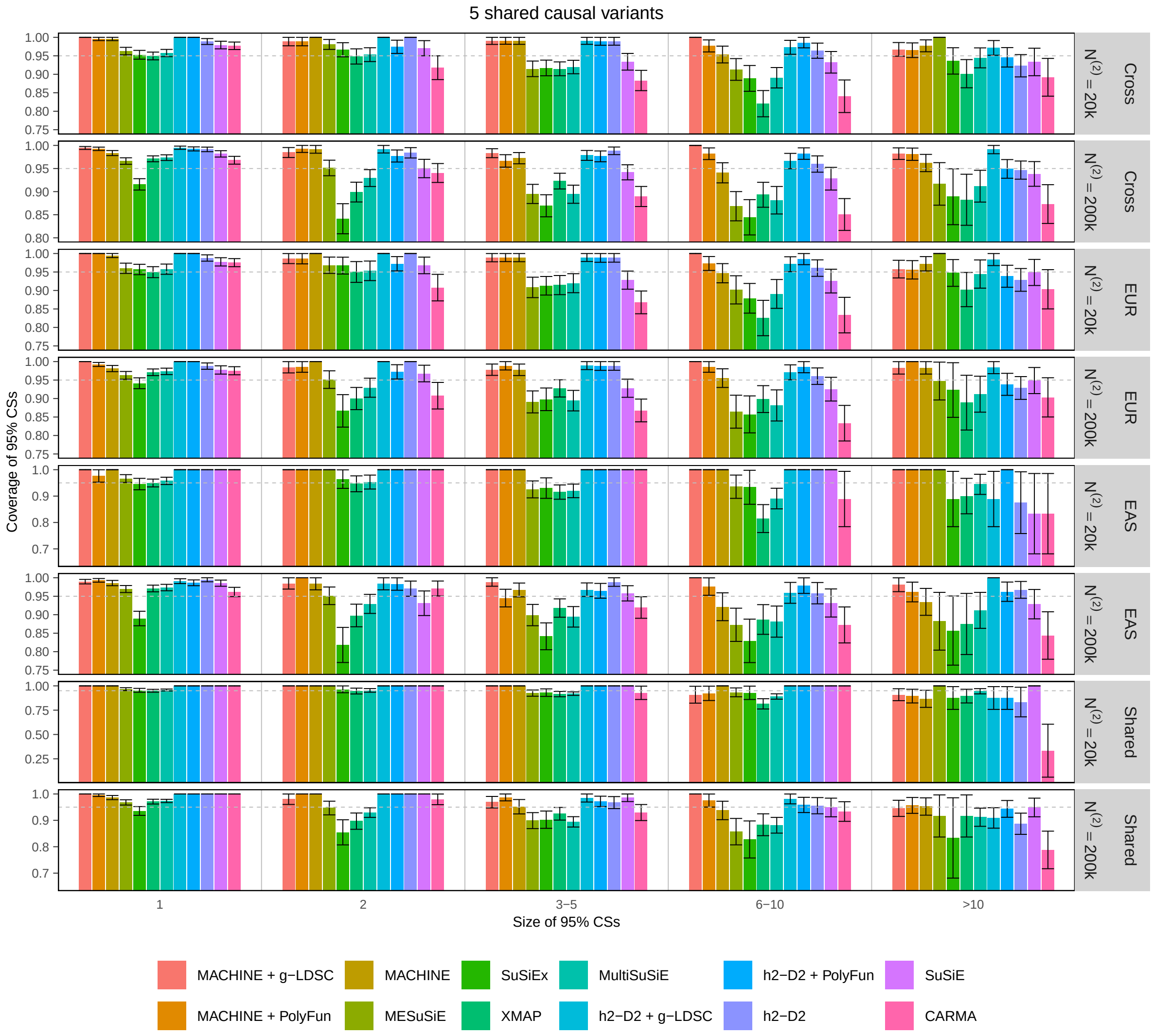


**Supplementary Fig. 6: Coverage of 95% CSs grouped by the CS size for scenarios with 5 shared causal variants per region when in-sample LD matrices are used.** For each method, 95% CSs from 200 simulated datasets are aggregated and grouped by their sizes. The proportion of 95% CSs containing at least one causal variant within each group is shown, with error bars indicating standard errors. Numerical results are available in Supplementary Table 6.

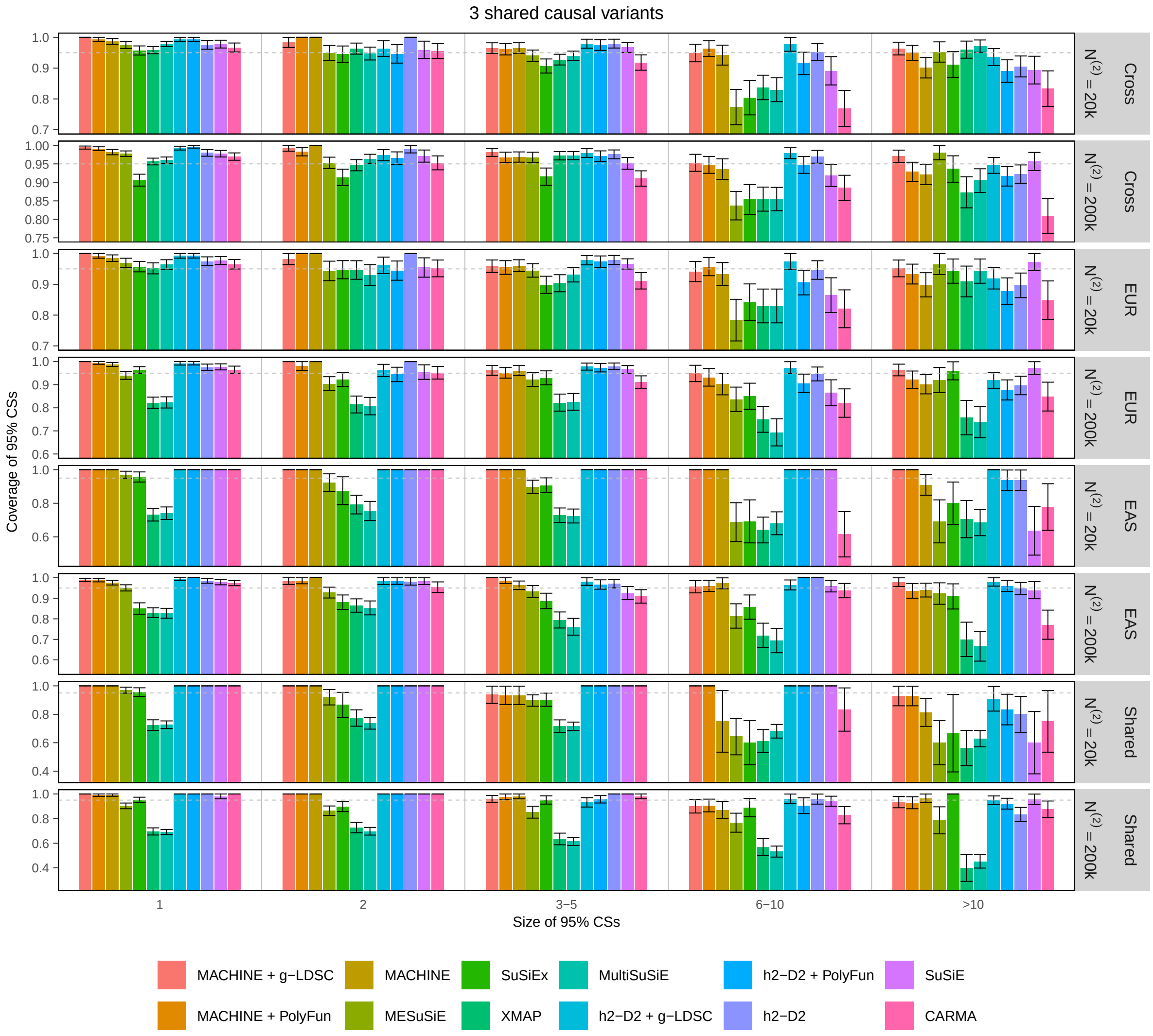


**Supplementary Fig. 7: Coverage of 95% CSs grouped by the CS size for scenarios with 3 shared causal variants per region when in-sample LD matrices are used.** For each method, 95% CSs from 200 simulated datasets are aggregated and grouped by their sizes. The proportion of 95% CSs containing at least one causal variant within each group is shown, with error bars indicating standard errors. Numerical results are available in Supplementary Table 6.

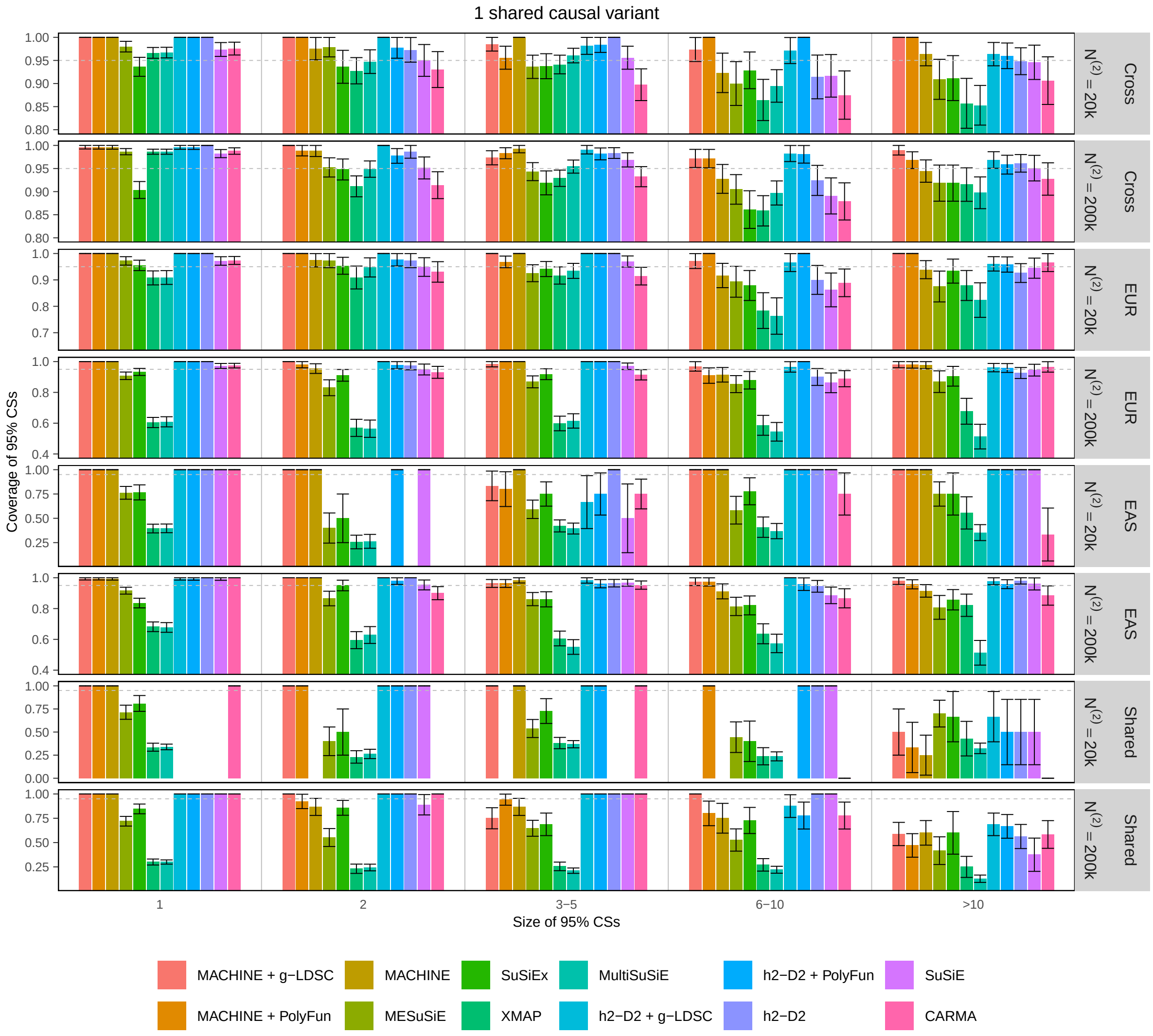


**Supplementary Fig. 8: Coverage of 95% CSs grouped by the CS size for scenarios with 1 shared causal variant per region when in-sample LD matrices are used.** For each method, 95% CSs from 200 simulated datasets are aggregated and grouped by their sizes. The proportion of 95% CSs containing at least one causal variant within each group is shown, with error bars indicating standard errors. Numerical results are available in Supplementary Table 6.

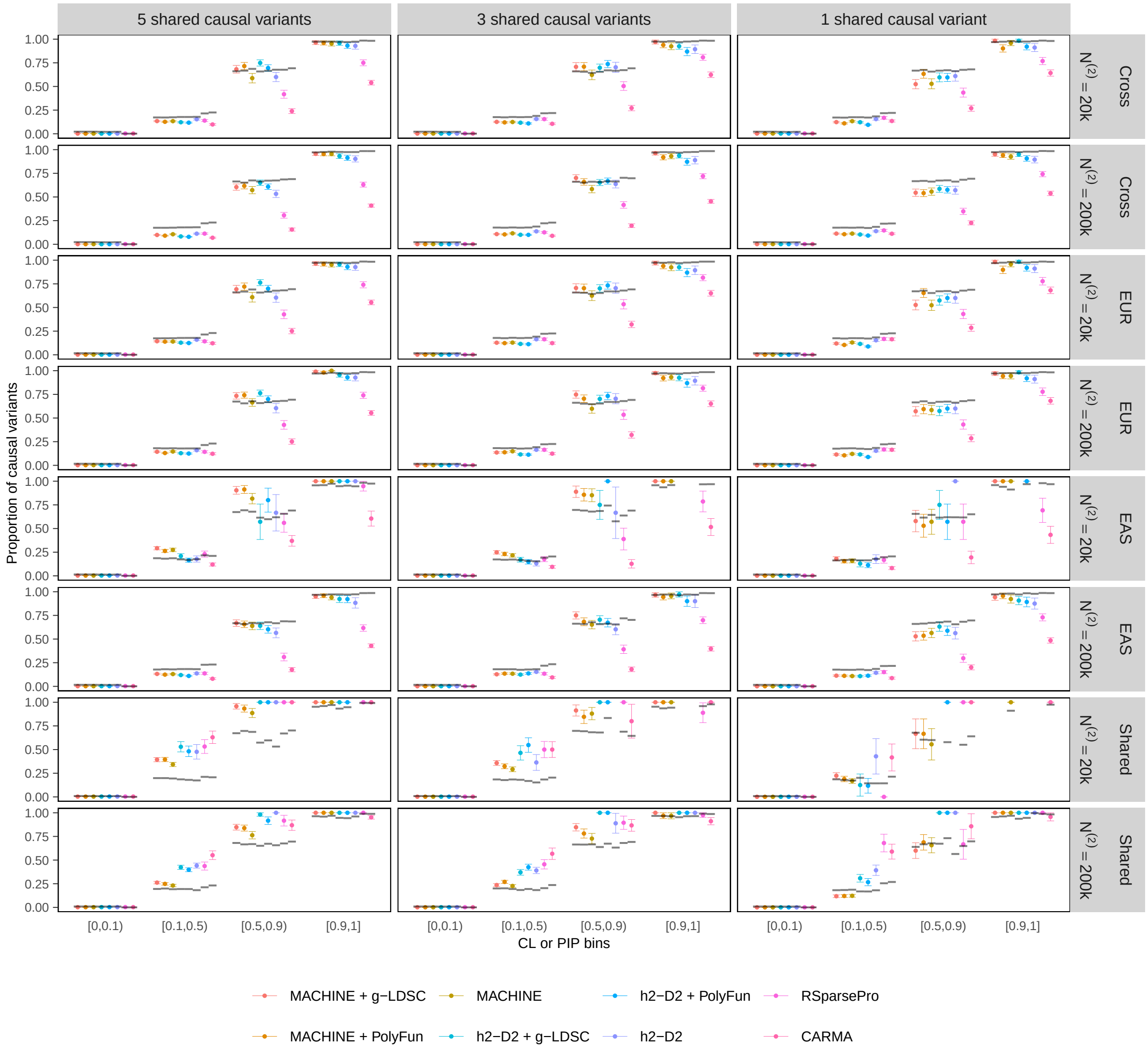


**Supplementary Fig. 9: Calibration plot of fine-mapping methods when out-of-sample LD matrices are used.** All values are computed by aggregating 200 simulated datasets. Colored dots represent the true proportion of causal variants among variants within each CL or PIP bin, with error bars indicating standard errors. Horizontal segments indicate the expected proportion of causal variants (i.e. the average CL or PIP within each bin). Numerical results are available in Supplementary Table 3.

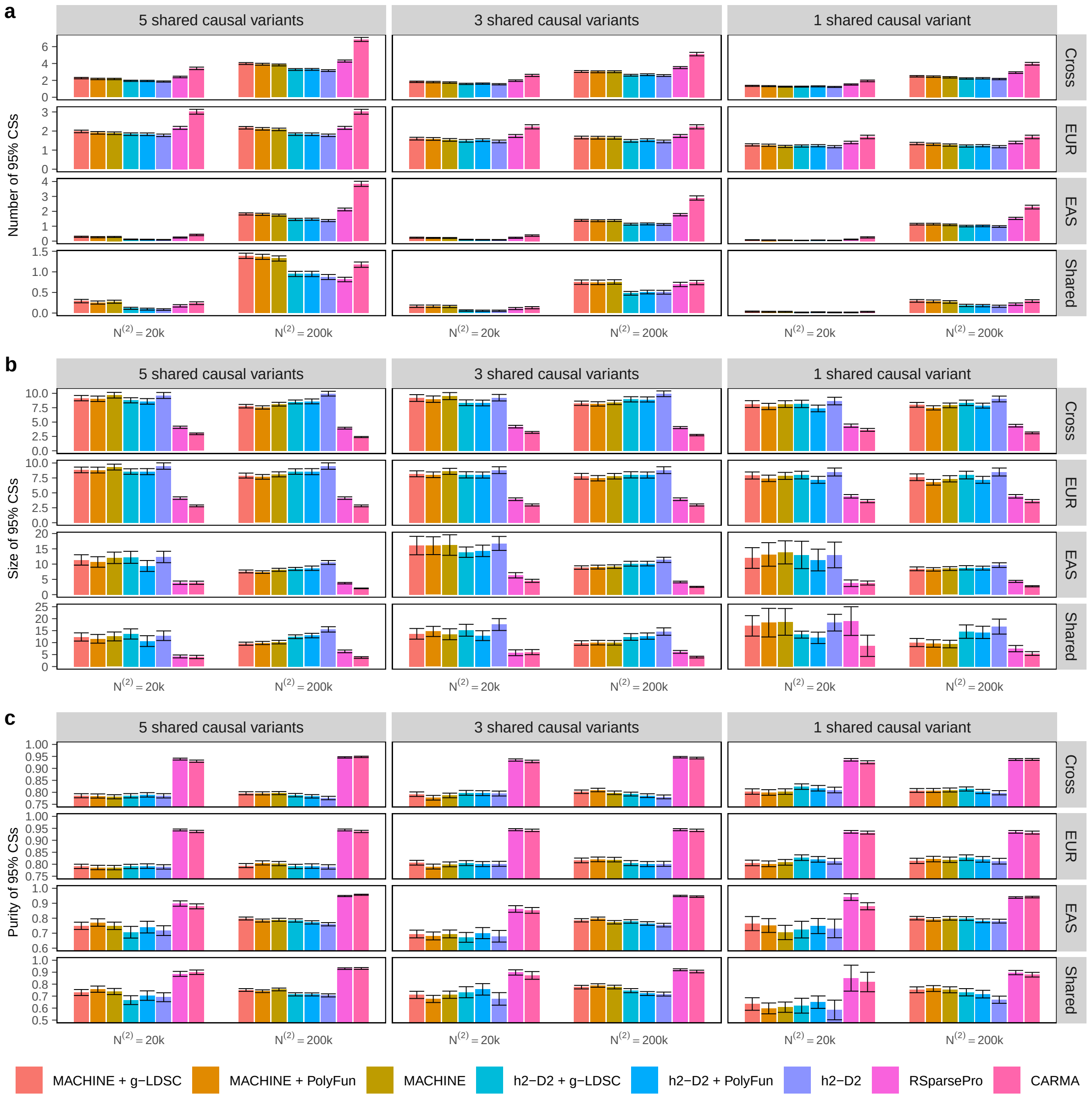


**Supplementary Fig. 10: Comparison of 95% CSs obtained by different fine-mapping methods when out-of-sample LD matrices are used.** All values are aggregated across 200 simulated datasets, with error bars indicating standard errors. **a,** Number of identified 95% CSs. **b,** Size of 95% CS, defined as the number of variants in each CS. **c,** Purity of 95% CS, defined as the minimum absolute correlation between any pair of variants within the CS. Numerical results are available in Supplementary Table 2.

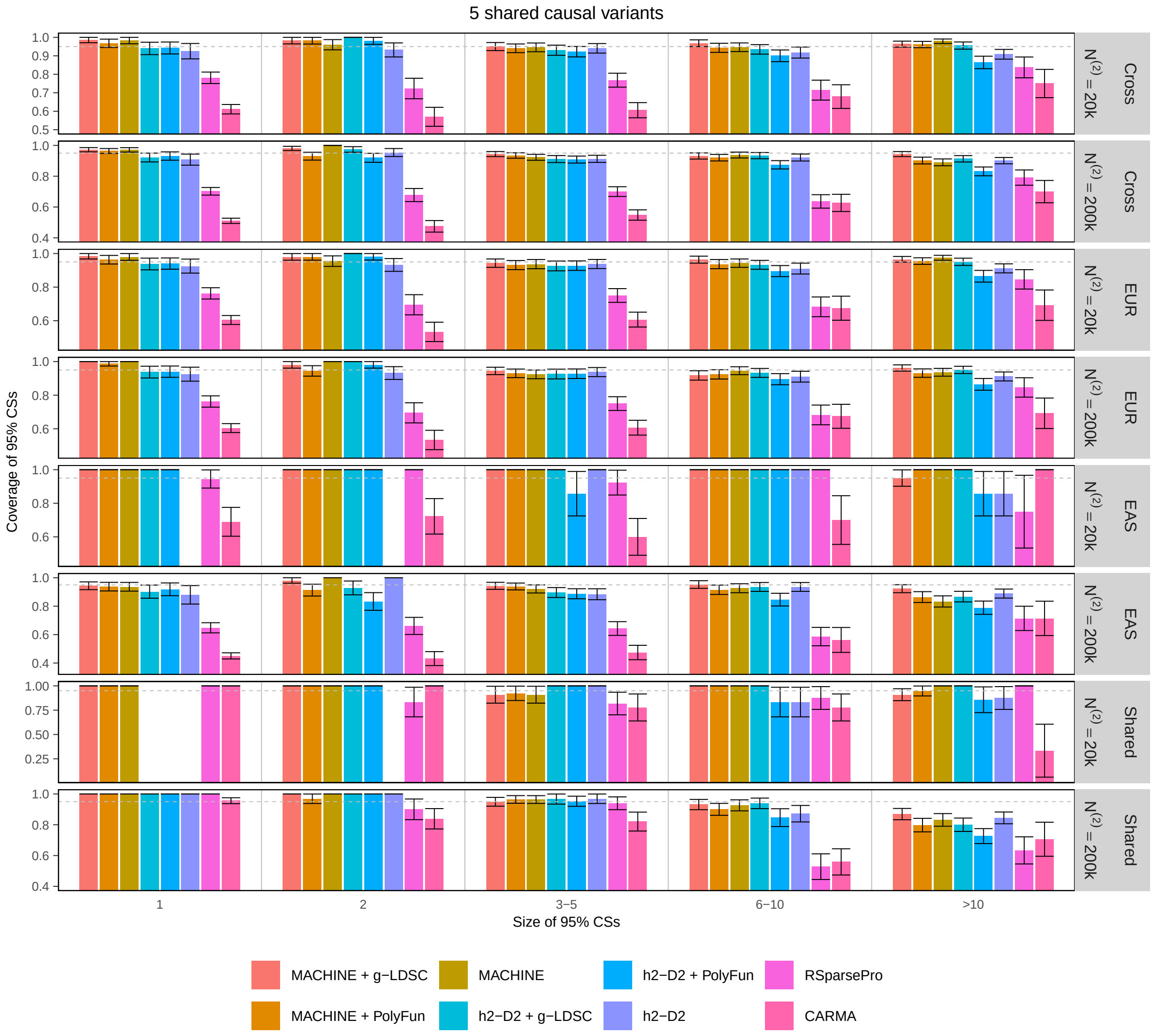


**Supplementary Fig. 11: Coverage of 95% CSs grouped by the CS size for scenarios with 5 shared causal variants per region when out-of-sample LD matrices are used.** For each method, 95% CSs from 200 simulated datasets are aggregated and grouped by their sizes. The proportion of 95% CSs containing at least one causal variant within each group is shown, with error bars indicating standard errors. Numerical results are available in Supplementary Table 6.

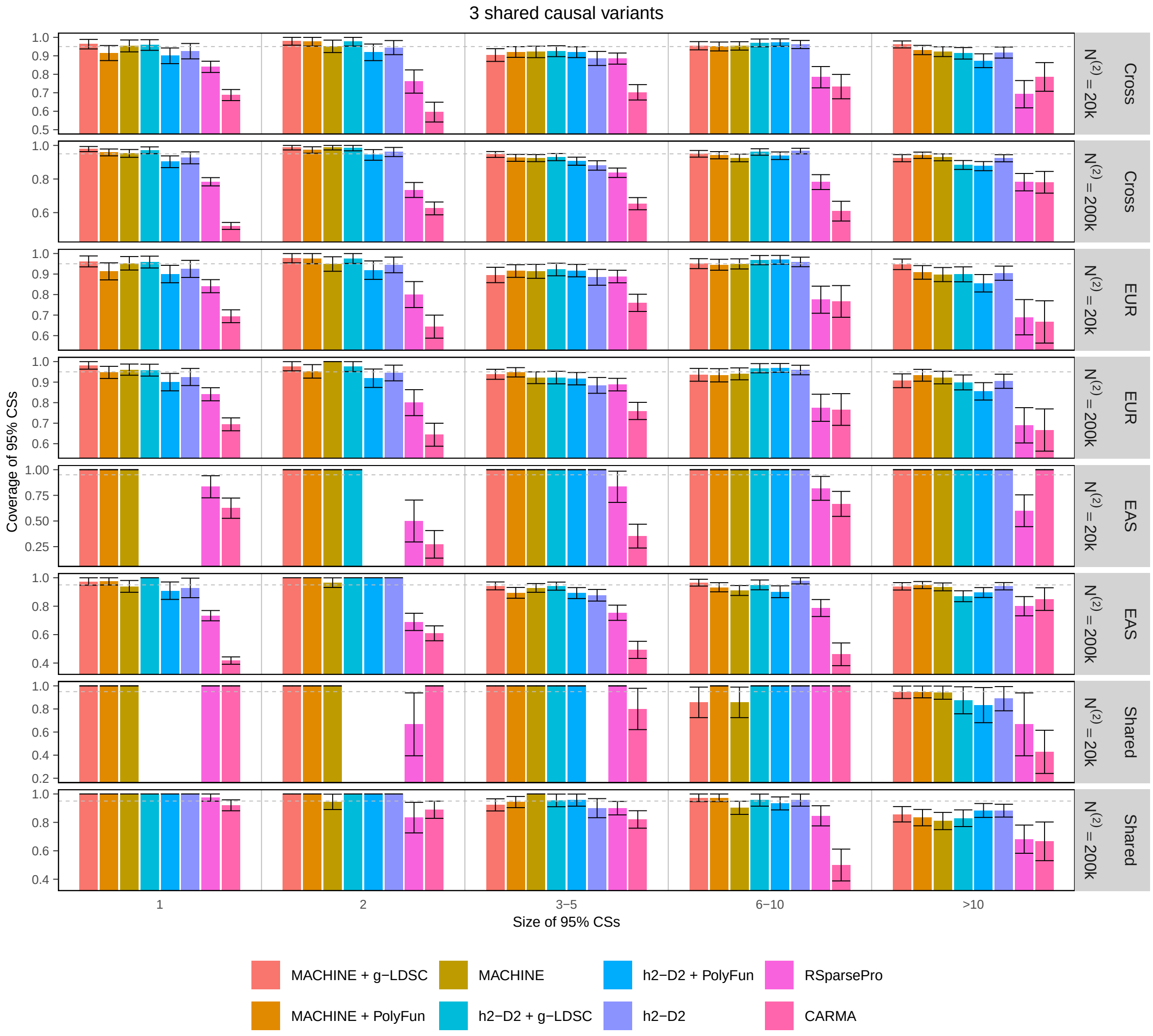


**Supplementary Fig. 12: Coverage of 95% CSs grouped by the CS size for scenarios with 3 shared causal variants per region when out-of-sample LD matrices are used.** For each method, 95% CSs from 200 simulated datasets are aggregated and grouped by their sizes. The proportion of 95% CSs containing at least one causal variant within each group is shown, with error bars indicating standard errors. Numerical results are available in Supplementary Table 6.

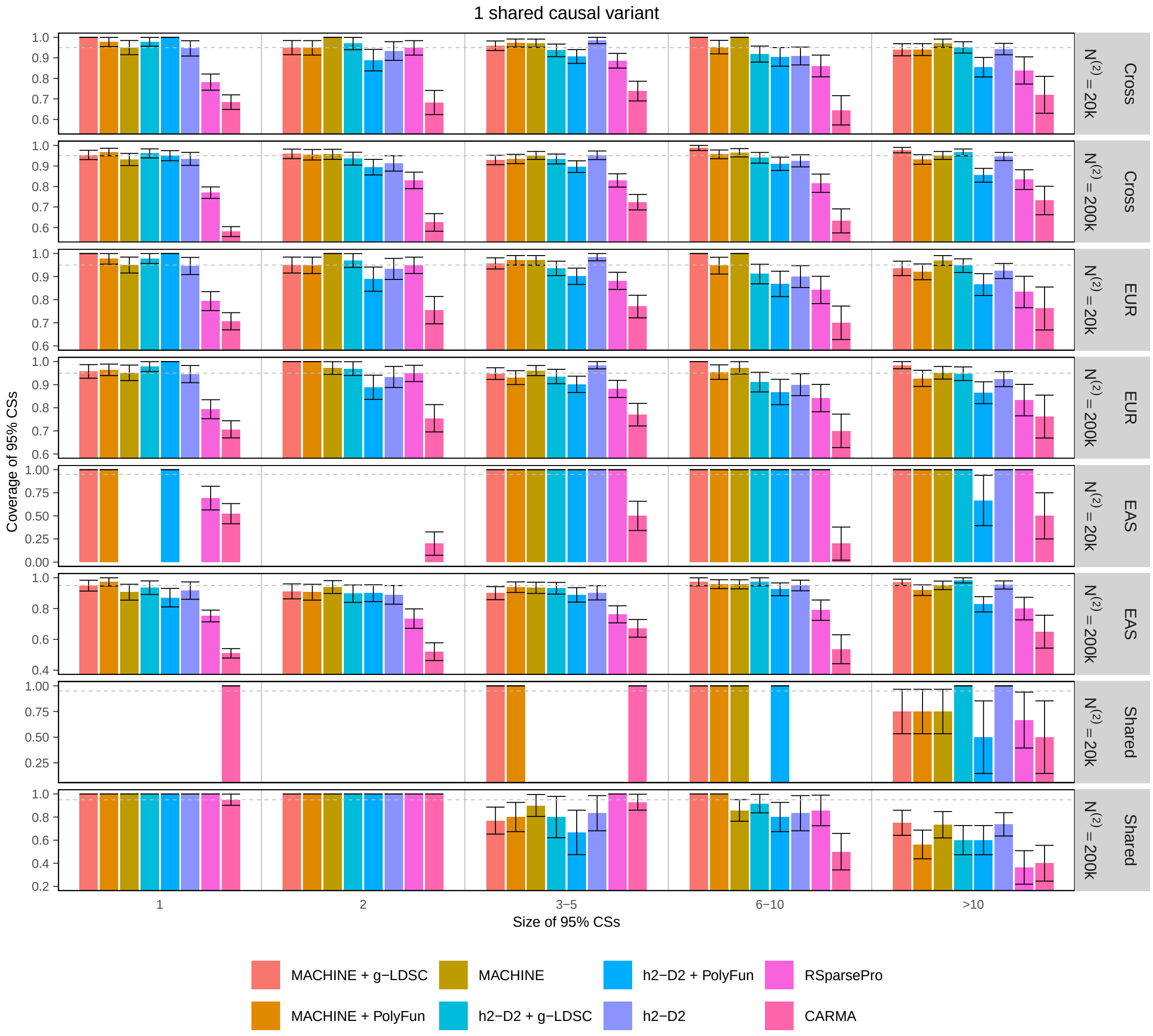


**Supplementary Fig. 13: Coverage of 95% CSs grouped by the CS size for scenarios with 1 shared causal variant per region when out-of-sample LD matrices are used.** For each method, 95% CSs from 200 simulated datasets are aggregated and grouped by their sizes. The proportion of 95% CSs containing at least one causal variant within each group is shown, with error bars indicating standard errors. Numerical results are available in Supplementary Table 6.

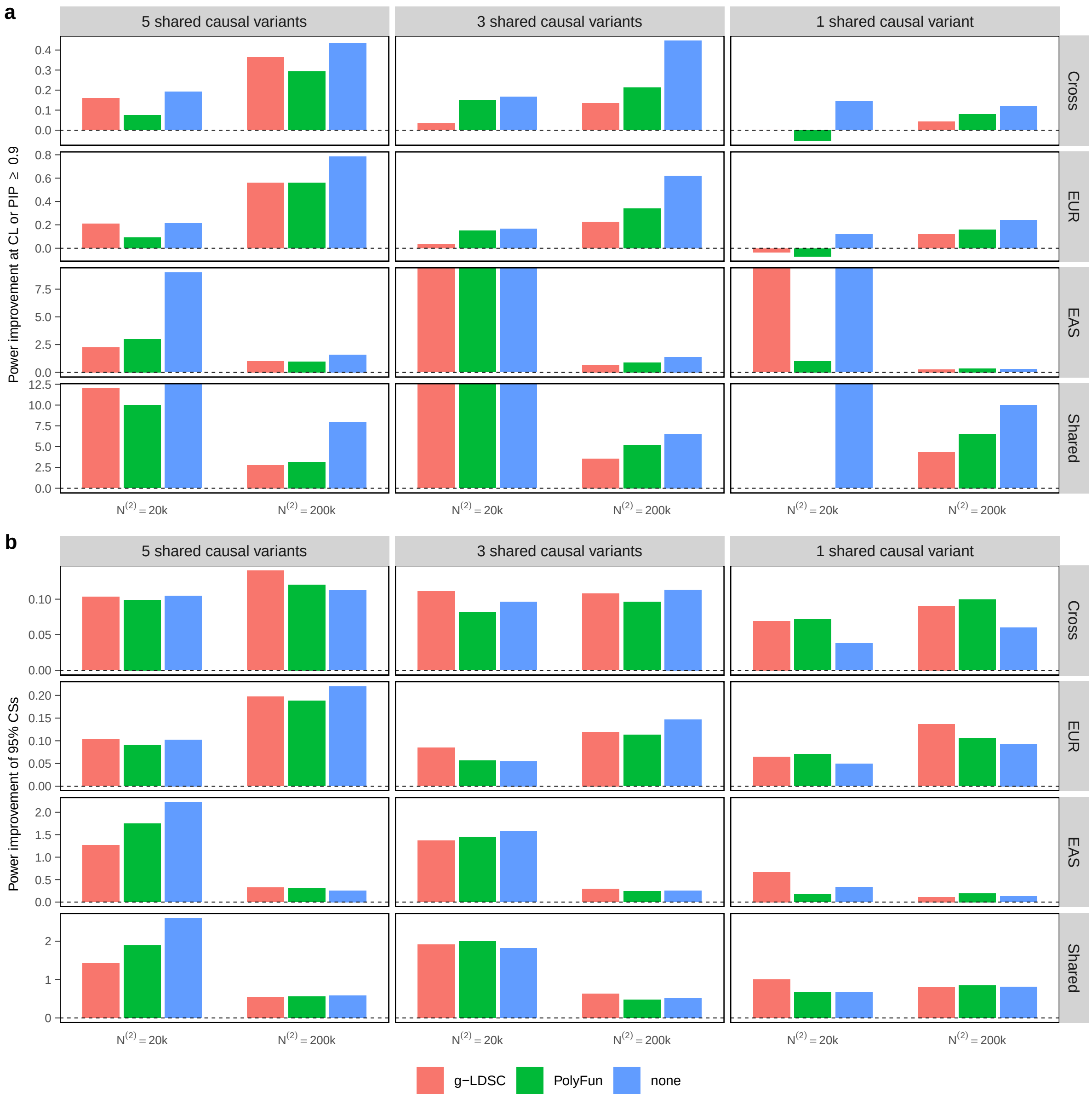


**Supplementary Fig. 14: Improvement in power of MACHINE compared to h2-D2 when out-of-sample LD matrices are used.** Power improvement is defined as $(\text{Power}^{(\text{MACHINE})} - \text{Power}^{(\text{h2-D2})})/\text{Power}^{(\text{h2-D2})}$. Numerical results are available in Supplementary Table 4.

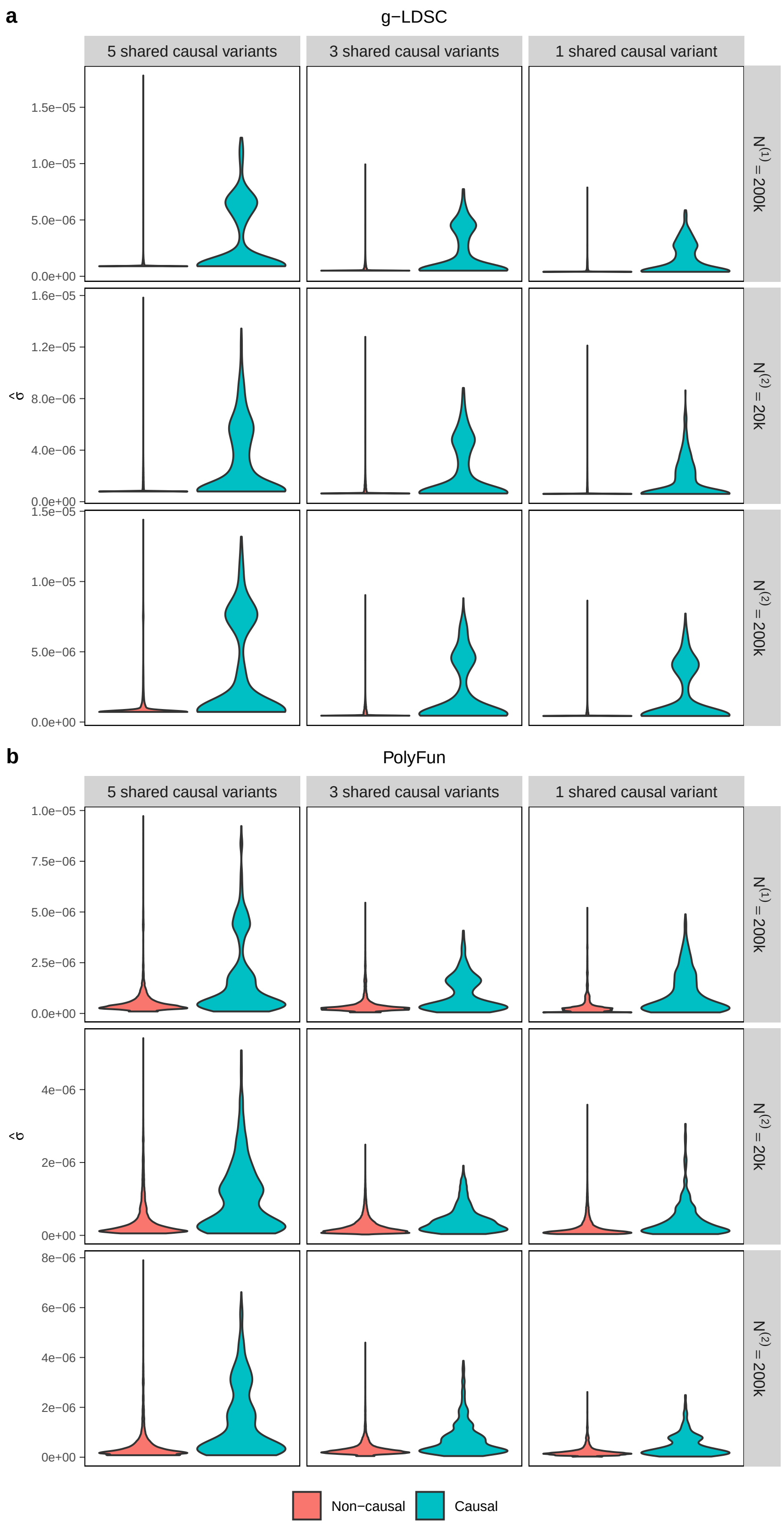


**Supplementary Fig. 15: Violin plots display the distribution of per-variant heritabilities estimated by (a) g-LDSC and (b) PolyFun for underlying causal and non-causal variants when out-of-sample LD matrices are used.** For each simulation setting, results are aggregated across 200 simulated datasets.

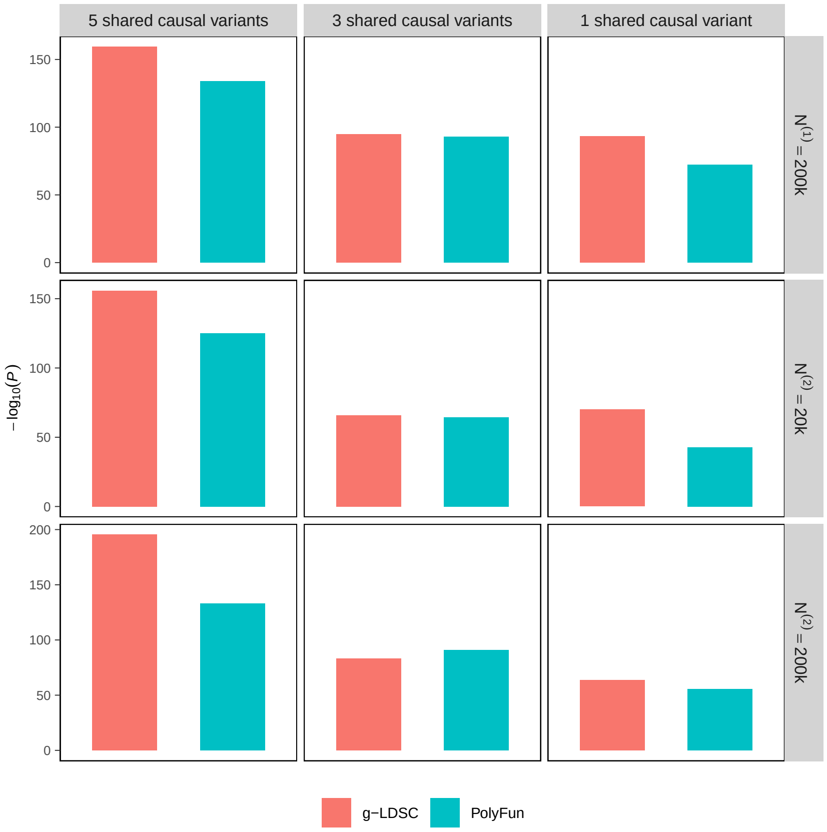


**Supplementary Fig. 16: Negative $\log_{10}(P$ value) of Wilcoxon-Mann-Whitney tests comparing estimated per-variant heritabilities between causal and non-causal variants when out-of-sample LD matrices are used.** For each simulation setting, results are aggregated across 200 simulated datasets. Numerical results are available in Supplementary Table 5.

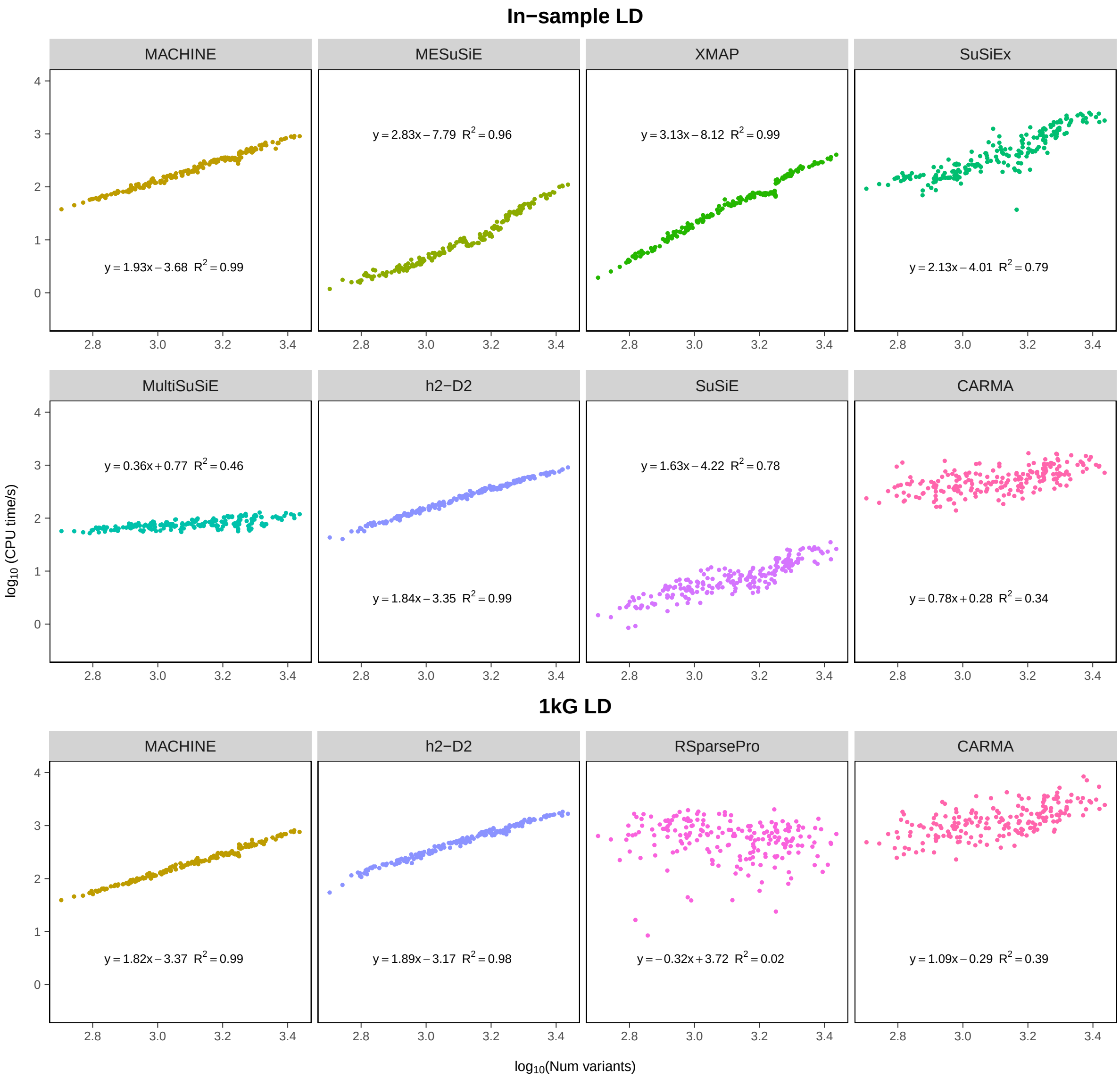


**Supplementary Fig. 17: Comparison of computational time in simulation studies.** The scatterplot depicts the CPU time (in seconds) versus the number of variants per locus, both on a logarithmic scale. For each method, a linear regression model was fitted with $\log_{10}$(CPU time/s) as the response variable and $\log_{10}$(number of variants) as the predictor. The fitted regression models and corresponding $R^2$ values are displayed. Numerical results are available in Supplementary Table 7.

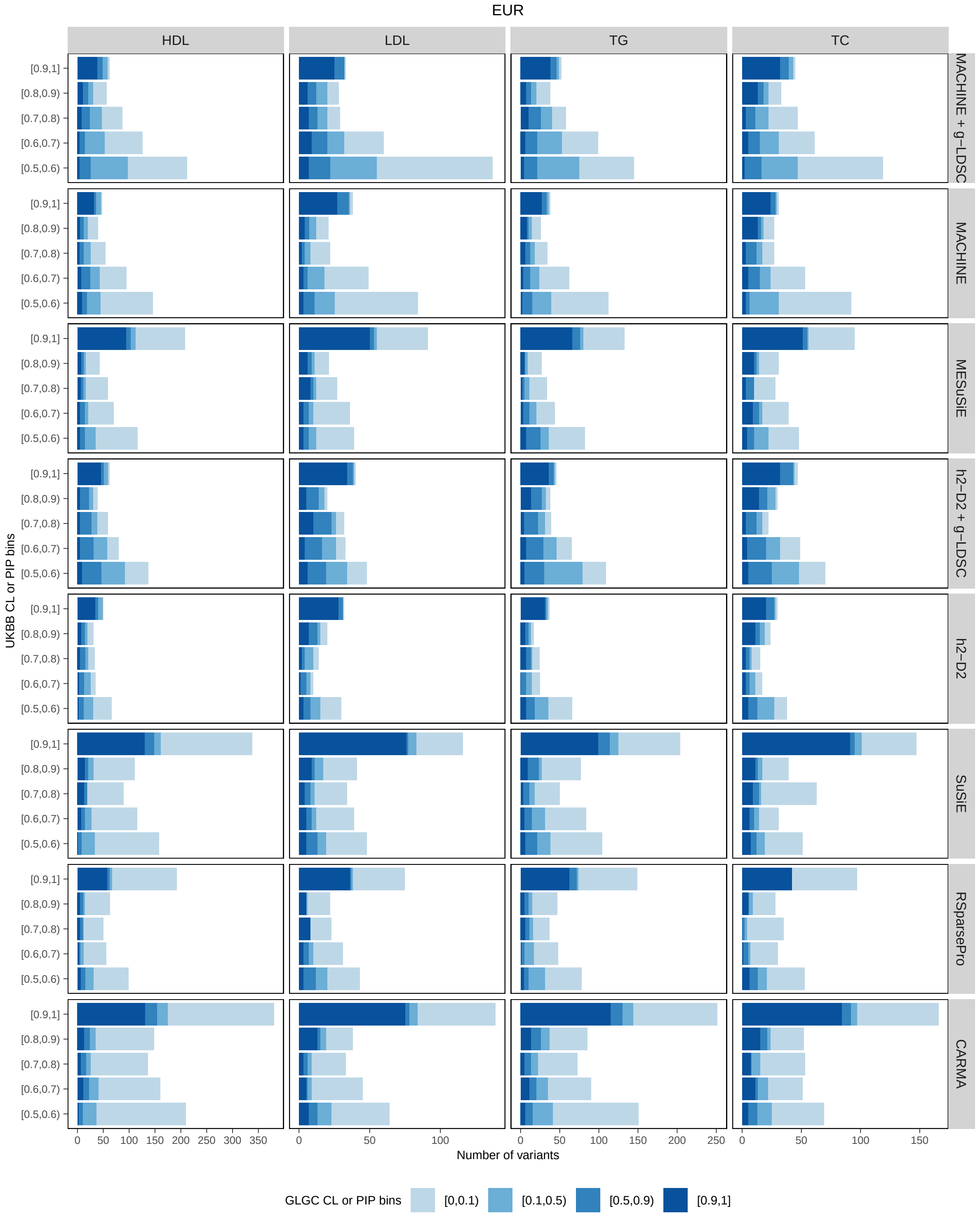


**Supplementary Fig. 18: Comparison of CL and PIP distributions between GLGC fine mapping and UKBB fine mapping for four lipid traits in EUR.** For each CL or PIP bin defined by GLGC fine mapping, the corresponding distributions of CL or PIP from UKBB fine mapping are presented. Numerical results are available in Supplementary Table 10.

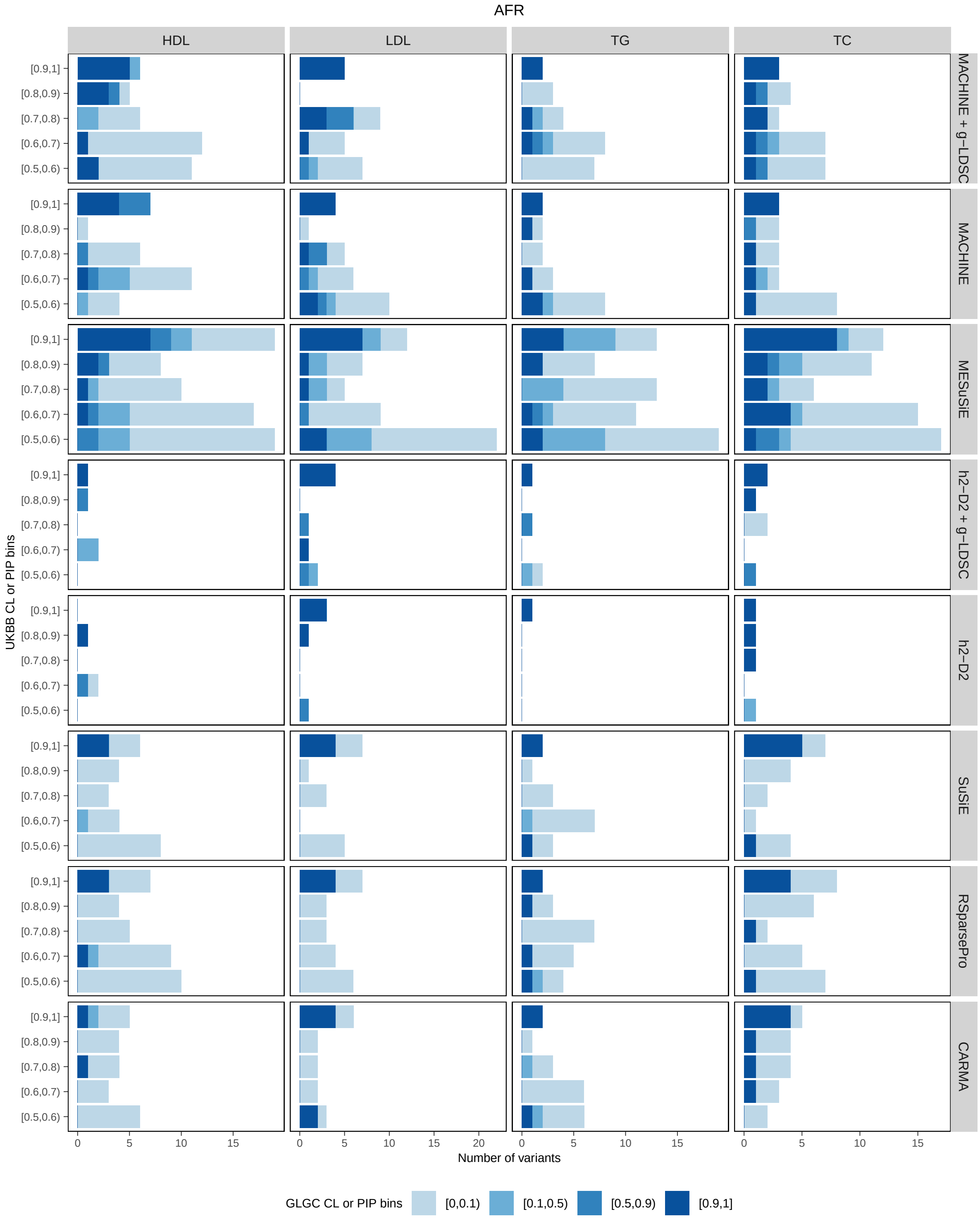


**Supplementary Fig. 19: Comparison of CL and PIP distributions between GLGC fine mapping and UKBB fine mapping for four lipid traits in AFR.** For each CL or PIP bin defined by GLGC fine mapping, the corresponding distributions of CL or PIP from UKBB fine mapping are presented. Numerical results are available in Supplementary Table 10.

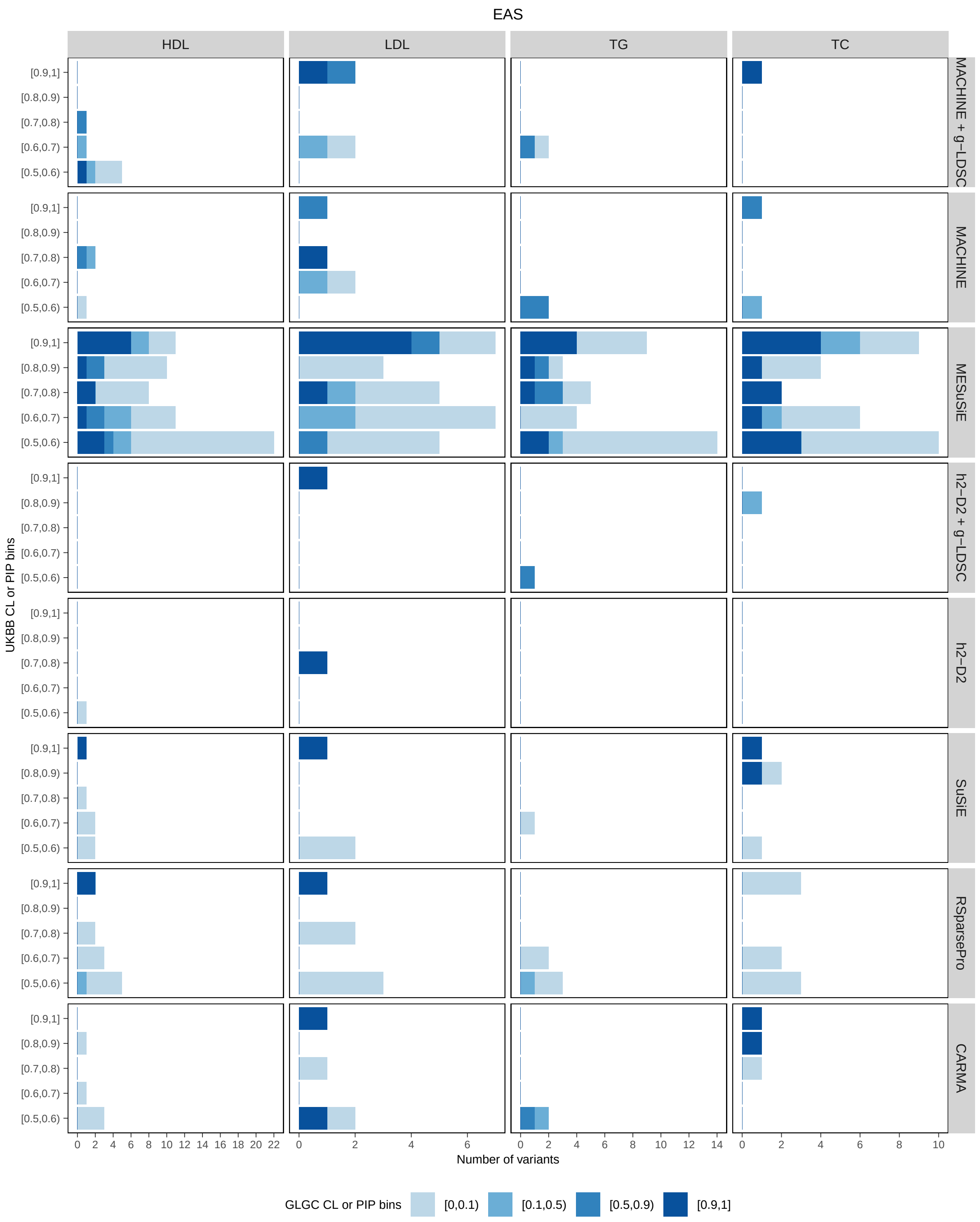


**Supplementary Fig. 20: Comparison of CL and PIP distributions between GLGC fine mapping and UKBB fine mapping for four lipid traits in EAS.** For each CL or PIP bin defined by GLGC fine mapping, the corresponding distributions of CL or PIP from UKBB fine mapping are presented. Numerical results are available in Supplementary Table 10.

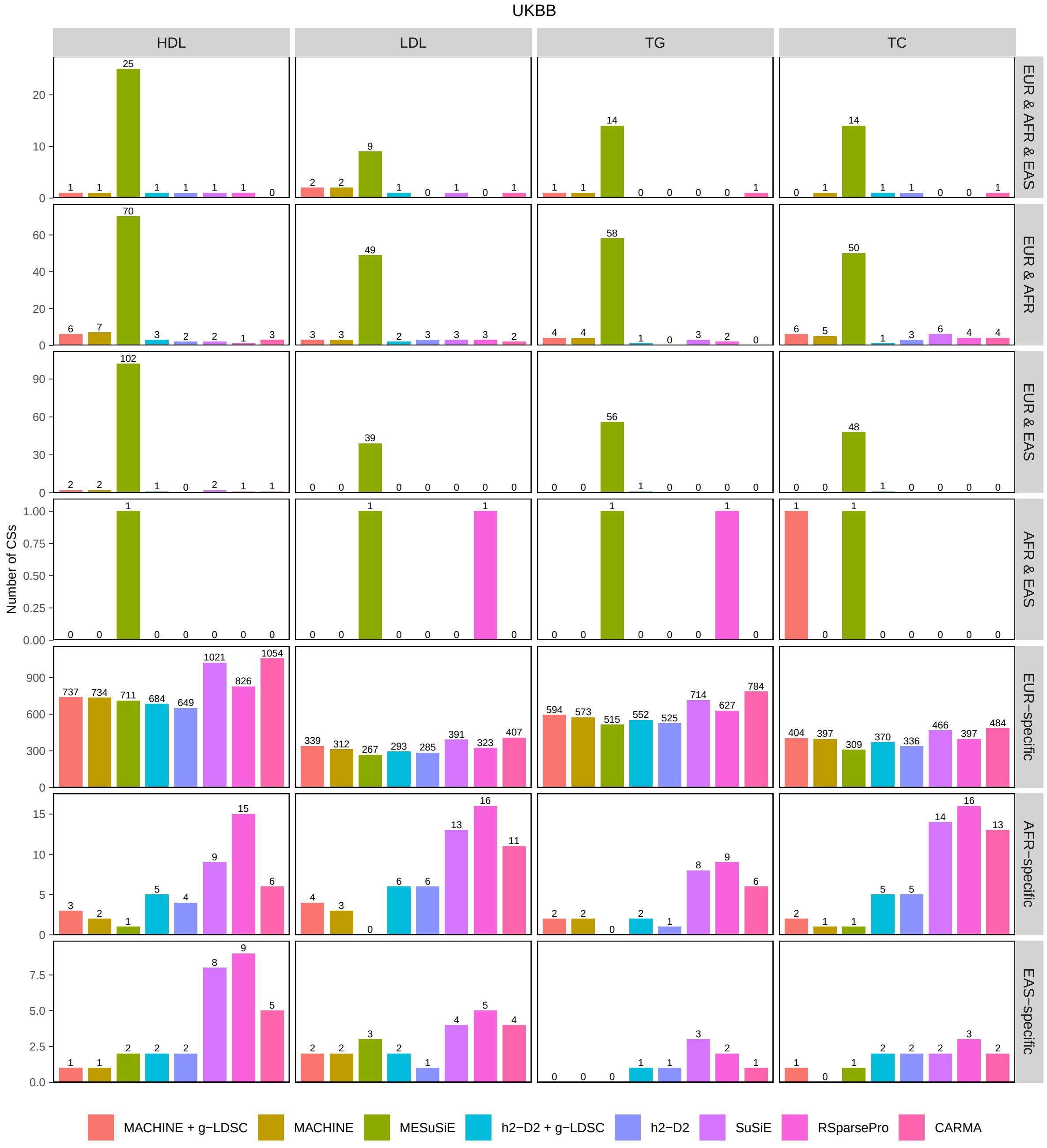


**Supplementary Fig. 21: Number of shared and ancestry-specific 95% CSs identified by each method using UKBB summary statistics for four lipit traits.** Numerical results are available in Supplementary Table 11.

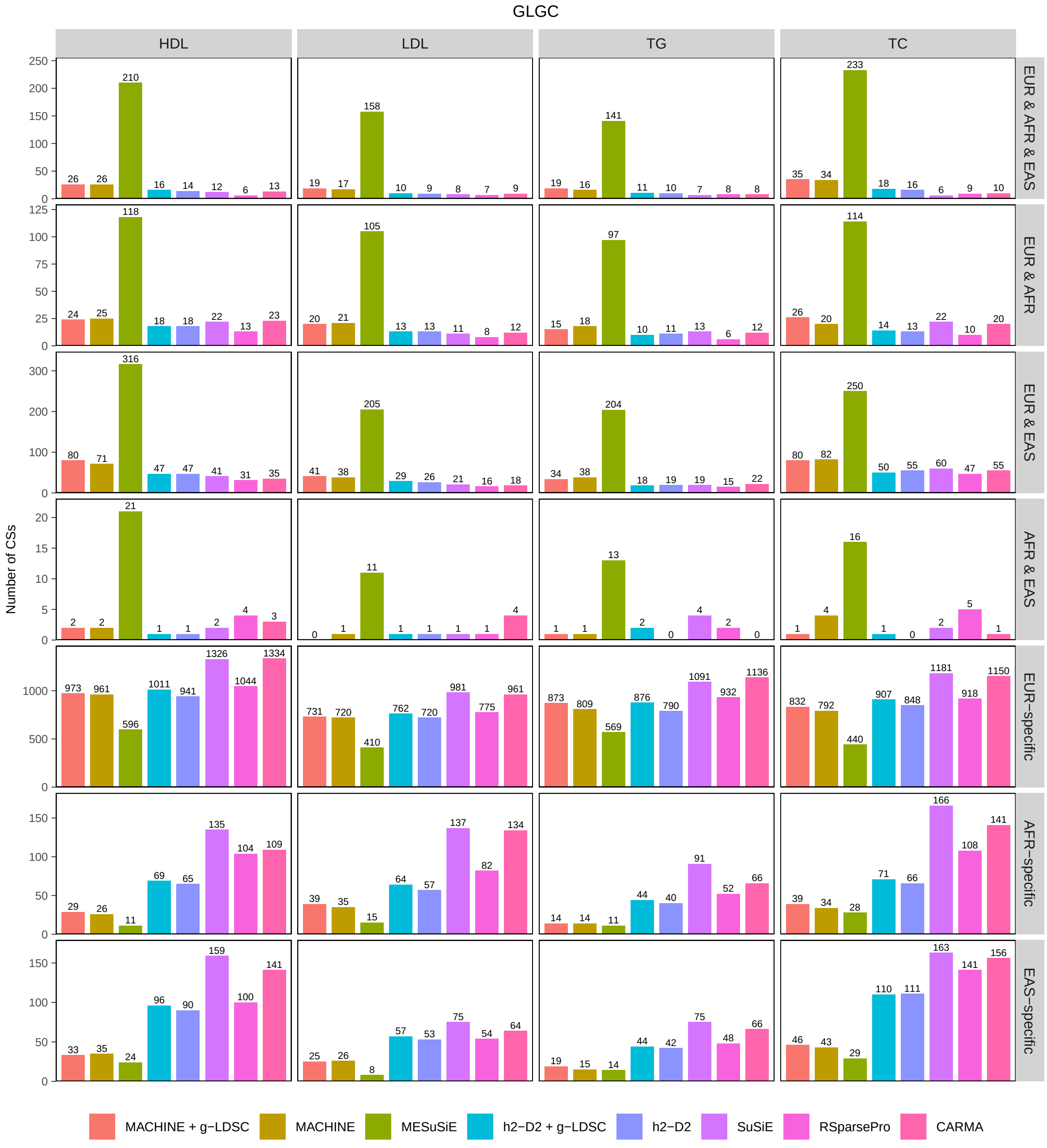


**Supplementary Fig. 22: Number of shared and ancestry-specific 95% CSs identified by each method using GLGC meta-analysis summary statistics for four lipit traits.** Numerical results are available in Supplementary Table 11.

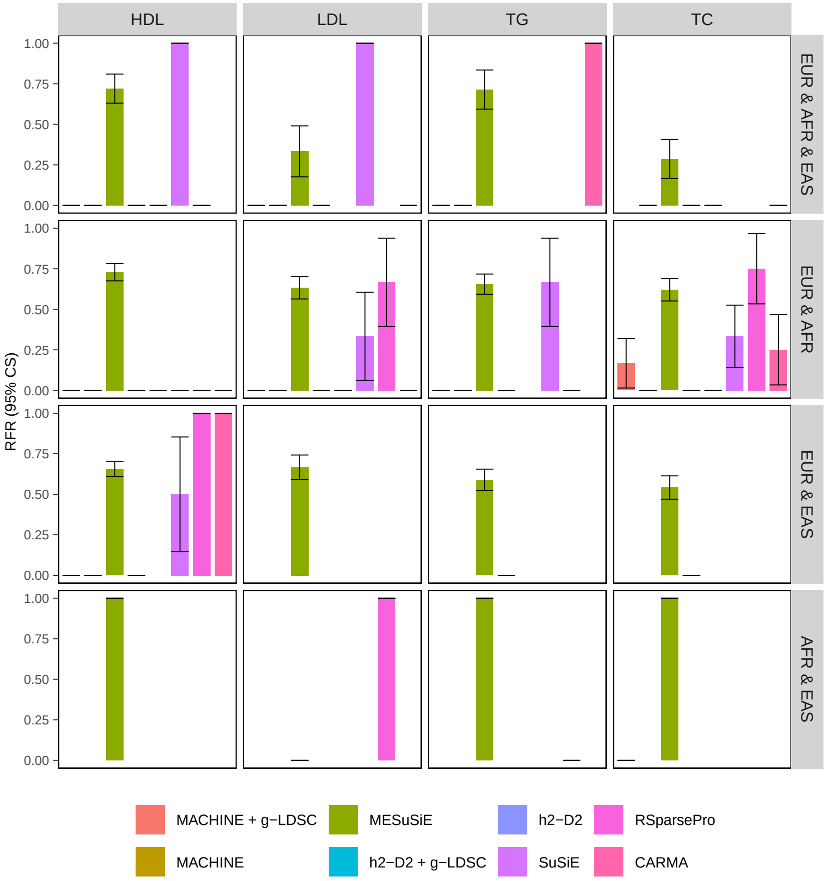


**Supplementary Fig. 23: RFRs for 95% CSs shared by two or three ancestries.** RFR is defined as the proportion of 95% CSs shared by two or three ancestries identified in UKBB fine mapping that do not overlap with any 95% CS shared by these ancestries identified in GLGC fine mapping. Numerical results are available in Supplementary Table 12.

### Supplementary Tables

**Supplementary Table 1: List of 200 blocks on chr1 selected for simulation studies.** For each block, the 1-based start coordinate and end coordinate in GRCh37 are shown (both positions are inclusive).

**Supplementary Table 2: Numerical results of simulations.** The columns are (1) LD: LD matrices used; (2) scenario: simulation scenario (1, 2, or 3); (3) N2: sample size of EAS; (4) POP: Cross for cross-ancestry causal variants, EUR for EUR causal variants, EAS for EAS causal variants, Shared for shared causal variants; (5) method: method name; (6) nVar_0.9: number of variants with CL or PIP ⩾ 0.9; (7) FDR_0.9: proportion of non-causal variants among variants with CL or PIP ⩾ 0.9; (8) FDR_0.9_sd: s.d. of FDR_0.9; (9) power_0.9: proportion of causal variants with CL or PIP ⩾ 0.9; (10) power_0.9_sd: s.d. of power_0.9; (11) n_CS95: total number of identified 95% CSs across 200 simulated datasets; (12) n_CS95_sd: s.d. of the number of identified 95% CSs across 200 simulated datasets; (13) coverage_CS95: the proportion of CSs that contain at least one causal variant; (14) coverage_CS95_sd: s.d. of coverage_CS95; (15) power_CS95: the proportion of causal variants included in the 95% CSs; (16) power_CS95_sd: s.d. of power_CS95; (17) size_CS95: average size of 95% CSs; (18) size_sd: s.d. of size across 95% CSs; (19) purity_CS95: average purity of 95% CSs; (20) purity_sd: s.d. of purity across 95% CSs.

**Supplementary Table 3: Numerical results of calibration in simulations.** The columns are (1) LD: LD matrices used; (2) scenario: simulation scenario (1, 2, or 3); (3) N2: sample size of EAS; (4) POP: Cross for cross-ancestry causal variants, EUR for EUR causal variants, EAS for EAS causal variants, Shared for shared causal variants; (5) method: method name; (6) group: CL or PIP bin; (7) nSNPs: number of variants within bin; (8) Expected: the average CL or PIP within bin; (9) Prop: proportion of causal variants within bin; (10) Prop_sd: s.d. of Prop.

**Supplementary Table 4: Numerical results of improvement in power of MACHINE compared to h2-D2 calibration in simulations.** The columns are (1) LD: LD matrices used; (2) scenario: simulation scenario (1, 2, or 3); (3) N2: sample size of EAS; (4) POP: Cross for cross-ancestry causal variants, EUR for EUR causal variants, EAS for EAS causal variants, Shared for shared causal variants; (5) anno_method: method for incorporating functional annotations; (6) h2-D2_0.9: power of CL⩾ 0.9 for h2-D2; (7) MACHINE_0.9: power of CL⩾ 0.9 for MACHINE; (8) improve_0.9: improvement in power of CL⩾ 0.9; (9) h2-D2_CS95: power of 95% CSs for h2-D2; (10) MACHINE_CS95: power of 95% CSs for MACHINE; (11) improve_CS95: improvement in power of 95% CSs.

**Supplementary Table 5: P values of Wilcoxon-Mann-Whitney tests comparing estimated per-variant heritabilities between causal and non-causal variants.** The columns are (1) LD: LD matrices used; (2) scenario: simulation scenario (1, 2, or 3); (3) setting: the combination of ancestry and sample size; (4) anno_method: method for incorporating functional annotations; (5) p: P values of Wilcoxon-Mann-Whitney tests.

**Supplementary Table 6: Coverage of 95% CSs grouped by the CS size.** The columns are (1) LD: LD matrices used; (2) scenario: simulation scenario (1, 2, or 3); (3) N2: sample size of EAS; (4) POP: Cross for cross-ancestry causal variants, EUR for EUR causal variants, EAS for EAS causal variants, Shared for shared causal variants; (5) method: method name; (6) size: size group; (7) n_CS95: total number of identified 95% CSs across 200 simulated datasets within the group; (8) coverage: the proportion of 95% CSs within the group that contain at least one causal variant; (9) coverage_sd: s.d. of coverage.

**Supplementary Table 7: Information of fitted linear regression models with** $\log_{10}$**(CPU time/s) as the response variable and** $\log_{10}$**(number of variants) as the predictor.** The columns are (1) LD: LD matrices used; (2) method: method name; (3) intercept: intercept of the fitted linear model; (4) intercept_sd: s.d. of intercept; (5) slope: slope of the fitted linear model; (6) slope_sd: s.d. of slope; (7) adj.r.squared: adjusted $R^2$ value of the fitted linear model.

**Supplementary Table 8: Sample size of each lipid trait in each ancestry.** The columns are (1) pheno: name of lipid trait; (2) pid: ancestry ID; (3) UKBB.n: sample size of UKBB summary data; (4) GLGC.n: sample size of GLGC summary data.

**Supplementary Table 9: RFRs of fine-mapping methods in real data analysis of four lipid traits.** The columns are (1) pheno: name of lipid trait; (2) pid: ancestry ID; (3) method: method name; (4) num_variants_0.9.UKBB: number of variants with CL or PIP ⩾ 0.9 in UKBB fine mapping; (5) num_variants_0.9.UKBB_0.1.GLGC: number of variants with CL or PIP ⩾ 0.9 in UKBB fine mapping and < 0.1 in GLGC fine mapping; (6) RFR_0.9: RFR for variants with CL or PIP ⩾ 0.9 in UKBB fine mapping; (7) RFR_0.9_sd: s.d. of RFR_0.9; (8) UKBB_nCS: number of 95% CSs identified in UKBB fine mapping; (9) UKBB_in_GLGC: number of 95% CSs identified in UKBB fine mapping that overlap with any 95% CSs identified in GLGC fine mapping; (10) RFR_CS95: RFR for 95% CSs; (11) RFR_CS95_sd: s.d. of RFR_CS95.

**Supplementary Table 10: Comparison of CL and PIP distributions between GLGC fine mapping and UKBB fine mapping for four lipid traits.** The columns are (1) pheno: name of lipid trait; (2) pid: ancestry ID; (3) method: method name; (4) group_UKBB: CL or PIP bin of UKBB fine mapping; (5) group_GLGC: CL or PIP bin of GLGC fine mapping; (6) nVar: number of variants within bins.

**Supplementary Table 11: Number of shared and ancestry-specific 95% CSs identified by each method for four lipid traits.** The columns are (1) db: database of GWAS summary data; (2) pheno: name of lipid trait; (3) method: method name; (4)-(10): number of shared and ancestry-specific 95% CSs.

**Supplementary Table 12: RFRs for 95% CSs shared by two or three ancestries for four lipid traits.** The columns are (1) pheno: name of lipid trait; (2) method: method name; (3) pops: combination of ancestries; (4) nCS_common: number of shared 95% CSs identified in UKBB fine mapping that overlap with any shared 95% CSs identified in GLGC fine mapping; (5) RFR_CS95: RFR for shared 95% CSs; (6) RFR_CS95_sd: s.d. of RFR_CS95.

**Supplementary Table 13: Enrichment of functionally important variants in high-confidence variants and 95% CSs identified by each method for four lipid traits.** The columns are (1) db: database of GWAS summary data; (2) pheno: name of lipid trait; (3) method: method name; (4) num_variants: total number of variants included in fine-mapping analyses; (5) num_variants_0.9: number of high-confidence variants (CL or PIP $\geqslant$ 0.9); (6) num_CS95: number of identified 95% CSs; (7) num_variants_eQTL: number of eQTLs among all variants included in fine-mapping analyses; (8) num_variants_0.9_eQTL: number of eQTLs among high-confidence variants; (9) num_CS95_eQTL: number of 95% CSs containing at least one eQTL; (10) num_variants_Coding: number of coding variants among all variants included in fine-mapping analyses; (11) num_variants_0.9_Coding: number of coding variants among high-confidence variants; (12) num_CS95_Coding: number of 95% CSs containing at least one coding variant; (13) num_variants_Regulatory: number of putative regulatory variants among all variants included in fine-mapping analyses; (14) num_variants_0.9_Regulatory: number of putative regulatory variants among high-confidence variants; (15) num_CS95_Regulatory: number of 95% CSs containing at least one putative regulatory variant.

**Supplementary Table 14: Number of shared and ancestry-specific 95% CSs identified by each method for SCZ.** The columns are (1) method: method name; (2) EUR_EAS: number of identified 95% CSs shared by EUR and EAS; (3) EUR-specific: number of identified EUR-specific 95% CSs; (4) EUR-specific: number of identified EAS-specific 95% CSs.

**Supplementary Table 15: Enrichment of functionally important variants in medium-high-confidence variants and 95% CSs identified by each method for SCZ.** The columns are (1) method: method name; (2) num_variants: total number of variants included in fine-mapping analyses; (3) num_variants_0.5: number of medium-high-confidence variants (CL or PIP $\geqslant 0.5$); (4) num_CS95: number of identified 95% CSs; (5) num_variants_eQTL: number of eQTLs among all variants included in fine-mapping analyses; (6) num_variants_0.5_eQTL: number of eQTLs among medium-high-confidence variants; (7) num_CS95_eQTL: number of 95% CSs containing at least one eQTL; (8) num_variants_Coding: number of coding variants among all variants included in fine-mapping analyses; (9) num_variants_0.5_Coding: number of coding variants among medium-high-confidence variants; (10) num_CS95_Coding: number of 95% CSs containing at least one coding variant; (11) num_variants_Regulatory: number of putative regulatory variants among all variants included in fine-mapping analyses; (12) num_variants_0.5_Regulatory: number of putative regulatory variants among medium-high-confidence variants; (13) num_CS95_Regulatory: number of 95% CSs containing at least one putative regulatory variant.

### Supplementary Note

#### MCMC sampling algorithm

For $k \in \{1, \ldots, K\}$ and $j \in \{1, \ldots, J\}$, we introduce a latent variable $\psi_j^{(k)} \sim \text{Exp}(1)$, such that the Laplace prior distribution $\beta_j^{(k)} \mid \sigma_j^{(k)} \sim \text{DE}\left(0, \sqrt{\sigma_j^{(k)}/2}\right)$ can be expressed as

$$\begin{aligned}
\beta_j^{(k)} \mid \psi_j^{(k)}, \sigma_j^{(k)} \sim & \mathrm{N}\left(0, \psi_j^{(k)} \sigma_j^{(k)}\right), \\
\psi_j^{(k)} \sim & \text{Exp}(1).
\end{aligned}$$

Let $\gamma_j^{(k)} = \log\left(\sigma_j^{(k)}/K_j\right)$. The prior distribution of $\mathbf{\Gamma} = \left\{\gamma_j^{(k)}\right\}$ can be expressed as

$$\begin{aligned}
p(\mathbf{\Gamma}) \propto & \prod_{\substack{j:K_j=1 \\ k:\gamma_j^{(k)}=1}} \exp\left\{a_j \gamma_j^{(k)}\right\} \times \prod_{j:K_j>1} \left\{\left(\sum_{k \in \mathcal{K}_j} \mathrm{e}^{\gamma_j^{(k)}}\right)^{a_j - \sum_{k \in \mathcal{K}_j} c_j^{(k)}} \prod_{k \in \mathcal{K}_j} \exp\left\{c_j^{(k)} \gamma_j^{(k)}\right\}\right\} \\
& \times \left(1 - \sum_{j=1}^{J} \sum_{k \in \mathcal{K}_j} \mathrm{e}^{\gamma_j^{(k)}}\right)^{b-1} \times \mathbf{1}\left\{\sum_{j=1}^{J} \sum_{k \in \mathcal{K}_j} \mathrm{e}^{\gamma_j^{(k)}} \leqslant 1\right\}.
\end{aligned}$$

Let

$$\begin{aligned}
\boldsymbol{\mu}^{(k)} &= \sqrt{N^{(k)}} \boldsymbol{R}^{(k)} \left(\boldsymbol{R}^{(k)} + \widehat{\lambda}^{(k)} \boldsymbol{I}_{J^{(k)}}\right)^{-1} \boldsymbol{z}^{(k)} \\
&= \sqrt{N^{(k)}} \boldsymbol{U}^{(k)} \boldsymbol{D}^{(k)} \left(\boldsymbol{D}^{(k)} + \widehat{\lambda}^{(k)} \boldsymbol{I}_{J^{(k)}}\right)^{-1} \left(\boldsymbol{U}^{(k)}\right)^{\top} \boldsymbol{z}^{(k)}, \\
\boldsymbol{W}^{(k)} &= N^{(k)} \boldsymbol{R}^{(k)} \left(\boldsymbol{R}^{(k)} + \widehat{\lambda}^{(k)} \boldsymbol{I}_{J^{(k)}}\right)^{-1} \boldsymbol{R}^{(k)} \\
&= N^{(k)} \boldsymbol{U}^{(k)} \boldsymbol{D}^{(k)} \left(\boldsymbol{D}^{(k)} + \widehat{\lambda}^{(k)} \boldsymbol{I}_{J^{(k)}}\right)^{-1} \boldsymbol{D}^{(k)} \left(\boldsymbol{U}^{(k)}\right)^{\top}.
\end{aligned}$$

The complete data likelihood is

$$\begin{aligned}
& p(\boldsymbol{Z}, \boldsymbol{B}, \boldsymbol{T}, \boldsymbol{\Psi}) \\
\propto & \prod_{k=1}^{K} \exp\left\{\boldsymbol{\mu}^{(k)\top} \boldsymbol{\beta}^{(k)} - \frac{1}{2} \boldsymbol{\beta}^{(k)\top} \boldsymbol{W}^{(k)} \boldsymbol{\beta}^{(k)}\right\} \\
& \times \prod_{j=1}^{J} \prod_{k \in \mathcal{K}_j} \left(\psi_j^{(k)} \mathrm{e}^{\gamma_j^{(k)}}\right)^{-\frac{1}{2}} \exp\left\{-\frac{\left(\beta_j^{(k)}\right)^2}{2K_j \psi_j^{(k)} \mathrm{e}^{\gamma_j^{(k)}}}\right\} \times \prod_{j=1}^{J} \prod_{k \in \mathcal{K}_j} \exp\left\{-\psi_j^{(k)}\right\} \\
& \times \prod_{\substack{j:K_j=1 \\ k:\gamma_j^{(k)}=1}} \exp\left\{a_j \gamma_j^{(k)}\right\} \times \prod_{j:K_j>1} \left\{\left(\sum_{k \in \mathcal{K}_j} \mathrm{e}^{\gamma_j^{(k)}}\right)^{a_j - \sum_{k \in \mathcal{K}_j} c_j^{(k)}} \prod_{k \in \mathcal{K}_j} \exp\left\{c_j^{(k)} \gamma_j^{(k)}\right\}\right\} \\
& \times \left(1 - \sum_{j=1}^{J} \sum_{k \in \mathcal{K}_j} \mathrm{e}^{\gamma_j^{(k)}}\right)^{b-1} \times \mathbf{1}\left\{\sum_{j=1}^{J} \sum_{k \in \mathcal{K}_j} \mathrm{e}^{\gamma_j^{(k)}} \leqslant 1\right\}.
\end{aligned}$$

We propose the following MCMC algorithm to sample parameters from the posterior distribution:

(i) Set initial values for $\boldsymbol{B}$, $\boldsymbol{T}$, and $\boldsymbol{\Psi}$.

(ii) For each $j \in \{1, \ldots, J\}$ and each $k \in \mathcal{K}_j$:

(i) Update $\gamma_j^{(k)}$ using Metropolis-Hastings algorithm according to the unnormalized density

$$
\begin{aligned}
& p\left(\gamma_j^{(k)} \mid \boldsymbol{\beta}_{-j}^{(k)}, \psi_j^{(k)}, \boldsymbol{T} \backslash \left\{\gamma_j^{(k)}\right\}, z_j^{(k)}\right) \\
\propto & \sqrt{\widetilde{\sigma}_j^{(k)}} \times \exp\left\{\frac{1}{2} \widetilde{\sigma}_j^{(k)} \left(u_j^{(k)}\right)^2\right\} \\
& \times \exp\left\{\left(c_j^{(k)} - \frac{1}{2}\right) \gamma_j^{(k)}\right\} \times \left(\mathrm{e}^{\gamma_j^{(k)}} + \sum_{k' \in \mathcal{K}_j \backslash \{k\}} \mathrm{e}^{\gamma_j^{(k')}}\right)^{a_j - \sum_{k' \in \mathcal{K}_j} c_j^{(k')}} \\
& \times \left(1 - \mathrm{e}^{\gamma_j^{(k)}} - \sum_{(k', j') \neq (k, j)} \mathrm{e}^{\gamma_{j'}^{(k')}}\right)^{b-1} \\
& \times \mathbf{1}\left\{\gamma_j^{(k)} \leqslant \ln\left(1 - \sum_{(k', j') \neq (k, j)} \mathrm{e}^{\gamma_{j'}^{(k')}}\right)\right\},
\end{aligned}
$$

if $K_j > 1$, otherwise

$$
\begin{aligned}
& p\left(\gamma_j^{(k)} \mid \boldsymbol{\beta}_{-j}^{(k)}, \psi_j^{(k)}, \boldsymbol{T} \backslash \left\{\gamma_j^{(k)}\right\}, z_j^{(k)}\right) \\
\propto & \sqrt{\widetilde{\sigma}_j^{(k)}} \times \exp\left\{\frac{1}{2} \widetilde{\sigma}_j^{(k)} \left(u_j^{(k)}\right)^2\right\} \\
& \times \exp\left\{\left(a_j - \frac{1}{2}\right) \gamma_j^{(k)}\right\} \times \left(1 - \mathrm{e}^{\gamma_j^{(k)}} - \sum_{(k', j') \neq (k, j)} \mathrm{e}^{\gamma_{j'}^{(k')}}\right)^{b-1} \\
& \times \mathbf{1}\left\{\gamma_j^{(k)} \leqslant \ln\left(1 - \sum_{(k', j') \neq (k, j)} \mathrm{e}^{\gamma_{j'}^{(k')}}\right)\right\},
\end{aligned}
$$

where

$$
\widetilde{\sigma}_j^{(k)} = \left[W_{jj}^{(k)} + \frac{1}{\psi_j^{(k)} \mathrm{e}^{\gamma_j^{(k)}}}\right]^{-1}, u_j^{(k)} = \mu_j^{(k)} - \sum_{j' \neq j} W_{jj'}^{(k)} \beta_j^{(k)}.
$$

(ii) Update $\beta_j^{(k)} \sim \mathrm{N}\left(\widetilde{\sigma}_j^{(k)} u_j^{(k)}, \widetilde{\sigma}_j^{(k)}\right)$.

(iii) Update

$$
\frac{1}{\psi_j^{(k)}} \mid \beta_j, \gamma_j \sim \text{InvGaussian}\left(\text{mean} = \frac{\sqrt{2} \mathrm{e}^{\gamma_j^{(k)}/2}}{\left|\beta_j^{(k)}\right|}, \text{shape} = 2\right).
$$

(iii) To improve mixing of MCMC chain, after every 5 steps, we will choose several pairs of SNPs with high LD in at least one ancestry and propose a proposal by switching the values of each pair. For a given pair $j_1$ and $j_2$ with high LD in the $k$-th ancestry, the proposal is given by setting

$$\gamma_{j_1,\text{new}}^{(k)} = \gamma_{j_2}^{(k)}, \gamma_{j_2,\text{new}}^{(k)} = \gamma_{j_1}^{(k)}, \psi_{j_1,\text{new}}^{(k)} = \psi_{j_2}^{(k)} K_{j_2}/K_{j_1}, \psi_{j_2,\text{new}}^{(k)} = \psi_{j_1}^{(k)} K_{j_1}/K_{j_2},$$

and

$$\beta_{j_1,\text{new}}^{(k)} = \beta_{j_2}^{(k)}, \beta_{j_2,\text{new}}^{(k)} = \beta_{j_1}^{(k)},$$

if $R_{j_1,j_2}^{(k)} > 0$, or

$$\beta_{j_1,\text{new}}^{(k)} = -\beta_{j_2}^{(k)}, \beta_{j_2,\text{new}}^{(k)} = -\beta_{j_1}^{(k)},$$

if $R_{j_1,j_2}^{(k)} < 0$. Let $s = \text{sgn}\left(R_{j_1,j_2}^{(k)}\right)$. The acceptance probability is

$$\begin{aligned}
&\exp\left\{\left[\sum_{l\in\mathcal{K}_j\setminus\{j_1,j_2\}} \left(W_{l,j_1}^{(k)} - sW_{l,j_2}^{(k)}\right)\beta_l^{(k)} - \mu_{j_1}^{(k)} + s\mu_{j_2}^{(k)}\right]\left(\beta_{j_1}^{(k)} - s\beta_{j_2}^{(k)}\right)\right\} \\
&\times \exp\left\{\frac{1}{2}\left(W_{j_1,j_1}^{(k)} - W_{j_2,j_2}^{(k)}\right)\left[\left(\beta_{j_1}^{(k)}\right)^2 - \left(\beta_{j_2}^{(k)}\right)^2\right]\right\} \\
&\times \exp\left\{\left(c_{j_1}^{(k)} - c_{j_2}^{(k)}\right)\left(\gamma_{j_2}^{(k)} - \gamma_{j_1}^{(k)}\right)\right\} \\
&\times \left\{\frac{\sum_{k'\in\mathcal{K}_{j_1}\setminus\{k\}} e^{\gamma_{j_1}^{(k')}} + e^{\gamma_{j_2}^{(k)}}}{\sum_{k'\in\mathcal{K}_{j_1}\setminus\{k\}} e^{\gamma_{j_1}^{(k')}} + e^{\gamma_{j_1}^{(k)}}}\right\}^{a_{j_1} - \sum_{k'\in\mathcal{K}_{j_1}} c_{j_1}^{(k')}} \\
&\times \left\{\frac{\sum_{k'\in\mathcal{K}_{j_2}\setminus\{k\}} e^{\gamma_{j_2}^{(k')}} + e^{\gamma_{j_1}^{(k)}}}{\sum_{k'\in\mathcal{K}_{j_2}\setminus\{k\}} e^{\gamma_{j_2}^{(k')}} + e^{\gamma_{j_2}^{(k)}}}\right\}^{a_{j_2} - \sum_{k'\in\mathcal{K}_{j_2}} c_{j_2}^{(k')}} \\
&\times \exp\left\{-\psi_{j_1}^{(k)} K_{j_1}/K_{j_2} - \psi_{j_2}^{(k)} K_{j_2}/K_{j_1} + \psi_{j_1}^{(k)} + \psi_{j_2}^{(k)}\right\} \wedge 1.
\end{aligned}$$

(iv) Repeat Steps (ii)-(iii) until convergence.

### Choice of hyper-parameter $b$

The choice of hyper-parameter $b$ is based on the relationship between hyper-parameters and the prior expectation of local heritability. The averaged prior expectation of local heritability across ancestries is

$$\begin{aligned}
\frac{1}{K}\sum_{k=1}^{K} \mathrm{E}(h^{(k)}) =& \frac{1}{K}\sum_{k=1}^{K}\sum_{j\in\mathcal{J}^{(k)}} K_j \mathrm{E}(\xi_j)\,\mathrm{E}\left(\eta_j^{(k)}\right) \\
=& \frac{1}{K}\frac{1}{\sum_{j=1}^{J} a_j + b}\sum_{k=1}^{K}\sum_{j\in\mathcal{J}^{(k)}} K_j a_j \frac{c_j^{(k)}}{\sum_{k'\in\mathcal{K}_j} c_j^{(k')}} = \frac{A^*}{A+b},
\end{aligned}$$

where $A = \sum_{j=1}^{J} a_j$ and

$$A^* = \frac{1}{K} \sum_{k=1}^{K} \sum_{j \in \mathcal{J}^{(k)}} K_j a_j \frac{c_j^{(k)}}{\sum_{k' \in \mathcal{K}_j} c_j^{(k')}}.$$

The procedure of specifying $b$ is:

(i) Set initial $h_{\text{est}} = 10^{-4}$ and set $b = A^*/h_{\text{est}} - A$.

(ii) Run MCMC algorithm for $n_{\text{mcmc}}$ (default 400) steps. Discard the first $n_{\text{burnin}}$ (default 200) samples as burn in.

(iii) For $n \in \{n_{\text{burnin}} + 1, \ldots, n_{\text{mcmc}}\}$, let $\boldsymbol{B}_n = \left[\boldsymbol{\beta}_n^{(1)}, \ldots, \boldsymbol{\beta}_n^{(K)}\right]$ denote the $n$-th MCMC sample of $\boldsymbol{B}$. The $n$-th MCMC sample of the local heritability of the $k$-th ancestry is computed by $h_n^{(k)} = \left(\boldsymbol{\beta}_n^{(k)}\right)^\top \boldsymbol{R}^{(k)} \boldsymbol{\beta}_n^{(K)}$. Let

$$\overline{h}_n = \frac{1}{K} \sum_{k=1}^{K} h_n^{(k)}.$$

(iv) Perform a $t$-test to compare the mean of $\left\{\overline{h}_n \mid n \in \{n_{\text{burnin}} + 1, \ldots, n_{\text{mcmc}}\}\right\}$ with $h_{\text{est}}$. If the $P$ value is less than 0.05, set

$$h_{\text{est}} = \frac{1}{n_{\text{mcmc}} - n_{\text{burnin}}} \sum_{n=n_{\text{burnin}}+1}^{n_{\text{mcmc}}} \overline{h}_n,$$

$b = A^*/h_{\text{est}} - A$, and go back to step (ii). Otherwise, stop the algorithm.
